# Metabolic Drivers of Dysglycemia in Pregnancy: Ethnic-Specific GWAS of 146 Metabolites and 1-Sample Mendelian Randomisation Analyses in a UK Multi-Ethnic Birth Cohort

**DOI:** 10.1101/2022.12.21.22283768

**Authors:** Harriett Fuller, Mark M Iles, J. Bernadette Moore, Michael A. Zulyniak

## Abstract

Gestational diabetes mellitus (GDM) is the most common pregnancy complication worldwide and is associated with short- and long-term health implications for both mother and child. Prevalence of GDM varies between ethnicities, with South Asians (SAs) experiencing up to three times the risk compared to white Europeans (WEs). This study aimed to evaluate the causal role of metabolic characteristics in the ethnic-associated differences in gestational dysglycemia.

A one-sample Mendelian Randomisation (MR) was performed separately on 3688 SA and 3354 WE women (<28^th^ week of pregnancy) from the Born in Bradford (BiB) cohort for 146 metabolites exposures for the outcomes of fasting glucose and 2-hr post glucose (P ≤ 1 x 10^-5^ was considered significant). Additional GWAS and MR analyses on 22 composite measures of metabolite classes were also conducted.

Through an extensive GWAS analysis this study identified 15 novel genome-wide significant (GWS) SNPs associated with tyrosine in the *FOXN* and *SLC13A2* genes and 1 novel GWS SNP (currently in no known gene) associated with acetate in SAs. Through the utilisation of a MR analysis, 14 metabolites were found to be associated with postprandial glucose in WEs, while in SAs a distinct panel of 11 metabolites were identified. Furthermore, in WEs, cholesterols were most the common metabolite associated with dysglycemia, while in SAs saturated fatty acids were most common. Furthermore, in SAs a composite measure of the fatty acid class was also found to associate with 2-hour post glucose.

The presence of ethnic-specific causal relationships between a comprehensive set of metabolites and postprandial glucose measures (fasting glucose and 2-hour post glucose) in mid-pregnancy has been established in a UK SA and WE population. Future work should aim to investigate the biological mechanisms of metabolites on GDM risk and inform ethnically tailored GDM prevention strategies are required.

## Introduction

Pregnancy is accompanied by a period of intense maternal metabolic adaptation to meet the energy demands of the foetus [1–3]. Mild maternal insulin resistance (IR) is a natural adaption to prioritise adequate glucose for the growing foetus [4]. However, if gestational IR exceeds healthy levels and glycaemia is uncontrolled, moderate IR can progress to gestational diabetes mellitus (GDM) [1, 5]. GDM is characterised by persistent maternal and foetal exposure to elevated levels of glucose, and places the mother and offspring at risk during pregnancy (i.e., macrosomia and haemorrhaging) and in later life (i.e. from obesity, type 2 diabetes (T2D) and cardiovascular disease) [6].

Globally, GDM is the most common pregnancy complication, affecting up to one in every seven births; however, its prevalence varies between ethnic groups, with South Asian (SA) women at 3-fold greater risk compared to white European (WE) women, irrespective of BMI and country of residence [5, 7]. Furthermore, SA women are more likely to develop T2D in later life following a GDM diagnosis [8]. Factors driving this disparity in prevalence are not fully understood but metabolism is thought to play a key role [6], given emerging evidence demonstrating (i) differences in metabolic profiles between GDM and non-GDM pregnancies in an ethnic-specific manner [9, 10]; and (ii) that a single dietary strategy to manage GDM across all ethnic groups appears ineffective [11, 12]. However, heterogeneity of reported metabolite-GDM associations between studies (due to differences in quantification methods, GDM diagnostic criteria [13, 14], ethnic and cultural groups), as well as residual confounding in observational studies have prevented complete understanding and, moreover, advancement to improved equitable care. In short, the field requires a clear and accurate understanding of ethnic-specific metabolic drivers of gestational dysglycemia to inform appropriate and effective prevention and management strategies across ethnic groups.

Mendelian Randomisation (MR) is an instrumental variable technique that can provide estimates of causal associations between an exposure (such as metabolites) and outcome (such as dysglycemia) [15–17]. However, no study has yet utilised MR to establish the presence of casual associations between metabolites and measures of glycaemia at or before the 28 week of pregnancy in an ethnic-specific manner. The present study aimed to address this using the multi-ethnic Born in Bradford (BiB) cohort to identify ethnic-specific metabolic drivers of GDM.

## Methods

### Born in Bradford Cohort

#### Exposure data

BiB is a prospective longitudinal birth cohort that aimed to recruit all mothers receiving maternity care in the Bradford Royal Infirmary between 2007-2010 [18]. Bradford, a large city in the north of England, has high levels of deprivation and a large SA population, predominantly of Pakistani ancestry. A total of 12,453 women (mean maternal age 27.8) were recruited, 45% of which were of SA ancestry [18, 19]. All participants provided written consent and ethnical approval was obtained from the Bradford Research Ethnics Committee (ref07/H1302/112) [18].

Fasted plasma sample collection and high-throughput metabolite quantification by automated NMR (Nightingale Health©; Helsinki, Finland) has been previously described and validated to a high accuracy [2]. Briefly, samples were taken by trained phlebotomists from BiB participants (26-28 weeks’ gestation) and were processed in the absence of freeze-thaw cycles within 2.5 hours before storage at −80°C. One hundred and forty-six absolute measures of metabolites were included in the analysis following the removal of metabolites expressed as a percentage or ratio to minimise redundancy. In total, 10 overarching classes of metabolites were included in the analysis: lipoproteins (n=97), amino acids (n=9), apolipoproteins (n=2), cholesterols (n=8), fatty acids (n=8), glycerides and phospholipids (n=8), glycolysis related metabolites (n=4), ketone bodies (n=2), measures of fluid balance and inflammation (n=3) and measures of lipoprotein particle diameter (n=3). A full list of included metabolites can be found in **Supplementary Table 1**.

#### Outcome data

Participants were prior to GDM assessment and the 28^th^ week of pregnancy. In BiB, individuals were diagnosed with GDM if either their fasting glucose or if 2-hour post-load glucose concentration exceeded 6.1 mmol/L or 7.8 mmol/L following a 75g oral glucose tolerance test (OGTT) [20]. The OGTT was performed in the morning following an overnight fast. To maximise power, MR analyses were performed using continuous metabolite values and fasting glucose and 2-hour post glucose. Fasting glucose and 2-hour post glucose values were log normalised prior to analysis.

#### Metabolite data

Information on metabolite data preparation has been described in full elsewhere [9]. In brief, 11,480 blood samples were metabolically profiled from BiB, 54 of which were excluded due to failure of any one of five Nightingale© quality control measures leaving 11,426 samples prior to imputation. Missing data was imputed via multiple imputation using the missMDA package in R [21].

After combining with postprandial glucose data, 3,693 SA and 3,377 individuals whose samples were taken before the 28^th^ week of pregnancy were retained before outlier removal. Outliers were removed (those outside of 1.5 x IQR) for each metabolite in each ethnicity separately and metabolite values were normalised by taking the log, square root or normal score transformation (NST) as appropriate following the visual inspection of histograms and QQ plots. Following the removal of outliers, the number of individuals available for GWAS analysis of each metabolite was not the same (**Supplementary Table 2**) but was relatively consistent: for SAs the average sample size for each metabolite was 3622 (range 3472-3688), while for WEs it was 3301 (range 3158-3354). Information on gestational age at sample collection and parity was obtained from obstetric records. Ethnicity was self-reported or obtained from primary care records if missing. Individuals of a SA descent other than Pakistani were excluded from the analysis due to the small sample sizes of these populations.

#### Genetic data

Imputed genetic data were obtained from BiB. All samples were genotyped using two chips: the Infinium Global Sequencing Array-24 v.1 (GSA) (∼640K SNPs) and the Infinium CoreExome-24 v1.1 BeadChip (∼550K SNPs) [22]. Genetic data from the Illumina Global Sequencing Array (GSA) and Illumina CoreExome SNPs were combined. Where SNPs were missing in >5% of individuals, they were excluded [22]. When evaluating imputed data, the R^2^ value can be a measure of quality control as it reflects to the estimated proportion of genetic variation maintained in the imputed data. As a result, SNPs with an R^2^ <0.9 were excluded prior to analysis.

### GWAS analysis

Conventionally a GWAS assumes individuals are unrelated and the inclusion of related individuals can potentially lead to spurious associations [23, 24]. However, the removal of individuals from the BiB sample with close ancestry would substantially reduce the sample size. In addition, high rates of consanguinity in the SA stratum of the cohort makes relatedness difficult to assess [25]. As such, a GWAS mixed linear model association (MLMA) analysis was conducted in PLINK (version 1.9) that allowed for the inclusion of related individuals [24, 26–28]. MLMA models include a fixed effect, adjusted covariates, and an additional random effect comprised of a variance-covariance matrix that models the correlation (here relatedness) between individuals to be accounted for [24, 26]. GWAS MLMA models were implemented using the *GCTA* (Genome-wide Complex Trait Analysis) command line tool for each metabolite in both ethnicities [29]. To increase power, MLMA-loo (leave-one-out) analysis was utilised, preventing a SNP from being included in both the fixed and random effects concurrently, thereby avoiding double fitting [26]. MLMA models also included parity and principal components (PC) 1 and 2 to account for population stratification (**Supplementary Figure 1, Supplementary Table 3**). Gestational age, which showed little variation, was not included in the modelling (median gestational age SA = 184 days, IQR= 182-186.7, median gestational age WE = 184 days, IQR= 182-187). Genomic inflation factors (λ) were calculated for all models for a range of minor allele frequency (MAF) cut-offs (MAF <0.001, 0.001≤ MAF < 0.005, 0.005 ≤ MAF < 0.01, 0.01 ≤MAF < 0.05, 0.05 ≤ MAF < 0.1, and MAF ≥ 0.1) in order to minimise data loss while also minimising false positives. λ ≥ 1.1 was considered indicative of genomic inflation. [30, 31] A MAF cut-off of <0.05 was the least stringent cut-off found to reduce λ to ∼1 meaning this cut off was utilised in the analysis (**Supplementary Table 4**).

When a SNP was found to be associated with a metabolite value in only one ethnicity a fixed effect inverse-variance weighted meta-analysis was implemented to assess the heterogeneity (via the I^2^ statistic) of identified associations between ethnicities and to see if the SNP retained significance in a larger sample. Meta-analyses were conducted within the command-line tool METAL [32].

### One-sample MR

#### Genetic Instruments

One-Sample MR was conducted for all 146 metabolite values in both ethnic groups using SNPs identified as significant at a genome-wide suggestive level (p-value ≤ 1 x 10^-5^) in the GWAS. Metabolites were grouped into their overall classes and SNPs in each class were thinned by linkage disequilibrium (LD) (R^2^<0.2) via the NIH LDlink online tool (https://ldlink.nci.nih.gov) reducing the overlap of instruments in each class [33, 34]. For individuals of WE ancestry, all European (EUR) and South Asian (SAS) populations in LD link (software that utilises 1000 Genome data) were used to estimate LD due to the expected similarity in their LD structure allowing for an increase sample size and resultant improvement in the accuracy of LD estimates [35]. Similarity between 1000 Genome SA samples and BiB samples was assessed using principle components analysis (PCA) in PLINK (version 1.9) due to the fact that Pakistani samples from BiB originate from a different region of Pakistan (the Mirpur Region) from the 1000 Genome SA samples [27, 28]. This is of particular importance in SA populations as even geographically close populations can have differing allele frequencies due to differing Biraderi (‘Brotherhood’) membership between population subgroups. Biraderi membership is assigned at birth, is an indicator of male lineage as well as social-occupational status which largely governs partner choice and can result in higher levels of consanguinity in the population [22]. PCA plots were created using the *ggplot2* package in R studio (version 4.0.2) [36, 37]. No clear separation in SA BiB samples and SA 1000 Genome (1000G) samples was identified indicating that LD estimates obtained from 1000G was suitable for use in BiB (**Supplementary Figures 2-4**).

#### Analysis

Genetic Risk Scores (GRS) were created in PLINK (version 1.9) for each metabolite with each SNP receiving a weight based on its estimated effect size on the metabolite obtained from the GWAS [38]. One-sample MR was then performed by Two-Stage Least Squares regression (TSLS; *ivpack*, ivreg, and *AER* packages in R version 4.0.2) to obtain a causal estimate for the effect of each metabolite value on the log-normalised continuous measures of fasting glucose and 2-hour post glucose following a 75g oral glucose tolerance test (OGTT) [36, 39, 40]. Here, the level of a metabolite is regressed on its respective GRS and, subsequently, the outcome is regressed onto these fitted GRS-metabolite values in the second stage. All MR results have been reported according to STROBE-MR guidelines [41].

When significant associations were identified leave-one-out analysis was performed. For this, SNPs were removed sequentially from the instrument and changes to the effect estimate and F-statistic was assessed. If the exclusion of a SNP was found to alter either the effect estimates or F-statistic (through the visualisation of forest plots) it is possible that the SNP is influencing the outcome via an alternative pathway to other SNPs, potentially highlighting a violation of the 2^nd^ or 3^rd^ MR assumption. To further test for violations of these assumptions, included SNPs were searched for in both the Phenoscanner and GWAS Catalog databases to identify previously identified associations [42–44]. In both databases a p-value ≤ 1 x 10^-5^ was interpreted as indicative of an additional association. Differences between MR and linear regression results were also evaluated via the Wu-Hausman statistic to assess deviation of the instrumental variable estimate from the ordinary least squares (OLS) estimate [45]. Deviations in these two measures can indicate either confounding in the OLS estimate (indicating a need for MR) or violations of the MR assumptions due to pleiotropy.

#### Post-hoc power analyses

For metabolites that associated with a measure of postprandial glucose in only one ethnic group, *post-hoc* power analyses were performed using the mRnd CNS genomics tool (https://shiny.cnsgenomics.com/mRnd/) to assess whether the absence of an association in the alternate ethnicity was due to limited power [46]. Observational and ‘true’ associations required by the tool were obtained by performing linear regression of the outcome on the metabolite and obtaining unadjusted and adjusted estimates (adjusted for maternal age (years), BMI (continuous), smoking status, multiple pregnancy, parity, and gestational age) respectively. Due to the *post-hoc* nature of this analysis, additional power analyses could be conducted assuming the MR estimate to be the true causal effect in the MR calculation. This analysis was performed in the non-significant population for each metabolite associated in only one ethnicity. If power was found to be adequate (80%) at the 5% level (α = 0.05) power was also assessed at the 1% level (α = 0.01).

## Results

### GWAS of Metabolite Measures

A total of 6184 SNPs were associated with at least one metabolite in WEs at the suggestive level (1 x 10^-5^), with 2616 (42.3%) SNPs being associated with a single metabolite measure. However, no SNPs were identified below the genome-wide significant level (p-value <5 x 10^-8^) in WEs.

Fewer SNPs were identified at the suggestive level in SAs, with 3685 SNPs SNP-metabolite associations in total, of which 1544 (41.9%) SNPs being associated with only one metabolite measure. SNP associations were identified for 138/146 (94.5%) metabolite exposures in SAs (**Supplementary Table 5**). No SNP was identified as being associated at the suggestive level in both ancestries, although shared genomic regions were identified between ancestries (**Supplemental Excel**).

To evaluate the possibility of shared genomic predictors of metabolites, a pooled meta-analysis of effect estimates in both ethnicities was performed. For 90 metabolite values, no associations were found to exceed the genome-wide suggestive level (p<10^-5^) following meta-analyses of both ethnicities. SNP associations were identified at the suggestive level for four metabolite measures (concentration of XL-HDL, total lipids in M-VLDL, mean density of VLDL and citrate) despite these differing in direction of effects in SAs and WEs. In addition, 4 SNPs were associated with alanine, despite these SNPs initially being associated with alanine only in the SA population. These SNPs (rs12256633, rs17121228, rs7096521, rs12240368) are all found on chromosome 10 in the receptor gene *SORCS1* and have not been associated with alanine levels previously [47, 48].

### MR results

After LD thinning, genetic instruments were available for all 146 metabolites in WEs and for 136/146 (93.2%) metabolites in SAs. In WEs, 1040 SNPs were retained following LD thinning including 423 (40.67%) that were unique to an individual metabolite. Fewer SNPs were identified in SAs, where 383 SNPS remained after LD thinning, 195 (50.9%) of which were unique to a single metabolite. (**Supplementary Table 5**). 2.7% of included genetic instruments (4 metabolites) and 12.5% (9 metabolites) of included genetic instruments in WEs and SA respectively had an F-statistic < 10, indicating that most instruments were at low risk of weak instrument bias [49]. The average F-statistic for WEs instruments was 72.4, while in SAs it was considerably lower at 26.7.

#### White Europeans

Two metabolite values, leucine and mean density of HDL lipoproteins (HDL_D), were associated with both fasting glucose and 2-hour post glucose (**Table 1**). Specifically, a 1mmol/L increase in blood leucine associated with lower fasting glucose (−0.193 mmol/L, 95% CI −0.069, −0.319) and 2-hour post glucose (−0.443 mmol/L, 95% CI −0.113, −0.774). Likewise, a 1nm increase in mean diameter of HDL associated with lower fasting glucose (−0.082 mmol/L, 95% CI 0.026, 0.138) and 2-hour post glucose (−0.191, 95% CI 0.043, 0.339 mmol/L) **(Supplementary Figures 4-5**). No other metabolites were associated with both measures of glucose in WEs.

**Table 1:**
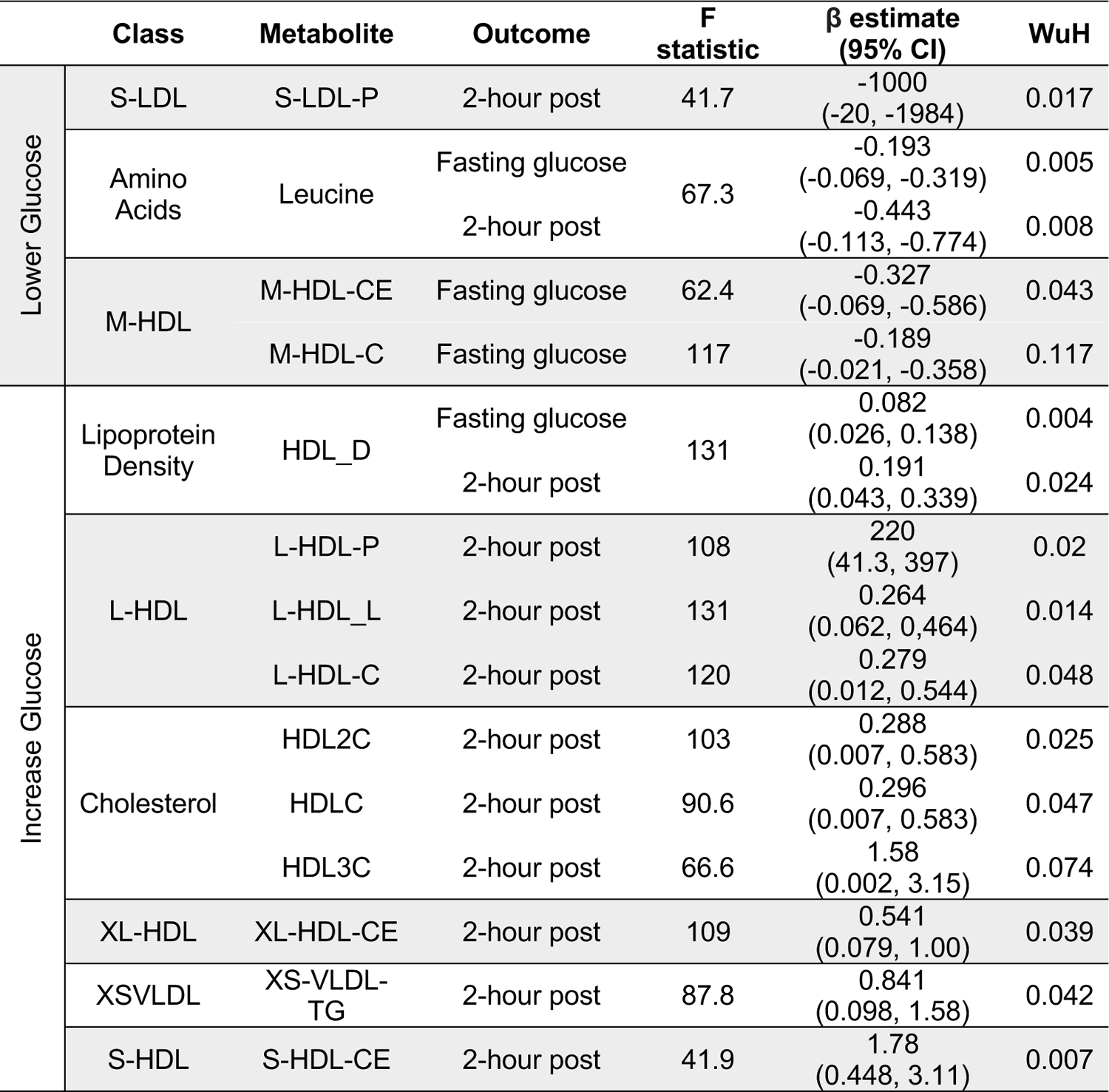
Significant MR results in white Europeans. Glucose measures are expressed as mmol/L. HDL_D: mean diameter of HDLs (nm). HDLC: total cholesterol in HDL (mmol/L). HDL2C: total cholesterol in HDL2 (mmol/L). HDL3C: total cholesterol in HDL3 (mmol/L). L-HDL-C: total cholesterols in L-HDL (mmol/L). L-HDL_L: total lipids in L-HDL (mol/L). L-HDL-P: concentration of L-HDL (mol/L). M-HDL-C: total cholesterol in M-HDL (mmol/L). M-HDL-CE: cholesterol esters in M-HDL (mmol/L). S-HDL-CE: cholesterol esters in S-HDL (mmol/L). S-LDL-P: concentration of S-LDL (mol/L). XL-HDL-CE: cholesterol esters in XL-HDL (mmol/L). XS-VLDL-TG: triglycerides in XSVLDL (mmol/L). WuH: Wu-Hausman p-value.

For fasting glucose, an increase of 1mmol/L total cholesterol in M-HDL (M-HDL-C) and cholesterol esters in M-HDL (M-HDL-CE) were associated with lower fasting glucose measures (−0.189 mmol/L, 95% CI −0.021, −0.358, and −0.327 mmol/L, 95% CI −0.069, −0.586 respectively). For 2-hr post-glucose, 8 metabolite values were positively associated with this (HDLC, HDL2C, HDL3C, triglycerides in XS-VLDL, cholesterol esters in XL-HDL, total concentration of L-HDL, total lipids in L-HDL and cholesterol esters in S-HDL) and one (total concentration of S-LDL) was negatively associated (**Table 1**). Cholesterol metabolites, measures of total cholesterols in lipoproteins and total cholesterols in lipoproteins were the most common types of metabolite class to be associated with postprandial glucose in WEs, with leucine being the only amino acid identified. Wu-Hausman p-values < 0.05 indicate deviations of the instrumental variable estimate from the OLS estimate (**Table 1**).

#### White Europeans: Sensitivity analyses

For 6 of 13 metabolite values, leave-out one analyses maintained significance (P≤ 0.05) indicating that no individual SNP was driving the identified associations in WEs: leucine, mean diameter of HDL, total lipids in L-HDL, cholesterol esters in S-HDL and cholesterol esters in M-HDL (**Supplementary Figure 7**). For the remaining 8 metabolites, β values were consistent across leave-one-out analyses although not all associations remained significant. Additionally, for 12/13 metabolites (all but M-HDL-CE), the F-statistic did not substantially differ through the exclusion of individual SNPs from the instruments, which suggests they were not substantially driven by a single SNP (**Supplementary Figure 8**). The exception to this was cholesterol esters in M-HDL, where the exclusion of rs2138011 or rs739018 increased the F-statistic.

Three of the metabolites (leucine, L-HDL-L, L-HDL-C) that were associated with postprandial glucose in WEs included a SNP that previous studies have associated (p ≤ 1 x 10^-5^) with at least one potential confounder (BMI, hypertension or waist circumference) (**Supplementary Table 7**). The removal of these SNPs from the instrument did not impact the significance of the associations identified for leucine or L-HDL-L (**Table 2**). However, for L-HDL-C instrument, the exclusion of two SNPs (rs5576825 and rs6811162) previously associated with a potential confounder (waist circumference and hypertension respectively) resulted in non-significant association between L-HDL-C and 2-hour post glucose. Importantly, for both SNPs, it is conceivable that the confounders could reside on their causal pathway (i.e., vertically pleiotropic, where L-HDL-C effects 2-hr post-prandial glucose through its effect on weight gain) rather than be in horizontal pleiotropy and may, therefore, violate the 2^nd^ MR assumption [49].

**Table 2:**
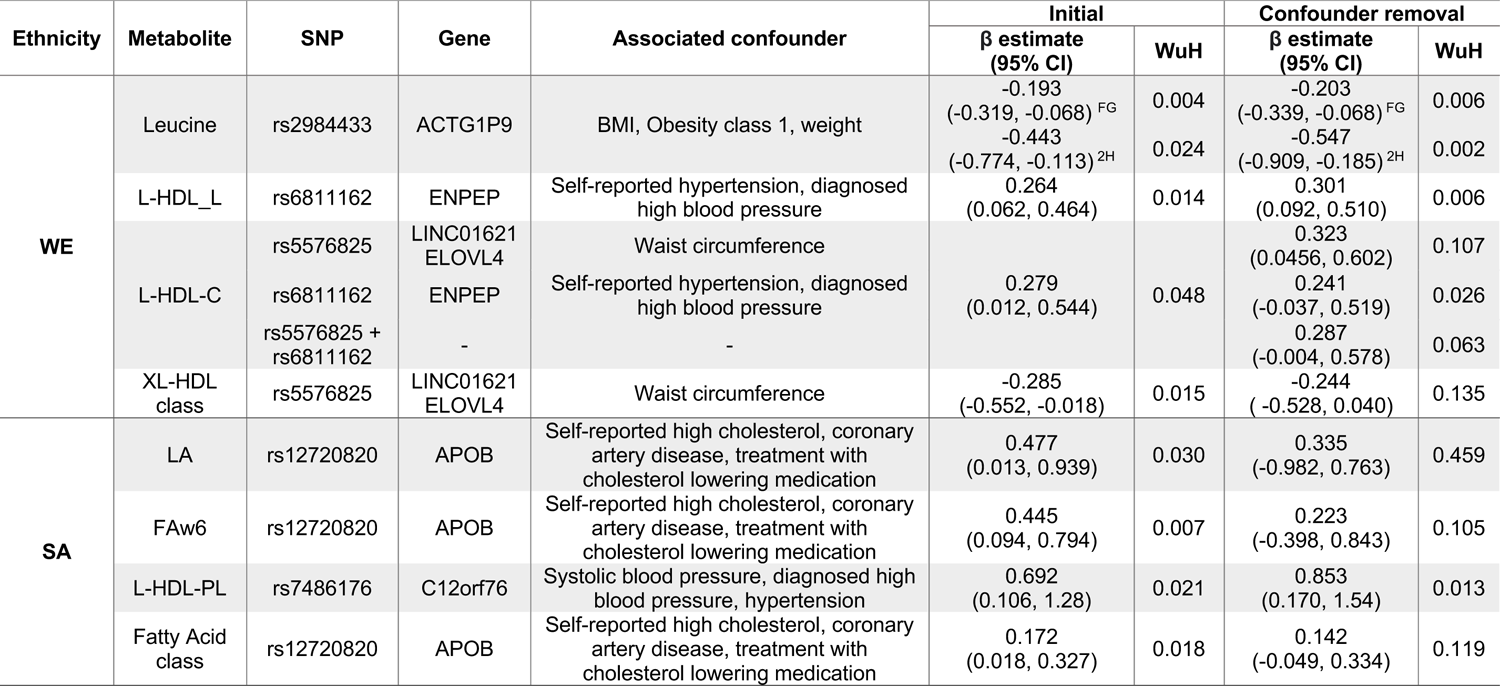
Removal of potentially pleiotropic SNPS. 2H: 2-hour post glucose. FAw6: Total n-6 fatty acids. FG: fasting glucose. LA: 18:2 linoleic acid (mmol/L). L-HDL-C: total cholesterols in L-HDL (mmol/L). L-HDL_L: total lipids in L-HDL (mmol/L). L-HDL-PL: phospholipids in L-HDL (mmol/L). SA: South Asian. WE: White European. WUH: Wu-Hausman p-value.

#### South Asians

No metabolite was associated with both fasting glucose and 2-hour post glucose in SAs. Although, for fasting glucose, a 1 mmol/L increase in either total FAw3 or S-HDL-C was associated with an increase of fasting glucose by 0.432 mmol/L (95% CI 0.063 – 0.798) and 1 mmol/L (95% CI 0.116 – 1.882) respectively. No metabolite associated with a decrease in fasting glucose in SAs (**Supplementary Figures 4-5**).

Nine metabolites associated with 2-hour post glucose levels in SAs. Of these, 4 metabolites, LA, FAw6, total lipids in M-VLDL (M-VLDL-L) and total phospholipids in L-HDL (L-HDL-PL), associated with an increase in with 2-hour post glucose, with the largest effect being identified for L-HDL-PL. Specifically, a 1mmol/L increase in L-HDL-PL associated with a 0.692 mmol/L increase (95% CI 0.106 - 1.280) in 2-hour post glucose. A further 5 additional metabolites were associated with a decrease in 2-hour post glucose: concentration of L-LDL (L-LDL-P), total cholesterols in IDL (IDL-C), cholesterol esters in IDL (IDL-CE) concentration, total cholesterols in IDL (IDL-C), total lipids in small S-LDL (S-LDL-L), and total lipids in small S-HDL (S-HDL-L). The largest decrease in 2-hour post glucose was observed for L-LDL-P where a 1mmol/L increase in L-LDL-P associated with a 3.86 mmol/L decrease (95% 0.467 - 7.27) in 2-hour post glucose levels (**Table 3**).

**Table 3:** Significant MR results in South Asians Glucose measures are expressed as mmol/L. FAw3: total n-3 fatty acids. FAw6: total n-6 fatty acids. IDL-C: total cholesterols in LDL (mmol/L). IDL-CE: cholesterol esters in LDL (mmol/L). LA: 18:2 Linoleic Acid (mmol/L). LDL_P: concentration of LDL particles (mol/L). L-HDL-PL: phospholipids in L-HDL (mmol/L). M-VLDL-L: total lipids in M-VLDL (mmol/L). S-HDL-C: total cholesterols in S-HDL (mmol/L). S-HDL-L: total lipids in S-HDL (mmol/L). S-LDL-L: total lipids in S-LDL (mmol/L). WuH: Wu-Hausman p-value.

| | Class | Metabolite | Outcome | F statistic | $\beta$ estimate (95% CI) | WuH |
| --- | --- | --- | --- | --- | --- | --- |
| Lowers Glucose | L-LDL | LDL-P | 2-hour post | 11.7 | -2238 (-4828 -193.7) | 0.024 |
|  | S-LDL | SLDL-L | 2-hour post | 11.1 | -3.86 (0.467, -7.27) | 0.015 |
|  | IDL | IDL-C | 2-hour post | 10.9 | -1.19 (-0.12, -2.27) | 0.021 |
|  |  | IDL-CE | 2-hour post | 11.8 | -1.34 (-0.144, -2.55) | 0.023 |
|  | S-HDL | S-HDL-L | 2-hour post | 11.4 | -1.23 (-0.137, -2.32) | 0.012 |
| Increase Glucose | Fatty Acids | LA | 2-hour post | 20.9 | 0.477 (0.013, 0.939) | 0.030 |
|  |  | FAw3 | Fasting glucose | 10 | 0.432 (0.063, 0.798) | 0.008 |
|  |  | FAw6 | 2-hour post | 33.4 | 0.445 (0.094, 0.794) | 0.007 |
|  | M-VLDL | M-VLDL-L | 2-hour post | 68.4 | 0.046 (0.009, 0.083) | 0.008 |
|  | L-HDL | L-HDL-PL | 2-hour post | 43.3 | 0.692 (0.106, 1.28) | 0.021 |
|  | S-HDL | S-HDL-C | Fasting glucose | 22 | 1 (0.116, 1.882) | 0.012 |

Fatty acids were the class of metabolites most frequently associated with postprandial glucose in SAs. All three fatty acids (LA, FAw3 and FAw6) associations identified in SAs were of similar magnitude: a 1 mmol/l increase of FAw3 associated with a +0.4 mmol/l increase in fasting glucose or and a 1 mmol/ increase od FAw6 and LA associated with a +0.4 mmol/l increase of 2-hour post glucose.

No metabolite found to be associated with postprandial glucose measures in WEs was found to be associated with postprandial glucose in SAs. However, in both populations, members of the S-HDL and L-HDL class were found to be associated with increased postprandial glucose.

#### South Asians: Sensitivity analyses

Six instruments in SAs were comprised of a single SNP meaning it was not possible to perform a leave-one-out analysis for these metabolites. For the remaining 5 metabolites, associations were consistent across each leave-one-out analyses (**Supplementary Figure 9**). Likewise, no large differences in F-statistics following the removal of individual SNPs were identified (**Supplementary Figure 10**).

Just as in WEs, 3 metabolites identified in SAs included SNPs associated with cholesterol or hypertension, which are potential confounders of the association between metabolites and dysglycemia (**Supplementary Table 6**). Significance was maintained following the removal of SNP rs7486176 (found within the *C12orf76* gene) from the total phospholipids in L-HDL instrument. For the LA and FAw6 exposures, the removal of SNP rs12720820 (found within the *APOB* gene) resulted in a non-significant association indicating that this SNP was the main driver of the identified association (**Table 2**). In leave-one-out analyses, the removal of SNP rs58865405 from the FAw6 instrument resulted in non-significance, although the biological role of this SNP remains unknown.

#### *Post-hoc* analysis: Analysis of metabolite classes

Numerous SNPs were found to be associated with more than one metabolite measure, particularly for metabolites in the same metabolite class (**Supplementary Figures 11-12**). This was anticipated since many metabolomic pathways are biologically intertwined. To minimise the risk of violation of the third MR assumption (that the genetic instrument must only influence the outcome via the exposure and not via an alternative biological pathway) [15], the collective effect of an entire class of metabolites on postprandial glucose measures was examined. A composite score for each metabolite class was created by placing all metabolites in a single class (e.g. all LDL metabolites), conducting a PCA and extracting PC1. This was only possible for 20 classes in WEs and 21 classes in SAs that had > 2 metabolites and ≥70% of the class variation was explained by PC1 (**Supplementary Table 7**). To assess the impact of outliers on PCA, outliers were defined and removed based on two cut-offs: standard (1.5 X IQR from the median) and stringent (3 x IQR from the median). For all classes, PC1 and PC2 scores were comparable after removal of both types of outliers so only 3xIQR outliers were removed prior to analyses (**Supplementary Table 8**).

138 SNPS remained after LD thinning in WEs, 87 (63.04%) of which were unique to a single metabolite class exposure. 19/20 (95%) of the metabolite classes examined in WEs had an F-statistic ≥10 (the only exception being the MHDL class). 54 SNPS remained after LD thinning in SAs, 42 (77.78%) of which were unique to a single metabolite class. Despite the lower number of SNPs identified in SAs,17/20 (85%) of the metabolite classes examined had a F-statistic ≥10 indicating that instrument strength was still sufficient, for all classes except for the non-branched amino acids, LLDL class, and all VLDL classes. On average, the mean F-statistic of metabolite class instruments was 19.83%. On average genetic instruments of composite measures of the metabolite classes were weaker than the instruments for individual metabolites in both WEs and SAs (**Supplementary Figure 13**).

In WEs, 4 metabolite classes were associated with a glucose measure: S-LDLs associated with fasting and 2-hour post glucose; M-LDL and all LDLS (i.e. the collective grouping of HDLs, MDLs and SDLs) were associated with fasting glucose, and XL-HDLs were associated with 2-hour post glucose (**Table 4**). In SAs, the fatty acid metabolite class were associated with 2-hour post glucose levels. No other associations were identified in SAs.

**Table 4:** Significant MR results from the analysis of metabolite classes Glucose measures are expressed as mmol/L. WuH: Wu-Hausman p-value.

| Ethnicity | Metabolite class | Outcome | F statistic | $\beta$ estimate (95% CI) | WUH |
| --- | --- | --- | --- | --- | --- |
| White European | XL-HDL | 2-hour post glucose | 96.2 | -0.285<br>(-0.018, -0.552) | 0.015 |
|  | M-LDL | Fasting glucose | 95.5 | -0.048<br>(-0.004, -0.091) | 0.024 |
|  | S-LDL | Fasting glucose | 98 | 0.084<br>(0.007, 0.162) | 0.024 |
|  |  | 2-hour post glucose | 98 | -0.249<br>(-0.016, -0.482) | 0.066 |
|  | All LDL | Fasting glucose | 95 | 0.038<br>(-0.005, -0.068) | 0.016 |
| South Asian | Fatty Acids | 2-hour post glucose | 32.8 | 0.172<br>(0.018, 0.327) | 0.0184 |

As with the sensitivity analyses of individual metabolites, the removal of individual SNPs was not found to greatly impact the F-statistic of most instruments (**Supplementary Figures 14-15**). However, for the fatty acid metabolites class, the exclusion of rs12720820 or rs7159441, and for ‘XL-HDL’, the removal of rs55768285, resulted in non-significant associations, suggesting that these SNPSs are key drivers of the association. (**Table 4, Supplementary Table 9**).

#### Power analysis

R^2^ values were consistently lower in the ethnic group where an effect was not detected but all genetic instruments for the metabolite values had an F-statistic ≥ 10, indicating that weak instrument bias was not responsible for the absence of significant effects. Where an association was identified in one ethnic group but not another, to determine whether the absence of an association was potentially due a lack of power in the other ethnicity rather than an ethnic-specific effect, *post-hoc* power analyses were performed.

When using MR estimates as an estimate for the true causal effect both the analyses of FAw3 and the overall fatty acid class in WEs were adequately powered to detect the observed MR effect in both populations. Therefore, the absence of an effect of FAw3 in WEs is unlikely due to inadequate power (**Supplementary Table 10**). The analysis of HDL2C and HDL3C in SAs was also sufficiently powered to detect the observed MR effect in WEs.

## Discussion

This study has identified ethnically distinct associations between a range of metabolites and postprandial glucose measures taken during pregnancy in SAs and WEs, with notably no shared associations were identified. Fourteen metabolites were found to be associated with postprandial glucose measures in WEs. Whereas, a distinct set of 11 metabolites were associated in SAs. In WEs, cholesterols and lipoproteins were the metabolite classes associated with postprandial glucose measures, while in SAs fatty acids were the most commonly associated.

Furthermore, through an extensive GWAS of metabolites, this study identified novel genome-wide significant associations in relation to acetate (1 SNP, rs10945476) and tyrosine (15 SNPs, all on chromosome 17) in SAs. No previous associations have been identified for SNP rs10945476, found within the non-coding transcript gene *PRDM15* in relation to acetate or any other exposure.

Interestingly, 3 of the 15 SNPs associated with tyrosine are found in a transmembrane transporter gene, *SLC13A2*. Moreover, an additional 10 of the newly identified 15 SNPs associated with tyrosine were found in the *FOXN* gene, a transcription factor that has previously been identified to be associated with ceramide levels (a lipid metabolite) in a GWAS from a Chinese cohort [50]. Moreover, ceramide has been shown to induce tyrosine phosphorylation in membrane proteins meaning it is plausible that a gene associated with ceramide is also associated with tyrosine levels in an Asian population [51]. Interestingly, ceramide has been proposed as a mediator of the interaction between saturated fat and insulin resistance and has been associated with T2D and cardiovascular disease [52]. To the best of our knowledge tyrosine levels have not previously been associated with either *FOXN SLC13A2*. The remaining 2 of the 15 SNPs identified as being associated with tyrosine in SAs, are currently not in any known genes. All 15 SNPs identified as being associated with tyrosine in SAs are in LD with each other in 1000G SA populations (all R^2^ ≥ 0.38).

### Identified associations in WEs

#### Leucine

Branched chain amino acids (BCAAs), including leucine, are predominantly metabolised in skeletal muscles where they regulate protein synthesis and mitochondrial functions [53]. In addition, BCAAs are hormonal signalling regulators and are expected to module insulin resistance (IR) through increasing insulin secretion in human pancreatic β-cells [54, 55]. Our study found leucine to be negatively associated with both fasting glucose and 2-hour post glucose levels during pregnancy in WEs; with 1 mmol/L of leucine associated with a decrease of 0.193 mmol/L in fasting glucose and 0.327 mmol/L 2-hour post glucose respectively. Although few studies have investigated the role of leucine in glucose regulation during pregnancy, interestingly the ratio of leucine/isoleucine was similarly found associated with reductions in fasting glucose in the HAPO study, a multi-ethnic cohort of pregnant women of Afro-Caribbean, Mexican American, Northern European, and Thai ancestry [56]. Common dietary sources of leucine include meat products and cheese, with smaller amounts also being present in other dairy products (such as dairy and yoghurt), fish and in certain legumes and nuts, such as dried raw broad beans and pine nuts [53]. Hence, dietary interventions aimed at increasing leucine levels during pregnancy, possibly through a dietary intervention promoting the consumption of lean animal protein, low-fat dairy and nuts, may help improve pregnancy hyperglycaemia in WEs.

#### Cholesterols

HDL cholesterol is colloquially described as ‘good cholesterol’ due to its role in the removal of cholesterols from atherosclerotic plaque, thereby reducing an individual’s risk of CVD [57]. Furthermore, low HDL levels have commonly been associated with diabetes in humans, with HDL shown to increase insulin secretion and β-cell survival [58, 59]. We identified four associations between HDL cholesterol and postprandial glucose measures in WEs. Herein, 1 mmol/L increase in S-HDL-CE confers a 1.78 mmol/L increase (95% CI 0.49 – 3.11) in 2-hour post glucose. This is consistent with previous evidence from a Finnish sample of overweight and obese women where cholesterol esters in S-HDL were higher in the serum samples of GDM cases at ∼14 weeks gestation [60]. Discrepancies in the direct effect of HDL cholesterol on dysglycemia have also been identified in the genetic literature [59], with a recent review highlighting that while a genetic study utilising linear relation analysis did find HDLs to have a protective effects against T2D (n cases = 2,447) [61], the same effect was not been replicated in an MR setting (n cases= 47,627) [62]. When considering LDL cholesterols, only S-LDLs was found to be significantly associated with a postprandial glucose measure (fasting glucose) in WEs. Additionally, in our composite analysis of metabolite classes, S-LDLs were associated with fasting glucose and 2-hour post glucose in WEs, whereas the M-LDL and all LDL (a combined measure of S-LDLs, M-LDLs and HDLs) classes were associated with fasting glucose. Unfortunately, because composite scores were comprised of PC1 coordinates the direction of effect of these associations could not be evaluated. To our knowledge, no previous study has conducted an MR of metabolites on dysglycemic predictors of GDM.

#### Triglycerides

Triglycerides are an abundant class of lipid particles found in the blood, originating from either from the consumption of dietary fats or as a result of hepatic metabolism [63, 64]. Once in the blood, triglycerides can be incorporated into HDL and LDL cholesterol particles. In addition to dietary triglyceride consumption, dietary fatty acids can be converted into triglycerides before they enter circulation, highlighting the complex relationship between triglyceride, cholesterol, and fatty acid levels [63].

Our results suggest triglycerides in XSVLDL (XS-VLDL-TG) associate with increased 2-hour post glucose (0.841 mmol/L) in WEs. In agreement with these findings, increased triglycerides in XSVLDL levels have also previously been associated with increased likelihood of GDM in a Finnish population [60]. No other triglyceride was found to be associated with in WEs. One explanation no additional associations were detected could be due to the average BMI of the WEs in BiB. For example, an analysis of a prospective Irish cohort (∼94% WE) found that triglyceride levels were only associated with GDM in obese individuals, a higher average BMI than that observed in the BiB cohort [65]. Further confirmation of these findings of increased triglycerides in XS-VLDLs would suggest that this association is, at least in part, responsible for the identified associations between diets high in fats and increased prevalence of GDM in WEs [11].

### Identified associations in SAs

#### Fatty acids

Polyunsaturated acids (PUFAs) are consumed in the diet and can be converted to long-chain PUFAs (LC-PUFAs) through a process of desaturation and elongation reactions that predominately occur in the liver [66]. Changes in dietary patterns can have a large impact on fatty acid composition in the body and with-it disease risk. For example, a western dietary pattern, which has high levels of n-6 fatty acids, has been associated with GDM risk [11, 67]. In a cohort of Chinese adults, total n-6 fatty acids and 18:2 n-6 levels at baseline in venous blood samples were both found to associate with an increased risk of T2D after ∼8 years of follow up, while increased n-3 fatty acid levels were protective [68]. However, a recent two-sample MR suggested only a negligible effect of n-6 PUFA synthesis on T2D in a predominantly WE cohort [69]. Moreover, the relationship between n-3, n-6, n-9 fatty acids and GDM remains inconclusive, and in a recent (2021) systematic review none of the identified studies (n=15) was conducted in a SA population [67], highlighting the need for more studies exploring the role of fatty acids in GDM development in Asians [70].

This study provides evidence of an association between LA and total FAw6 levels and an increase in 2-hour post glucose levels during pregnancy in SAs. In addition, the fatty acid class associated with 2-hour post glucose in SAs. Through a leave-out-one sensitivity analyses for the FAw6 and LA instruments, the removal of the SNP rs12720820 (found within the *APOB* gene) resulted in non-significant associations for both exposures, indicating that this rs12720820 was the largest contributor to the identified associations and has previously been associated with cholesterol levels and the use of cholesterol lowering drugs [71]. Interestingly, FAw3 also associated with increased 2-hour post glucose in SAs; however, with only one SNP potential pleiotropy could not be explored.

It is well established that fatty acid profiles can impact blood cholesterol levels [72–74]. In addition, increased dietary cholesterol has previously been associated with an increased risk of GDM in a systematic review of observational studies [75]. Taken together, our data confirm that fatty acids and cholesterol metabolites are in vertically pleiotropy and are likely impacting gestational dysglycemia via the same causal pathway. Unlike horizontal pleiotropy, vertical pleiotropy does not result in a violation of the 2^nd^ MR assumption as cholesterol is not acting as a confounder, meaning MR estimates are still valid (**Figure 1**). Furthermore, it is also possible that this interaction between fatty acids and cholesterols may be ethnic-specific due to the absence of associations identified between fatty acids and postprandial glucose measures in WEs. In addition to possible variations in cholesterol metabolism, it is plausible that variations in fatty acid synthesis are also partially responsible for the increased GDM risk experienced by SAs. For example, variants within the *FADS* genes impact LC-PUFA conversion [76, 77]. Current evidence suggests that SAs are likely to synthesise LC-PUFA more quickly than WEs, which could contribute to elevated risk of prolonged exposure to elevated LC-PUFA levels (namely, w6) and risk of dysglycemia [76, 77]. If these ethnic differences in fatty acid metabolism are confirmed to be linked to disease risk, it would aid in the development of tailored GDM prevention strategies that focus on modifying fatty acid profiles in an ethnic-specific manner.

**Figure 1:**
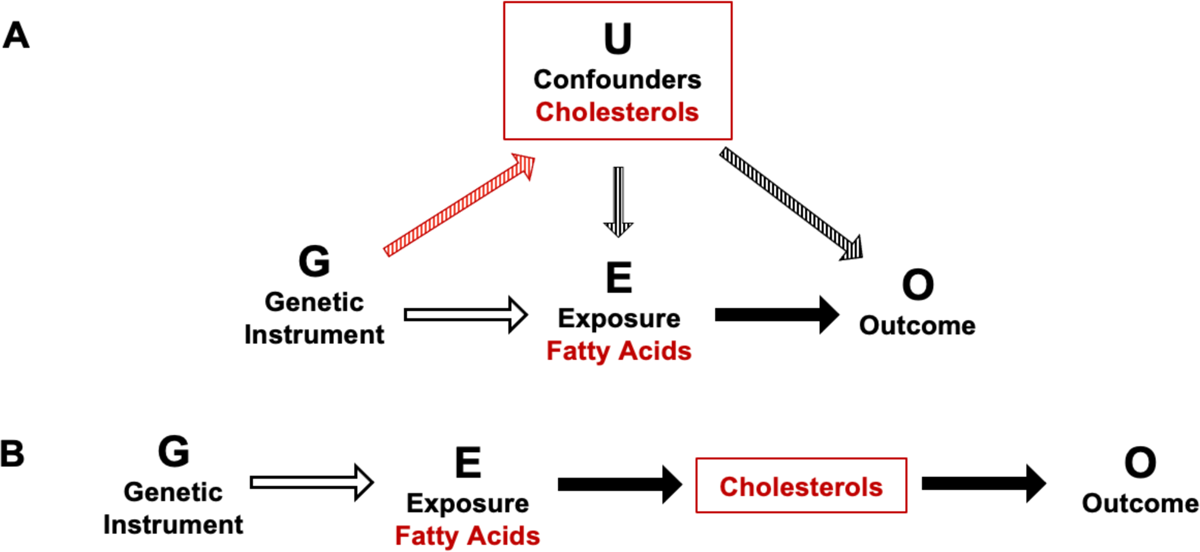
Schematic of potential horizontal and vertical pleiotropy in relation to fatty acid and cholesterol metabolites and postprandial glucose measures. **A:** Illustration of horizontal pleiotropy. **B:** Illustration of vertical pleiotropy. Vertical pleiotropy does not result in a violation of the 2^nd^ MR assumption.

The analyses found no association between triglycerides and dysglycemia in SAs. This agrees with a recent meta-analysis that concluded, although triglyceride levels associated with likelihood of GDM (I^2^ ≥ 84%) [78], after stratification by culture/geographical location, they found no association between triglyceride levels and likelihood of GDM. The reasons for this are unclear but it has also been shown that SAs have a higher prevalence of hypertriglyceridemia than WEs and at lower BMI levels, meaning it is possible that the difference in triglyceride levels and in SA GDM cases and controls is less pronounced than in WEs [79].

### Strengths and limitations

This analysis has several strengths. Firstly, this study involved a large and comprehensive panel of metabolites allowing for the relationships between metabolites and postprandial glucose to be thoroughly investigated. Secondly, this is the first MR study to investigate dysglycemia during pregnancy while also being one of the few MR studies to be conducted in a SA population. Finally, through leave-one-out analyses and the searching of both Phenoscanner and GWAS Catalog databases, violations of the 2^nd^ and 3^rd^ MR assumptions were thoroughly investigated meaning that it was possible to conclude that identified associations may not be subjected to horizontal pleiotropy and that identified causal associations are valid due to the absence of detectable violations of the MR assumptions.

Nonetheless, this study has some limitations. Metabolites are highly correlated meaning it is not possible to confidently interpret that an individual metabolite is independently associated with a postprandial outcome measure. To account for this limitation MR analyses were performed on composite measures of each metabolite class (when PC1 explained ≥70% of the variation in the metabolite class) in order to assess the overall impact of each metabolite class on pregnancy dysglycemia. Furthermore, MR also assumes the level of genetically conferred exposure from conception to the time of measurement is constant, which is unknown when studying metabolites. The limited sample size of this study also means that further adjustment could not be made at the GWAS stage since missing data in certain variables would further reduce sample size (eg, age) and some associations may have been underpowered to detect an effect. However, a *post hoc* power analyses found that for some metabolite values significant effects only identified in one ethnicity were possible to detect in the alternate ethnicity. In addition, some genetic instruments included only one SNP meaning it was not possible to evaluate the impact of pleiotropy for any identified associations involving these instruments. Finally, due to limitations in data availability in SAs a two-sample MR could not be conducted meaning it was not possible to assess the generalisability of these findings.

## Conclusions

The presence of causal relationships between a comprehensive set of metabolites and postprandial glucose measures (fasting glucose and 2-hour post glucose) in mid-pregnancy has been established in a UK SA and WE population. This study has found a range of metabolite values to be associated with postprandial glucose measures in WEs and high-risk SA women, although more associations were identified in WEs despite these individuals being at lower risk of GDM. In high-risk SA women, total n-6 fatty acids and the n-6 fatty acid, LA appear to increase postprandial glucose levels suggesting that fatty acids may be responsible for a large proportion of metabolically driven risk for GDM experienced by this population. Future work in a larger sample (potentially using a two-sample MR) and a larger panel of metabolites is needed to investigate our findings and hypotheses more closely, ideally over the course of a pregnancy in order to aid in GDM prevention in this high-risk population.

## Data Availability

All data produced in the present work are contained in the manuscript.

https://borninbradford.nhs.uk/research/

## Acknowledgements

Born in Bradford is only possible because of the enthusiasm and commitment of the children and parents in BiB. We are grateful to all the participants, health professionals, schools and researchers who have made Born in Bradford happen. This work was undertaken on ARC4, part of the High Performance Computing facilities at the University of Leeds, UK.

---

**Supplementary Figure 1:**
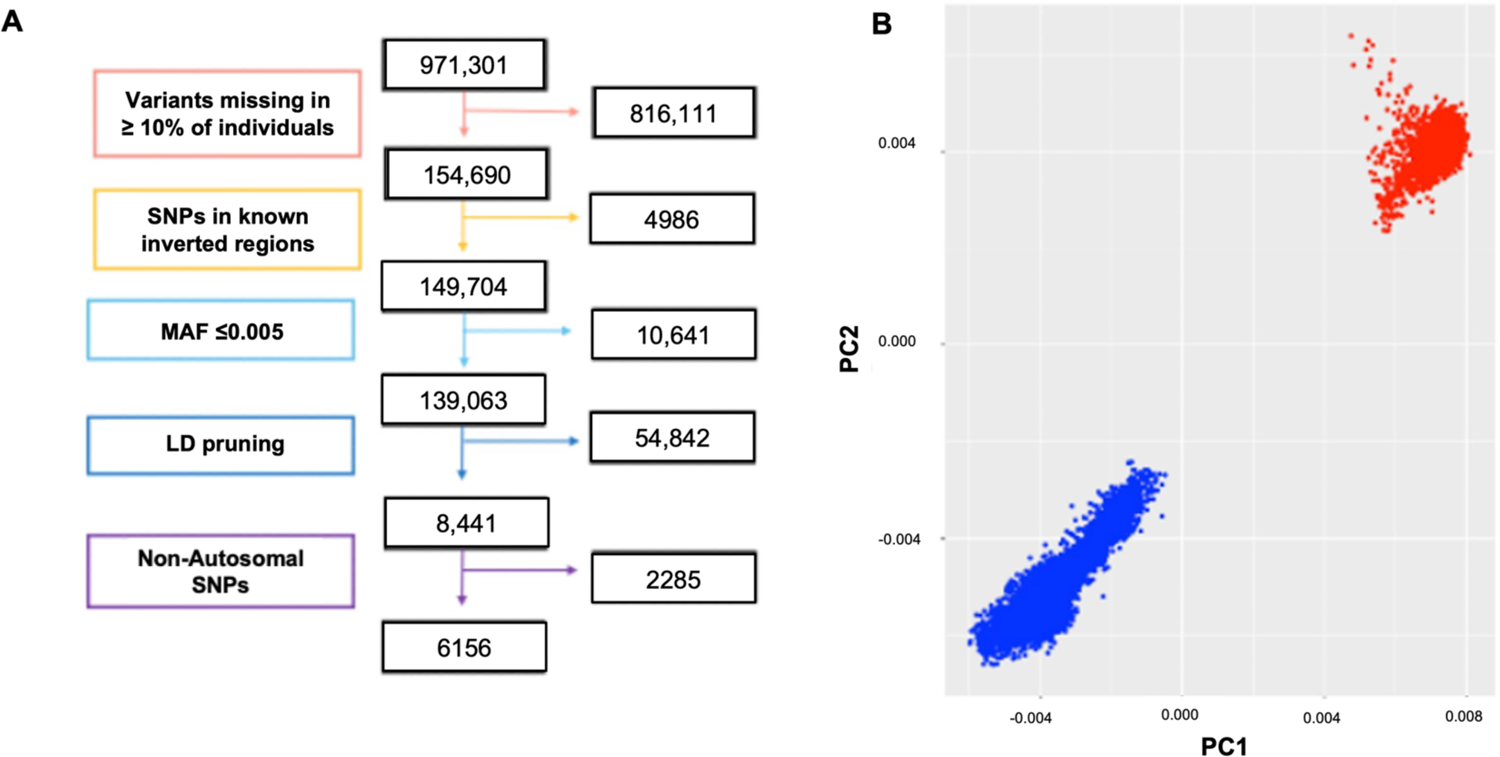
PCA to account for population stratification. A: Schematic of data processing steps taken prior to PCA of genetic data. R^2^ cut-off of 0.3 and 50 variant windows were utilised for LD pruning. MAF: minor allele frequency. B: PCA plot of BiB genotype data calculated to account for population stratification. Blue: South Asians. Red: white Europeans.

**Supplementary Figure 2:**
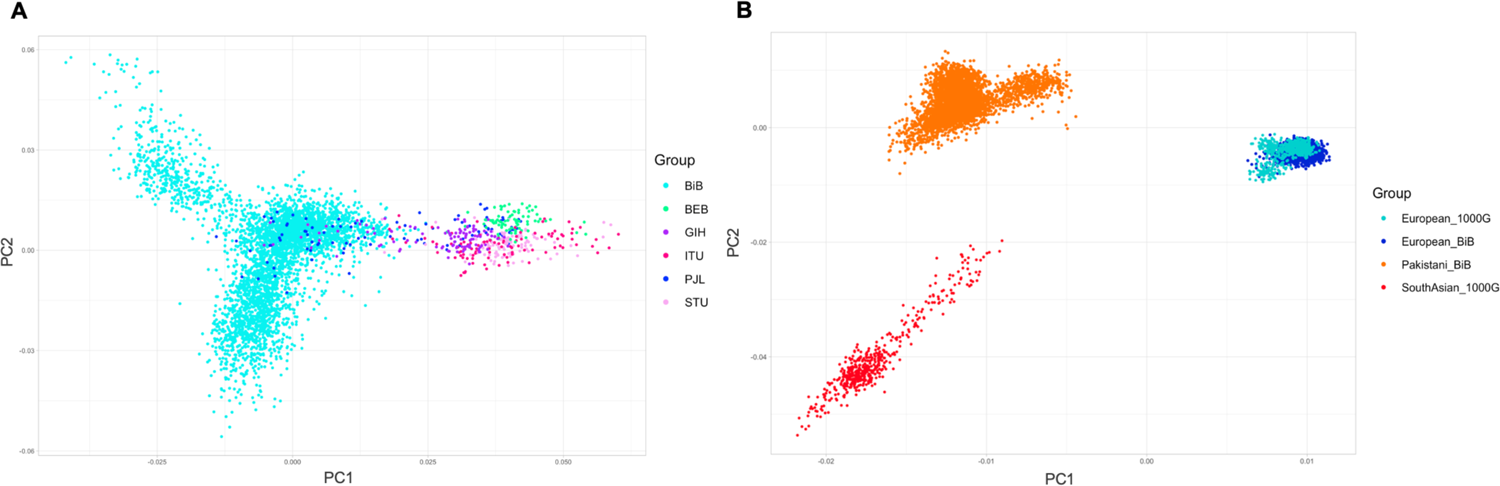
PCA comparing BiB data to all SA data from 1000G. A: PCA plot of SA BiB and SA data from 1000G. BiB: Born in Bradford, BEB: Bengali in Bangladesh, GIH: Gujarati Indian from Houston, Texas. ITU: Indian Telugu in the UK. PJL: Punjabi in Lahore, Pakistan STU: Sri Lankan Tamil in the UK. B: PCA plot of BiB data (WE and SA) and South Asian and European data from 1000G.

**Supplementary Figure 3:**
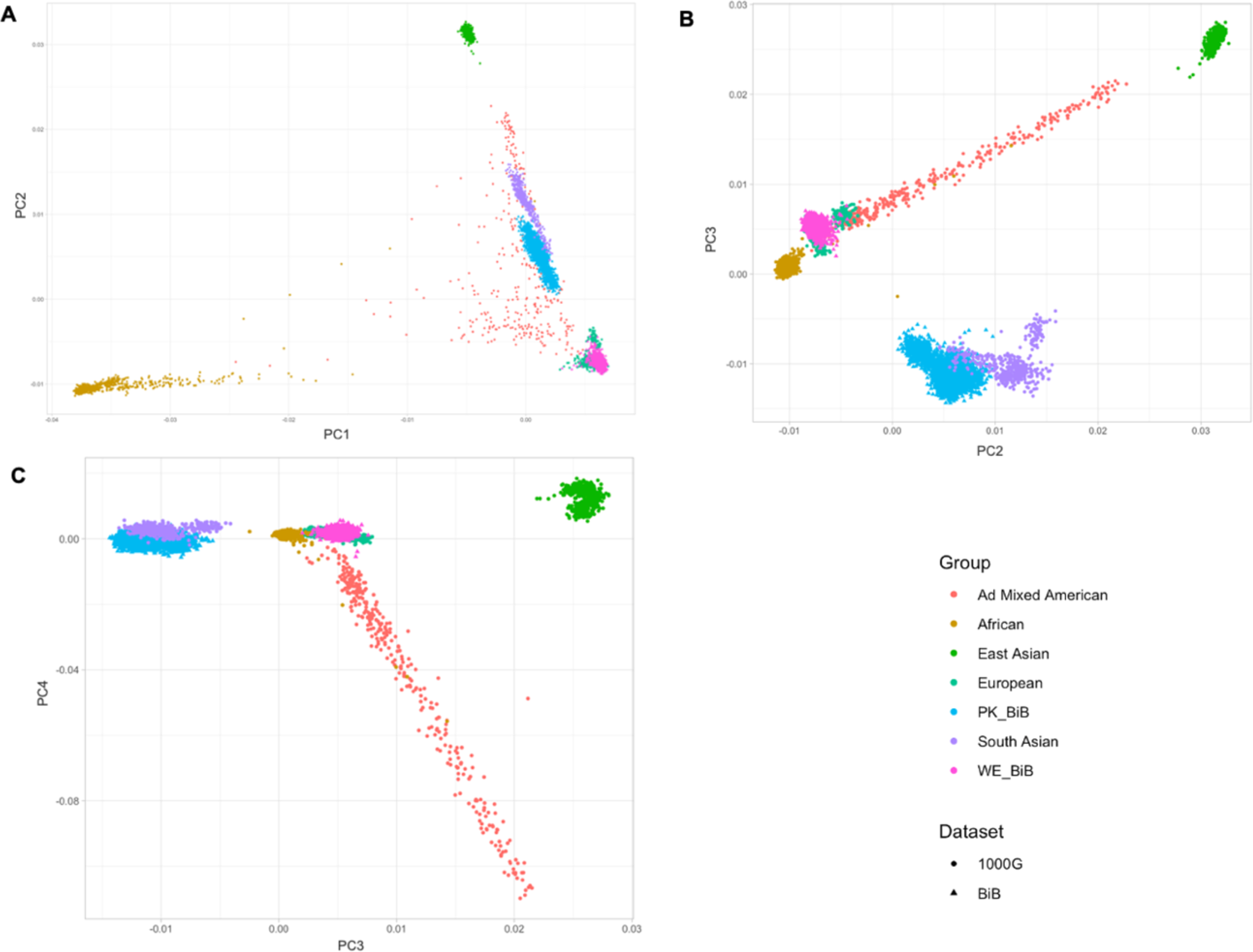
PCA comparing BiB data to all data from 1000G. A: PCA plot of PC1 vs PC2. B: PCA plot of PC2 vs PC3. C: PCA plot of PC3 vs PC4. PK_BIB: Pakistani BiB sample; WE_BIB: white European BiB Sample.

**Supplementary Figure 4:**
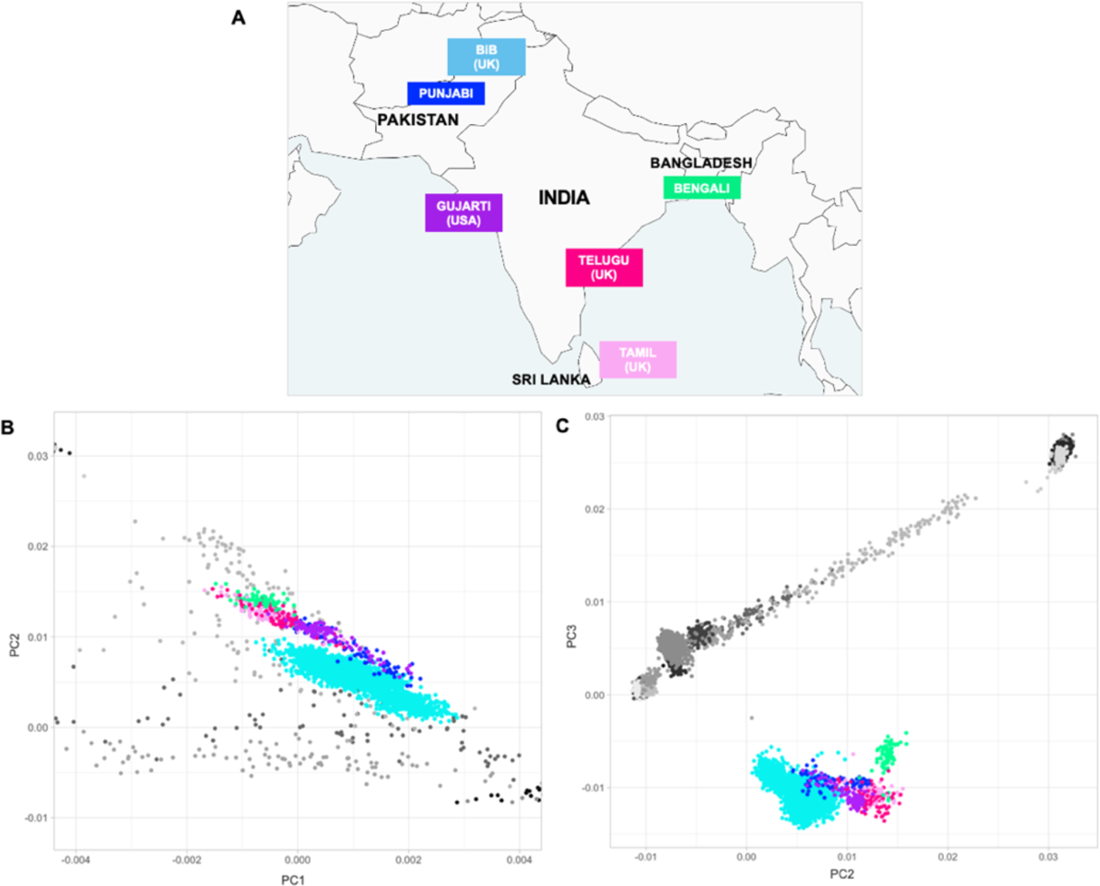
Modified Supplementary Figure 3 including the Pakistani BiB and South Asian 1000G populations.

**Supplementary Figure 5:**
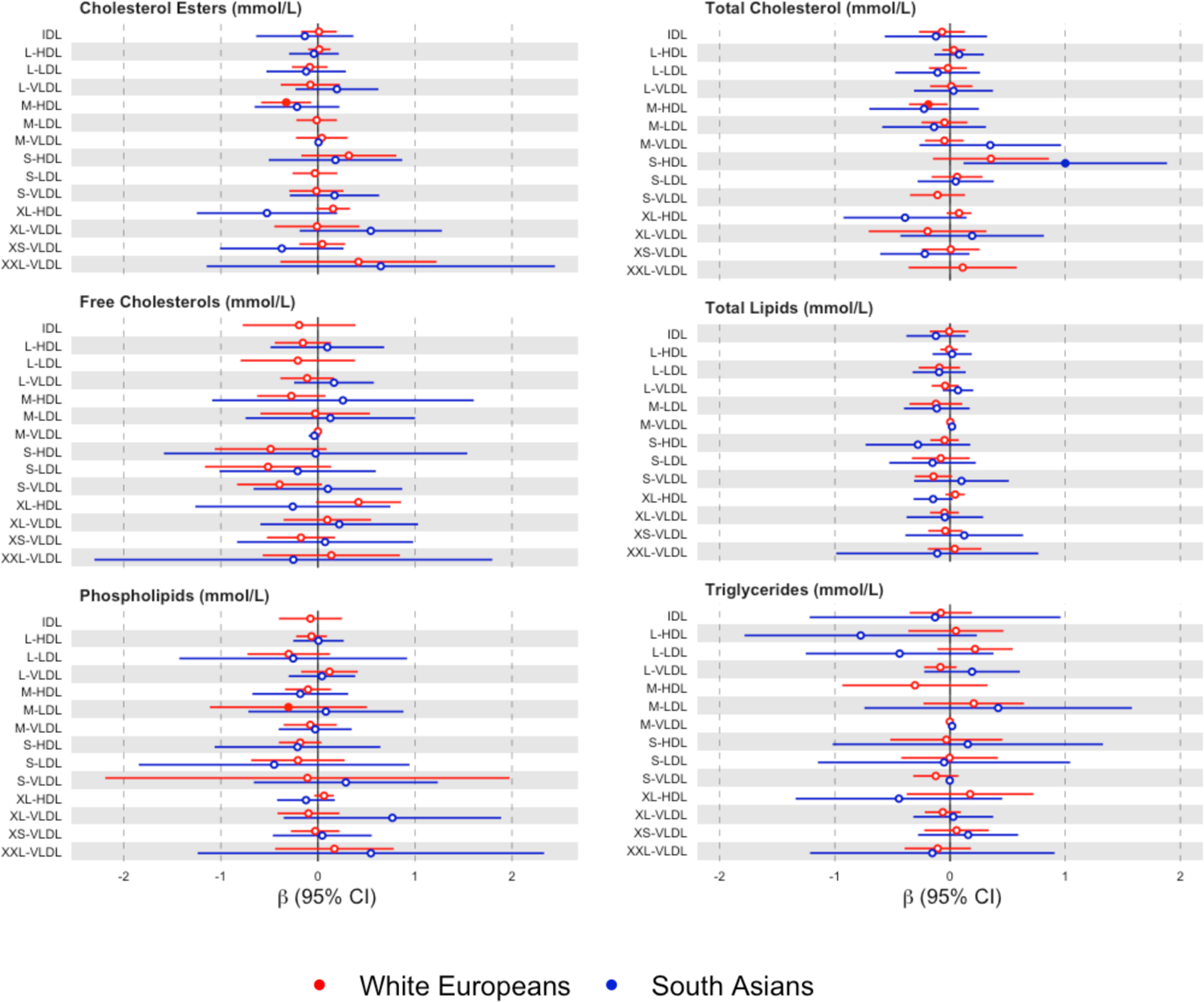
Forest plots of ethnic specific MR estimates for the association between metabolites and fasting glucose. Forest plots showing β values and 95% CIs of the association between metabolites and fasting glucose. Significant associations (p-value ≤ 0.05) are shown in shaded circles.

**Supplementary Figure 6:**
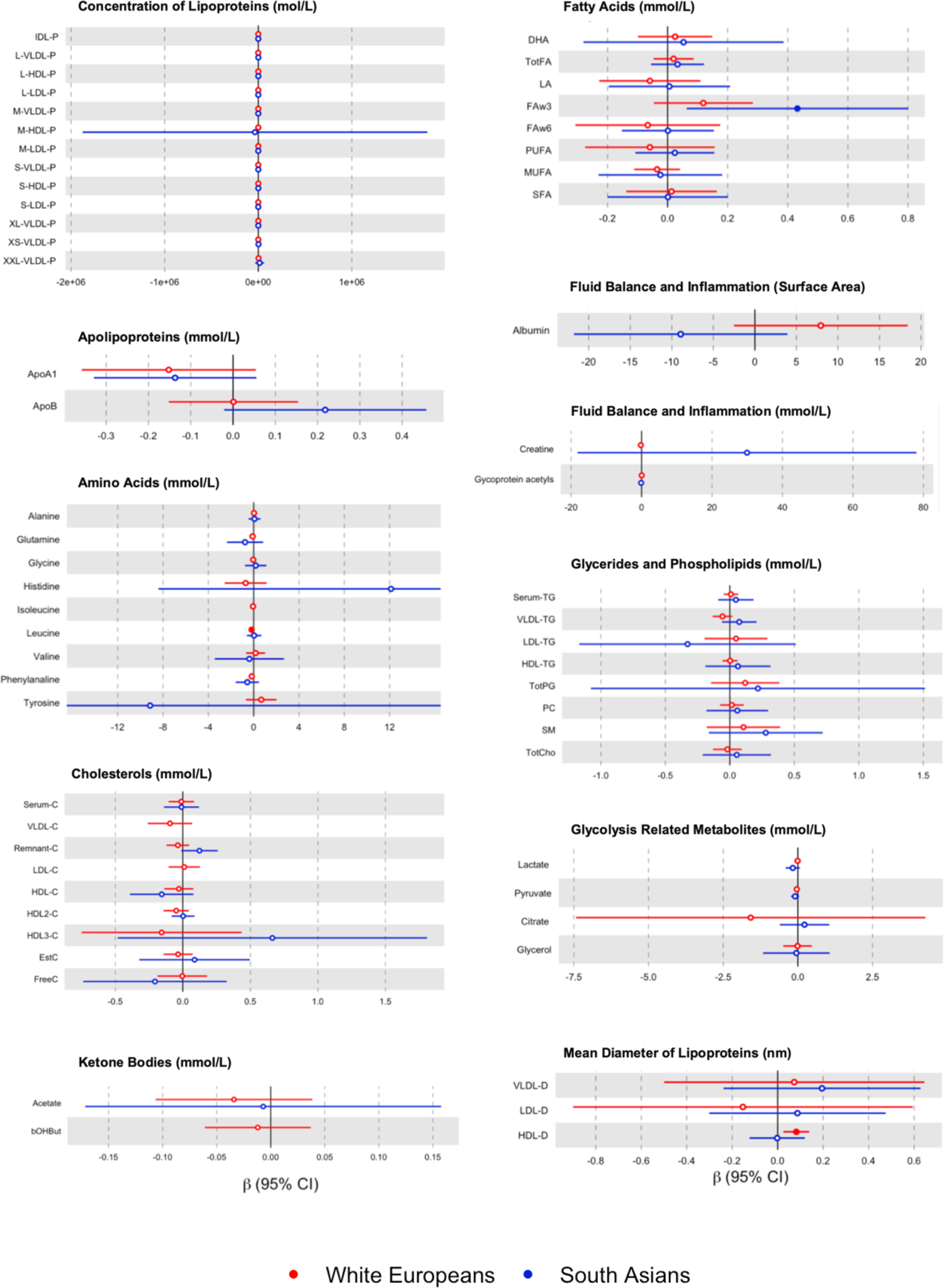
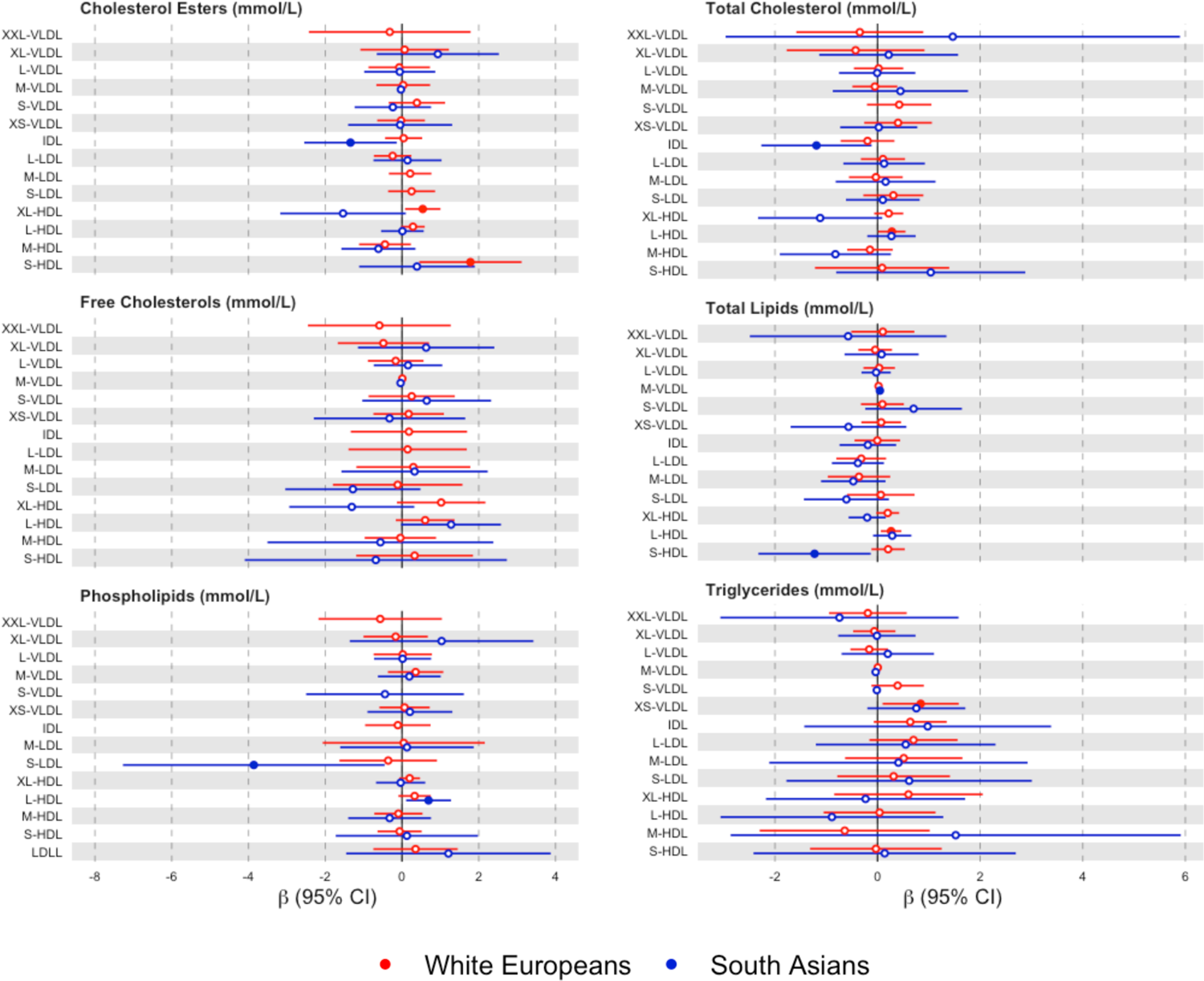
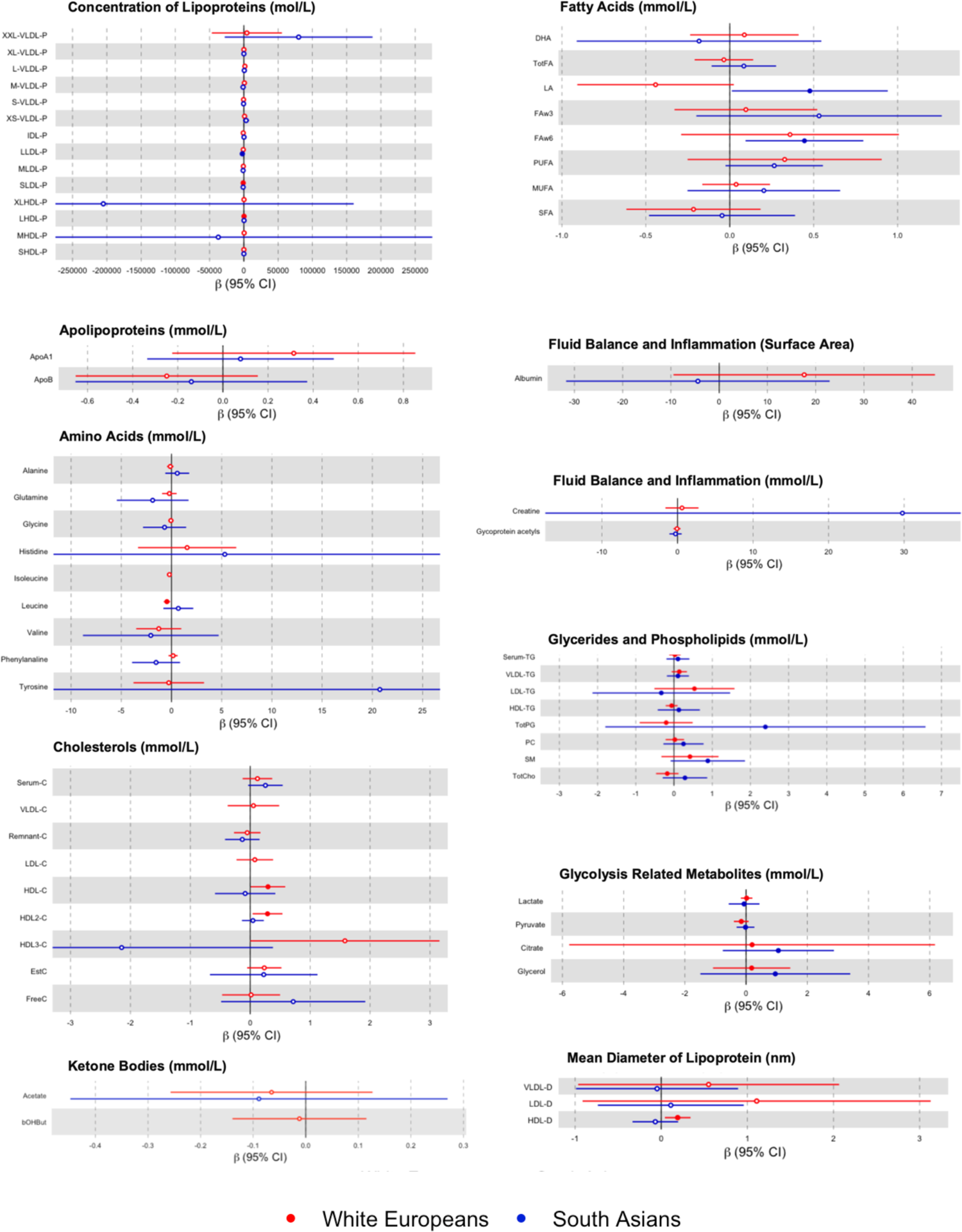
Forest plots of ethnic specific MR estimates for the association between metabolites and fasting glucose. Forest plots showing β values and 95% CIs of the association between metabolites and 2-hour post glucose. Significant associations (p-value ≤ 0.05) are shown in shaded circles.

**Supplementary Figure 7:**
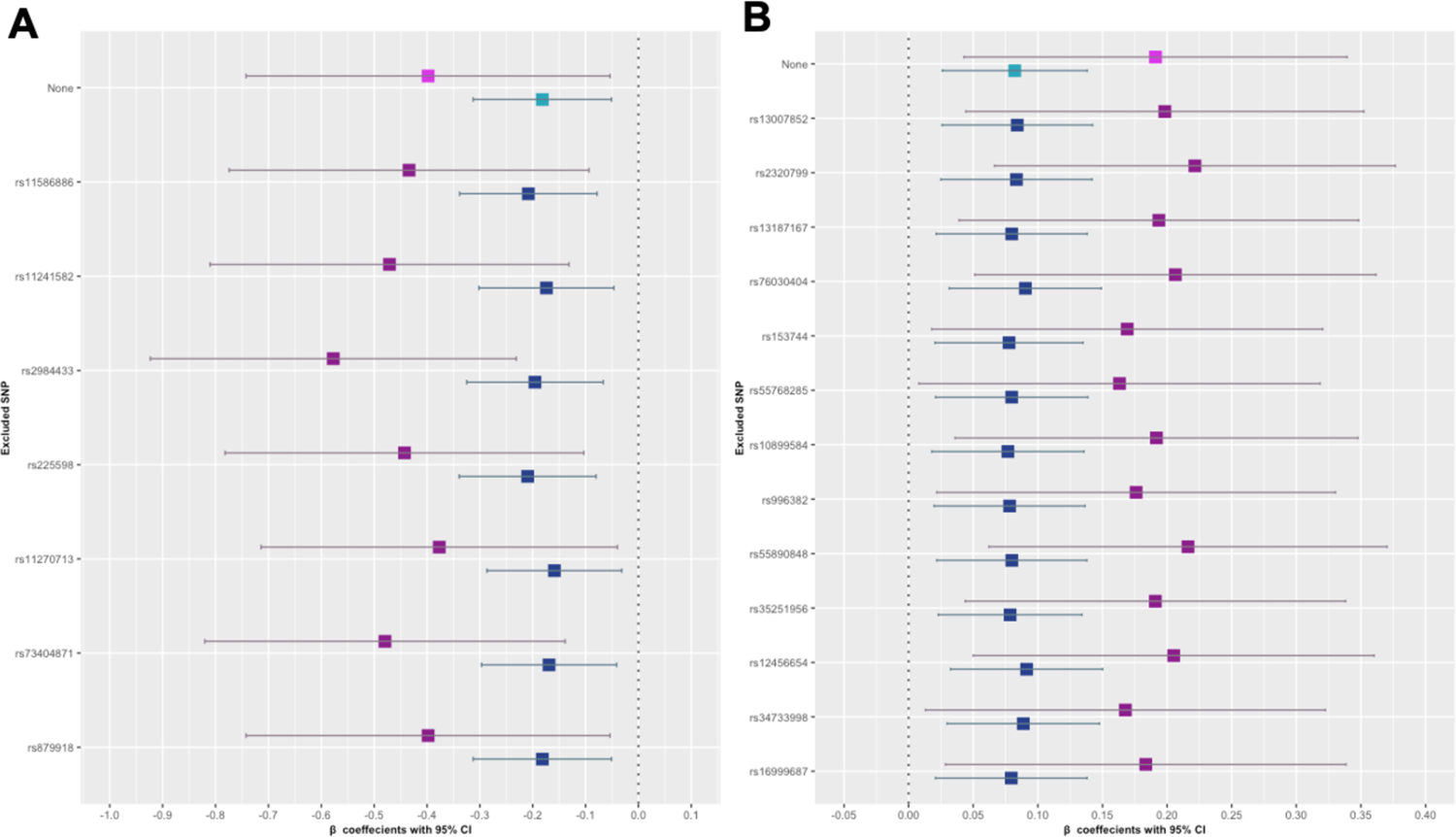
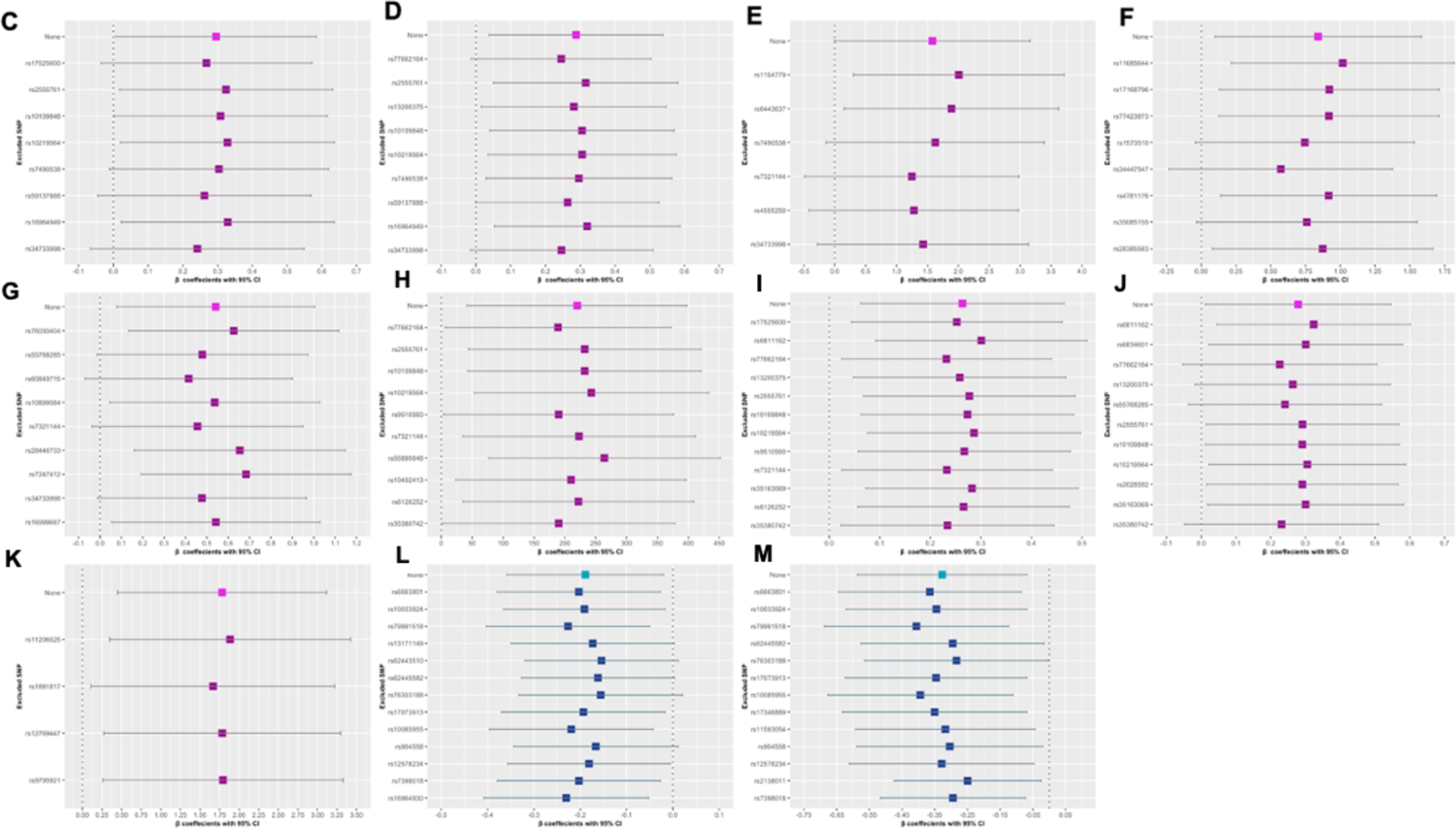
Forest plots of β values following leave-one-out analyses for identified associations in white Europeans. Forest plots showing β values and 95% CIs following the leave-one-out analyses of each SNP in each instrument. Dashed line represents no effect. Associations shown in purple indicating associations with 2-hour post glucose while associations shown in blue indicate associations with fasting glucose. A: Leucine. B: HDL_D. C: HDLC. D: HDL2C. E: HDL3C. F: XS-VLDL-TG. G: XL-HDL-CE. H: L-HDL-P. I: L-HDL_L. J: L-HDL-C. K: S-HDL-CE. L: M-HDL-C. M: M-HDL-CE.

**Supplementary Figure 8:**
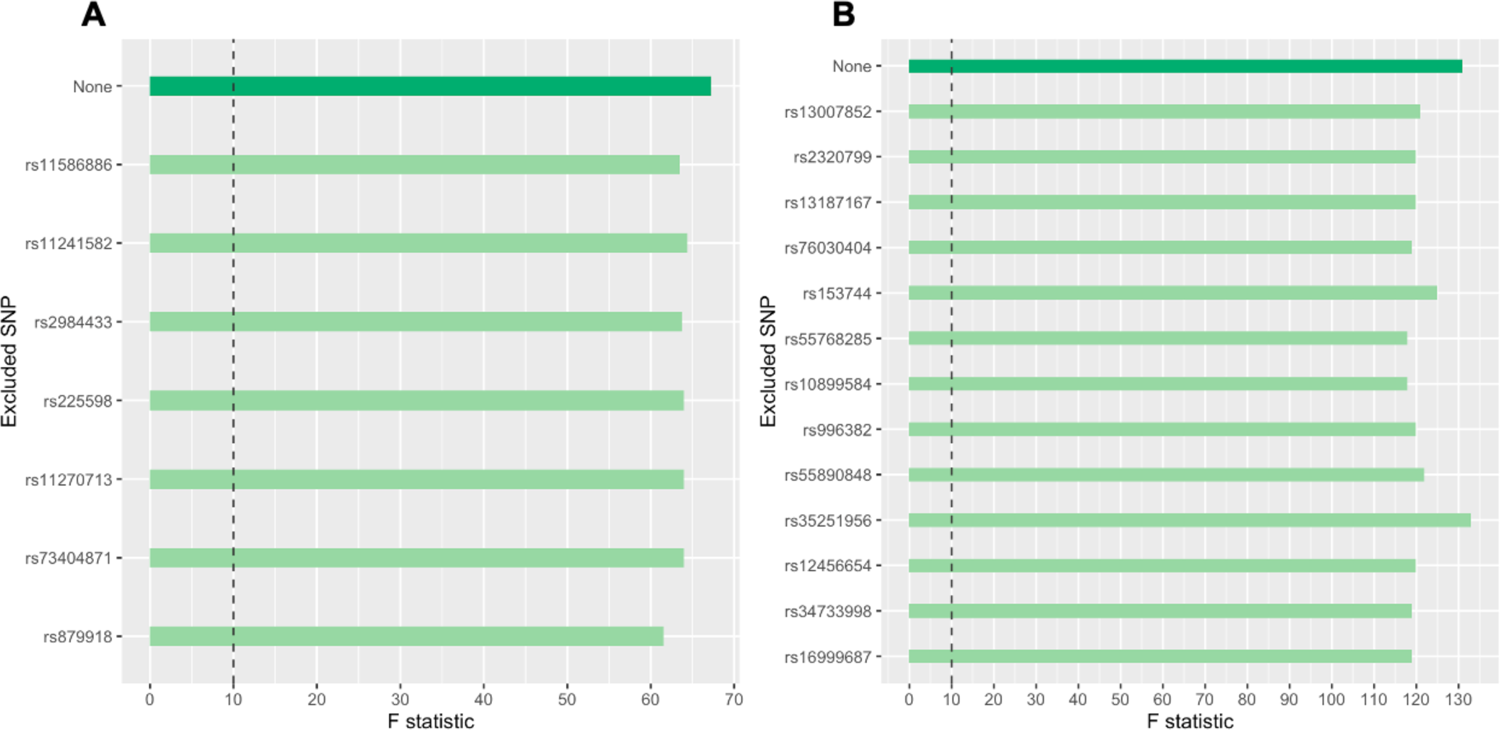
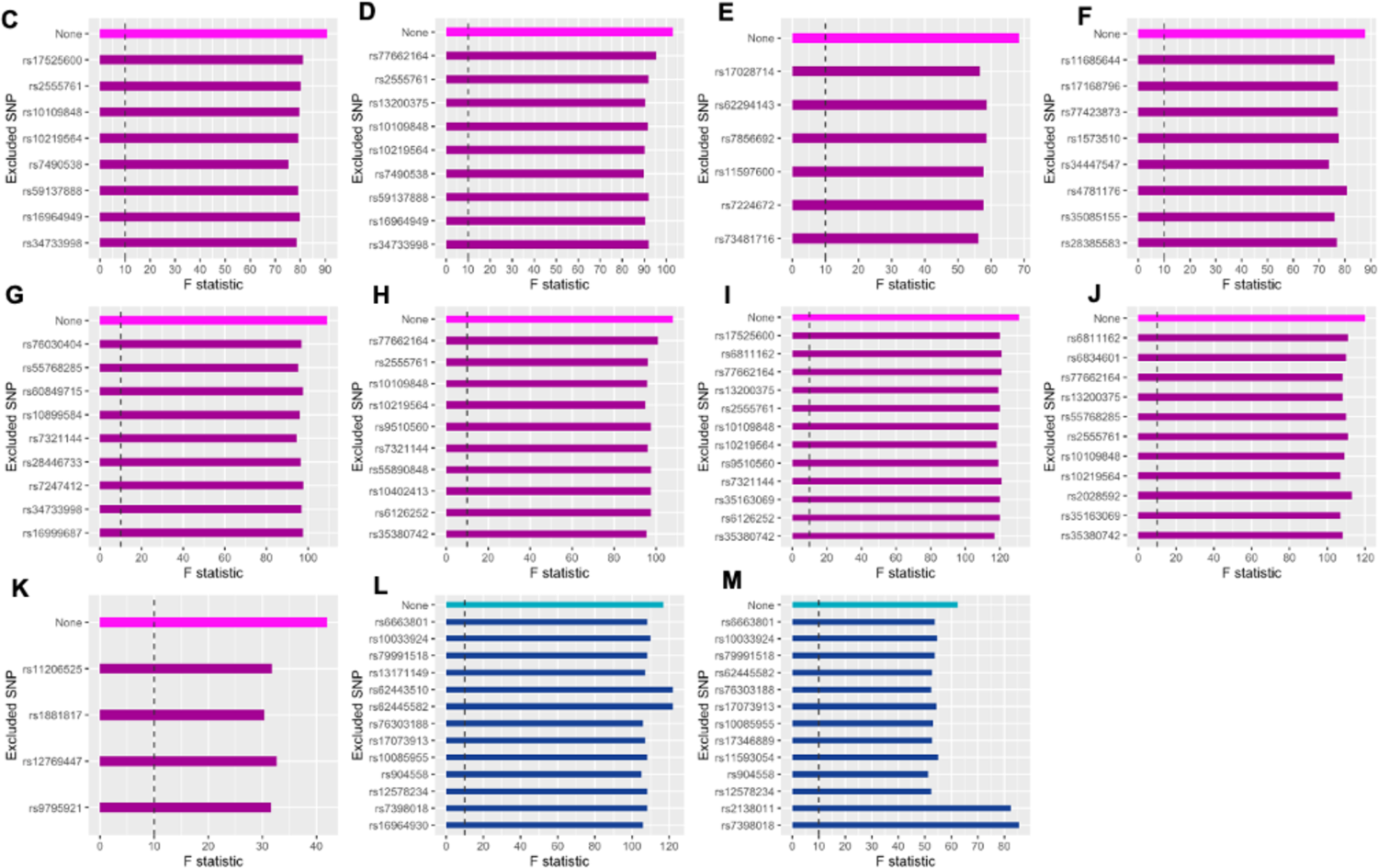
F-statistics following leave-one-out analyses for identified associations in white Europeans. Dashed line indicates an F-statistic of 10, below which an instrument is classified as weak. Associations shown in purple indicate associations with 2-hour post glucose while associations shown in blue indicate associations with fasting glucose. Green bars indicate metabolite measures associated with both fasting glucose and 2-hour post glucose. A: Leucine. B: HDL_D. C: HDLC. D: HDL2C. E: HDL3C. F: XS-VLDL-TG. G: XL-HDL-CE. H: L-HDL-P. I: L-HDL_L. J: L-HDL-C. K: S-HDL-CE. L: M-HDL-C. M: M-HDL-CE

**Supplementary Figure 9:**
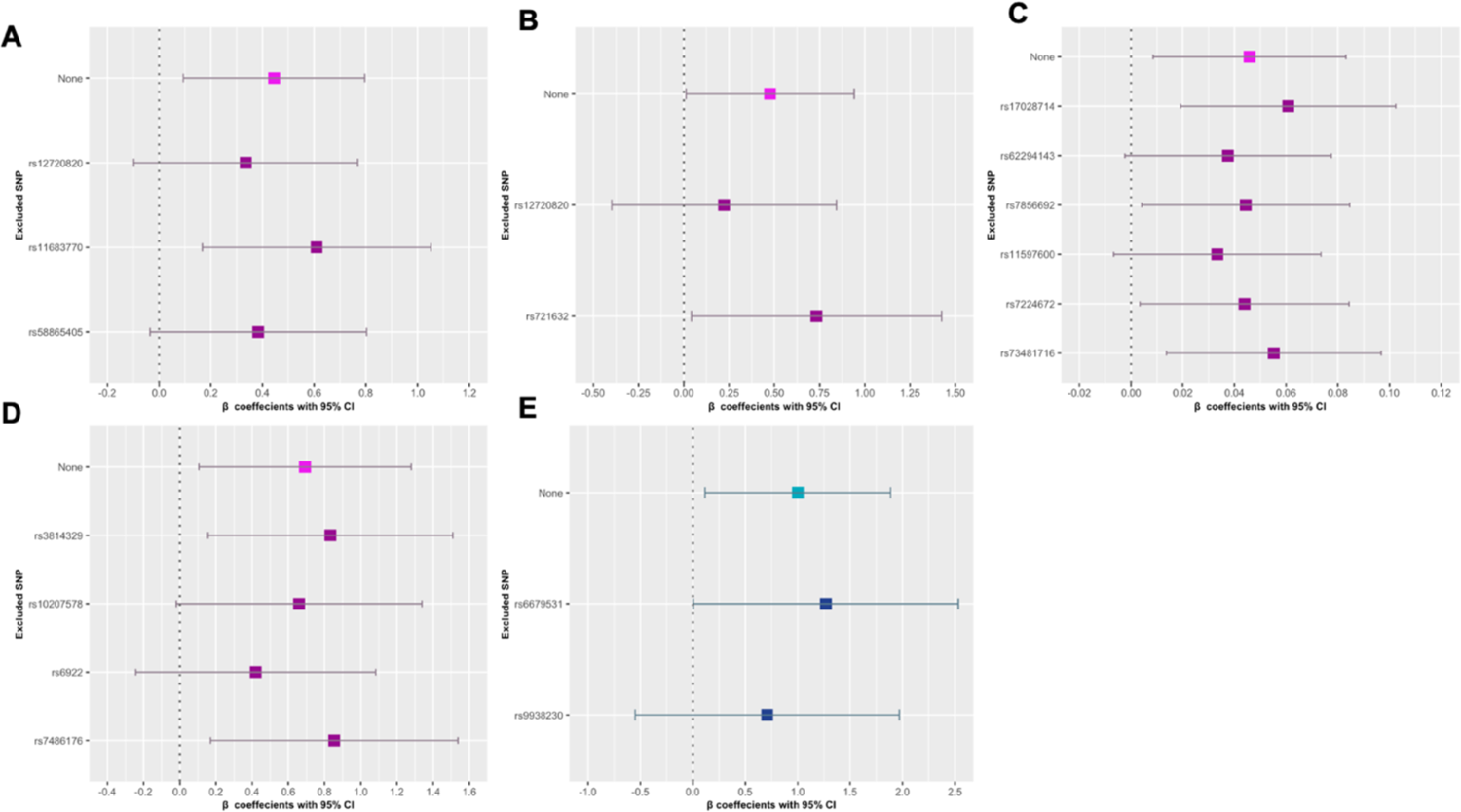
Forest plots of β values following leave-one-out analyses for identified associations in South Asians.

**Supplementary Figure 10:**
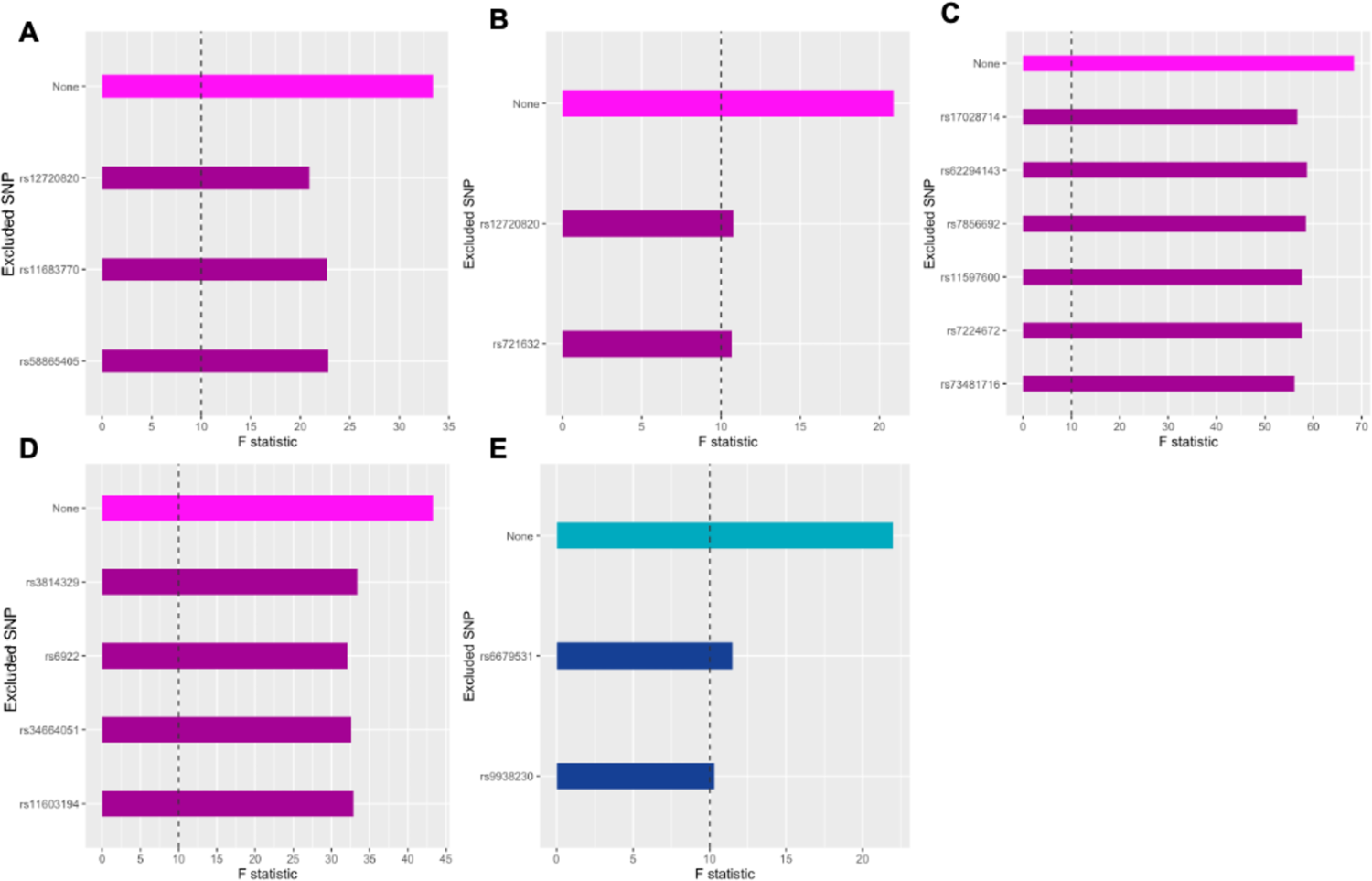
F-statistics following leave-one-out analyses for identified associations in South Asians. Dashed line indicates an F-statistic of 10, below which an instrument is classified as weak Associations shown in purple indicate associations with 2-hour post glucose while associations shown in blue indicate associations with fasting glucose. A: FAw6. B: LA. C: M-VLDL-L. D: L-HDL-PL. E: S-HDL-C.

**Supplementary Figure 11:**
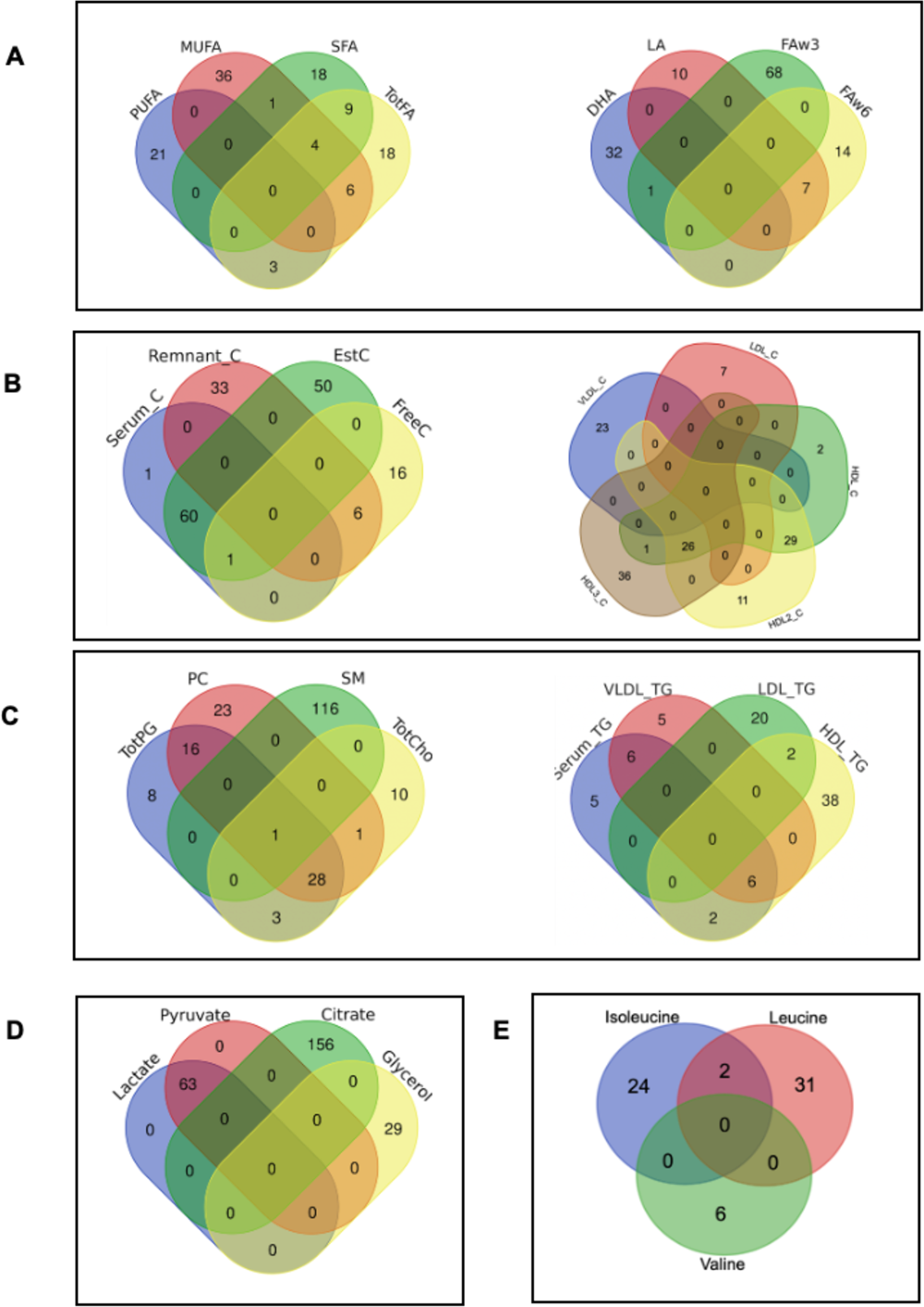
Overlap of SNPs identified by class in white Europeans. Venn diagram showing the overlap between suggestive (1 x 10^-5^) SNPs in WEs. SNPs in the apolipoprotein, lipoprotein density, ketone bodies and fluid balance/ inflammation, aromatic amino acids and non-branched amino acids were not found to overlap in WEs. Classes with ≥5 metabolites have been split for clarity. A: Fatty Acids. B: Cholesterols. C: Glycerides and Phospholipids. D: Glycolysis Related Metabolites. E: Branched Amino Acids. Plots were created using the online tool (https://bioinformatics.psb.ugent.be/webtools/Venn/) by the Bioinformatics and Genetics Group at the Department of Plant Systems Biology.

**Supplementary Figure 12:**
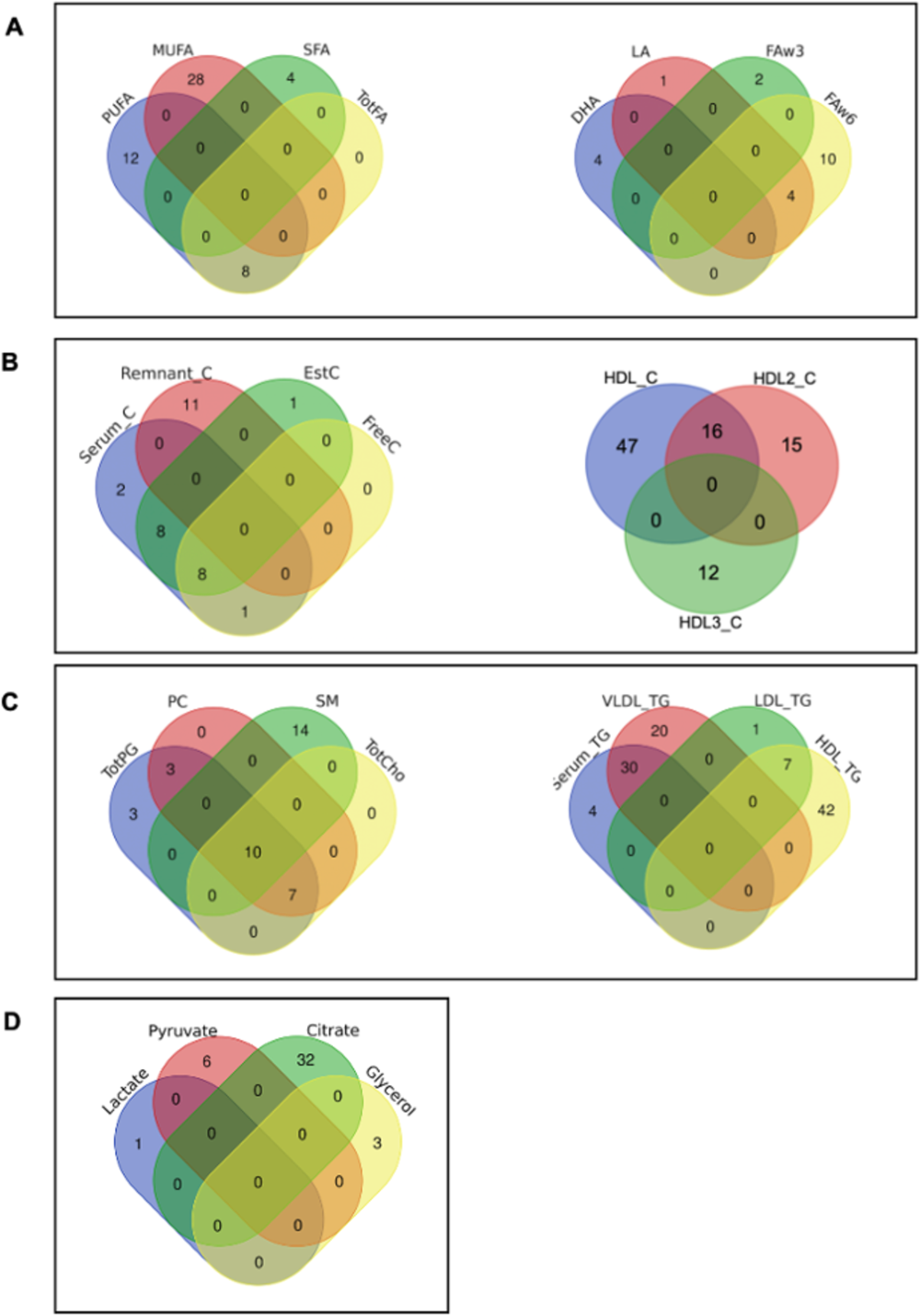
Overlap of SNPs identified by class in South Asians. Venn diagram highlighting the overlap between suggestive (1 x 10^-5^) SNPs in SAs. SNPs in the apolipoprotein, lipoprotein density, ketone Bodies and fluid Balance/ inflammation, and amino acids were not found to overlap in WEs. Classes with ≥5 metabolites have been split for clarity. A: Fatty Acids. B: Cholesterols. C: Glycerides and Phospholipids. D: Glycolysis Related Metabolites. Plots were created using the online tool (https://bioinformatics.psb.ugent.be/webtools/Venn/) by the Bioinformatics and Genetics Group at the Department of Plant Systems Biology.

**Supplementary Figure 13:**
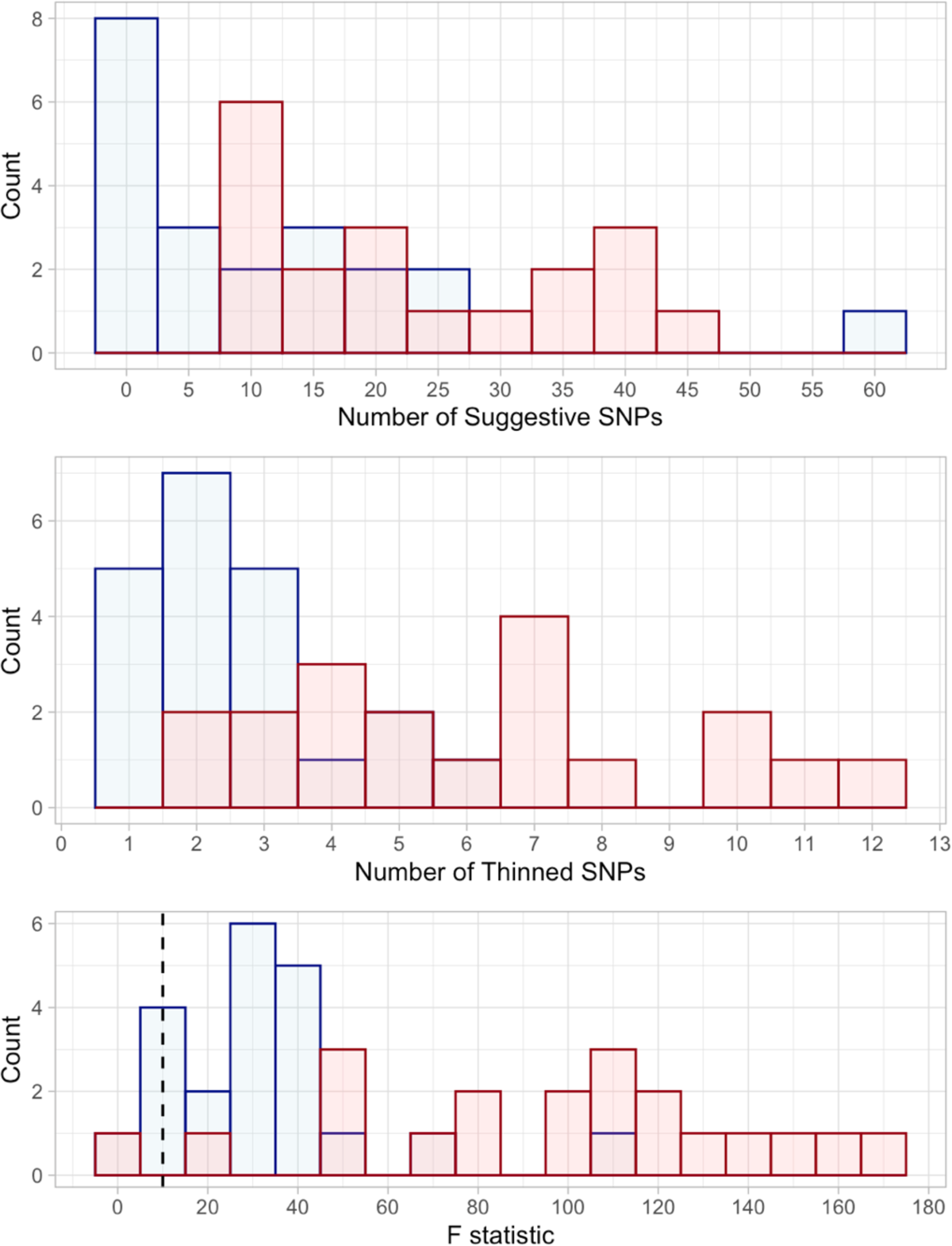
Histograms of the number of SNPs and strength of each instrument in the analysis of each metabolite class. A: Histogram of number of SNPs identified for each metabolite class at the suggestive level (p ≤ 1 x 10^-5^). B: Histogram of the number of SNPs remaining after thinning by LD (R^2^>0.2).C: Histogram of the F-statistics for each instrument. Dashed line shows an F-statistic of 10. An F-statistic < 10 is an indicator of weak instrument bias. Blue: South Asians. Red: white Europeans.

**Supplementary Figure 14:**
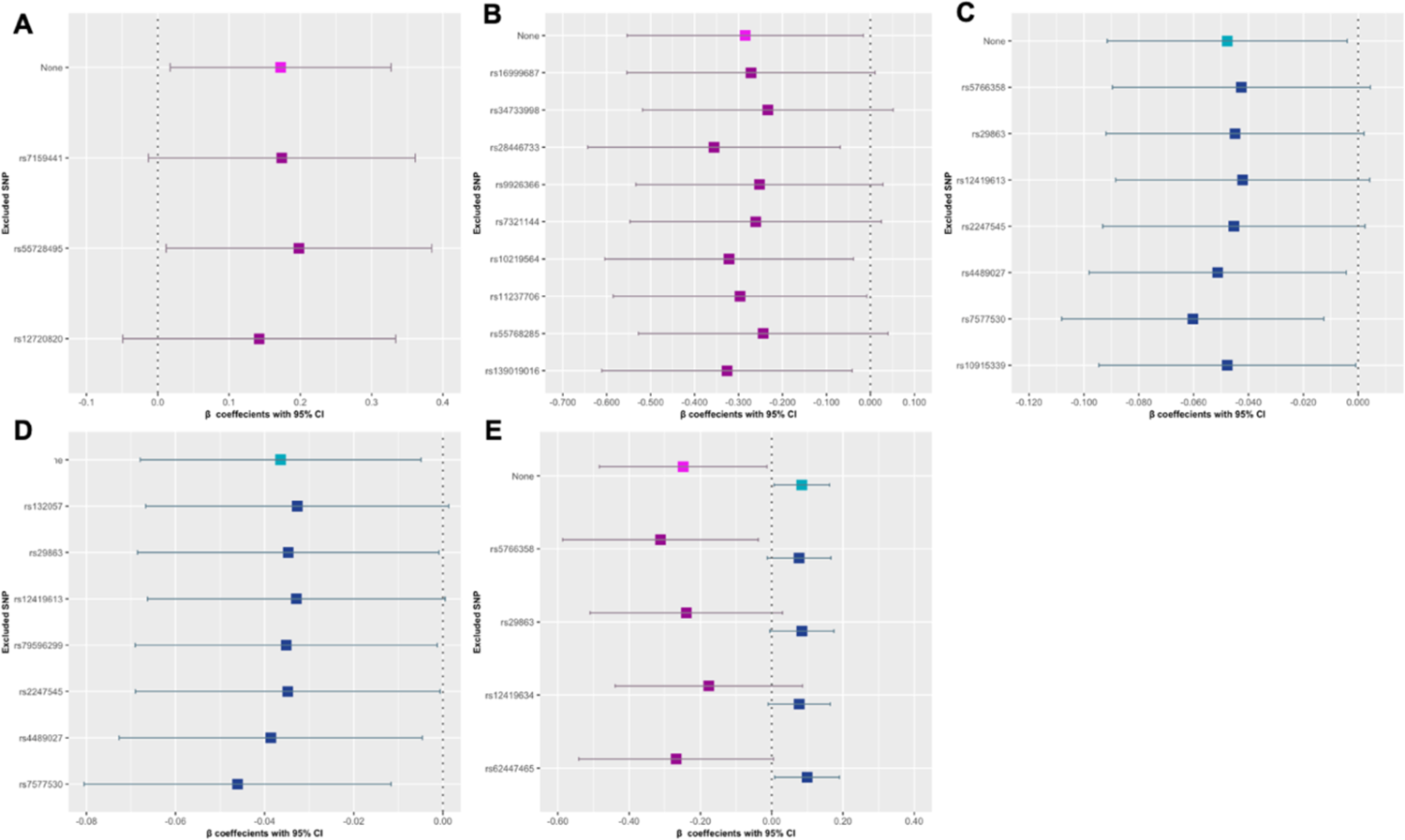
Forest plots of β values following leave-one-out analyses for identified associations in the analysis of metabolite class. Forest plots showing β values and 95% CIs following the leave-one-out analyses of each SNP in each instrument. Dashed line represents no effect. Associations shown in purple indicate associations with 2-hour post glucose while associations shown in blue indicate associations with fasting glucose. A: Fatty acid class, SAs. B: XL-HDL class, WEs. C: M-LDL class, WEs. D: All LDL class, WEs. E: S-LDL class, WEs.

**Supplementary Figure 15:**
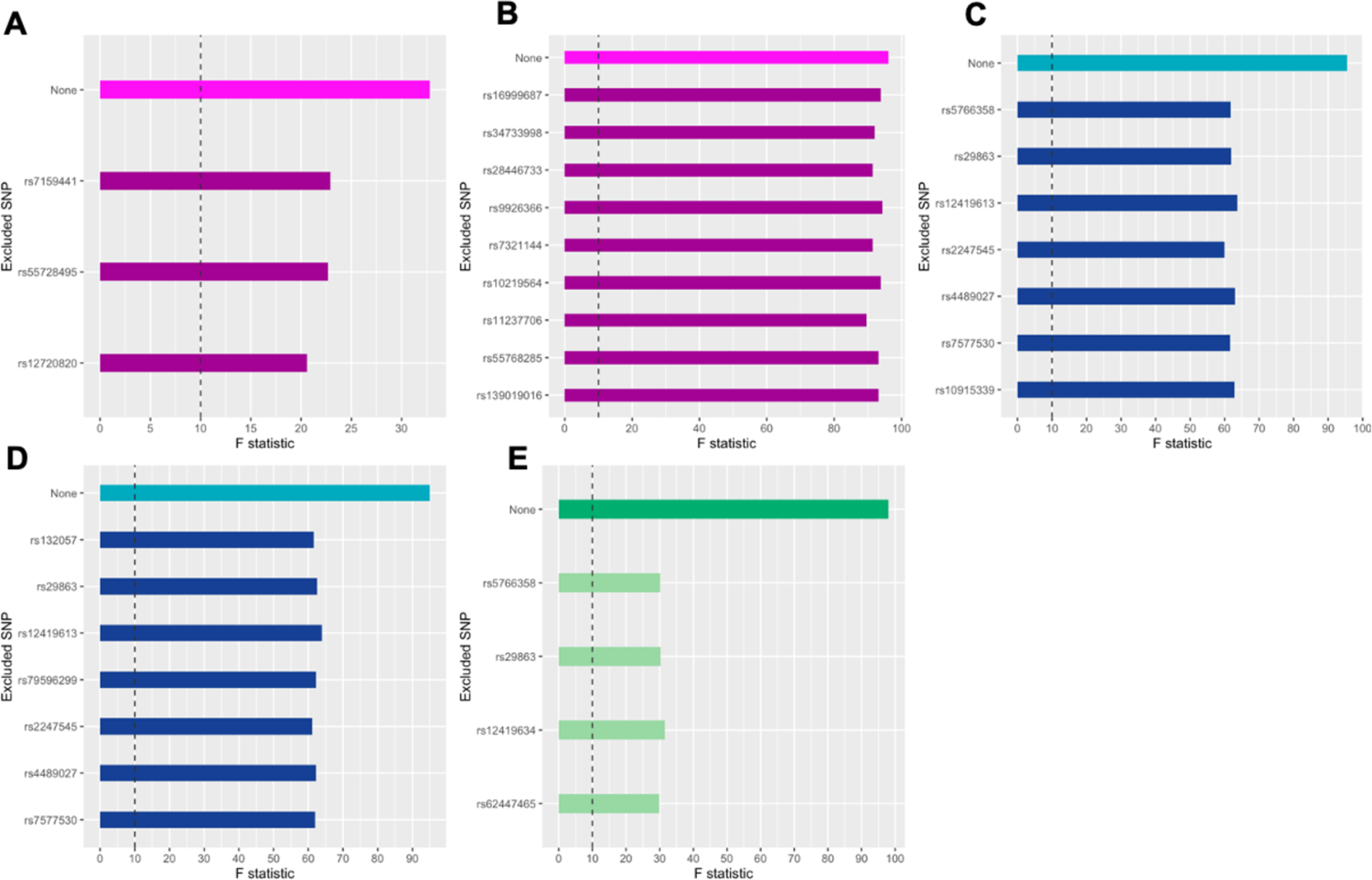
F-statistics following leave-one-out analyses for identified associations in the analysis of metabolite classes. Dashed line indicates an F-statistic of 10, below which an instrument is classified as weak. Associations shown in purple indicating associations with 2-hour post glucose while associations shown in blue indicate associations with fasting glucose. Green bars indicate metabolite measures associated with both fasting glucose and 2-hour post glucose. A: Fatty acid class, SAs. B: XL-HDL class, WEs. C: M-LDL class, WEs. D: All LDL class, WEs. E: S-LDL class, WEs.

**Supplementary Table 1:**
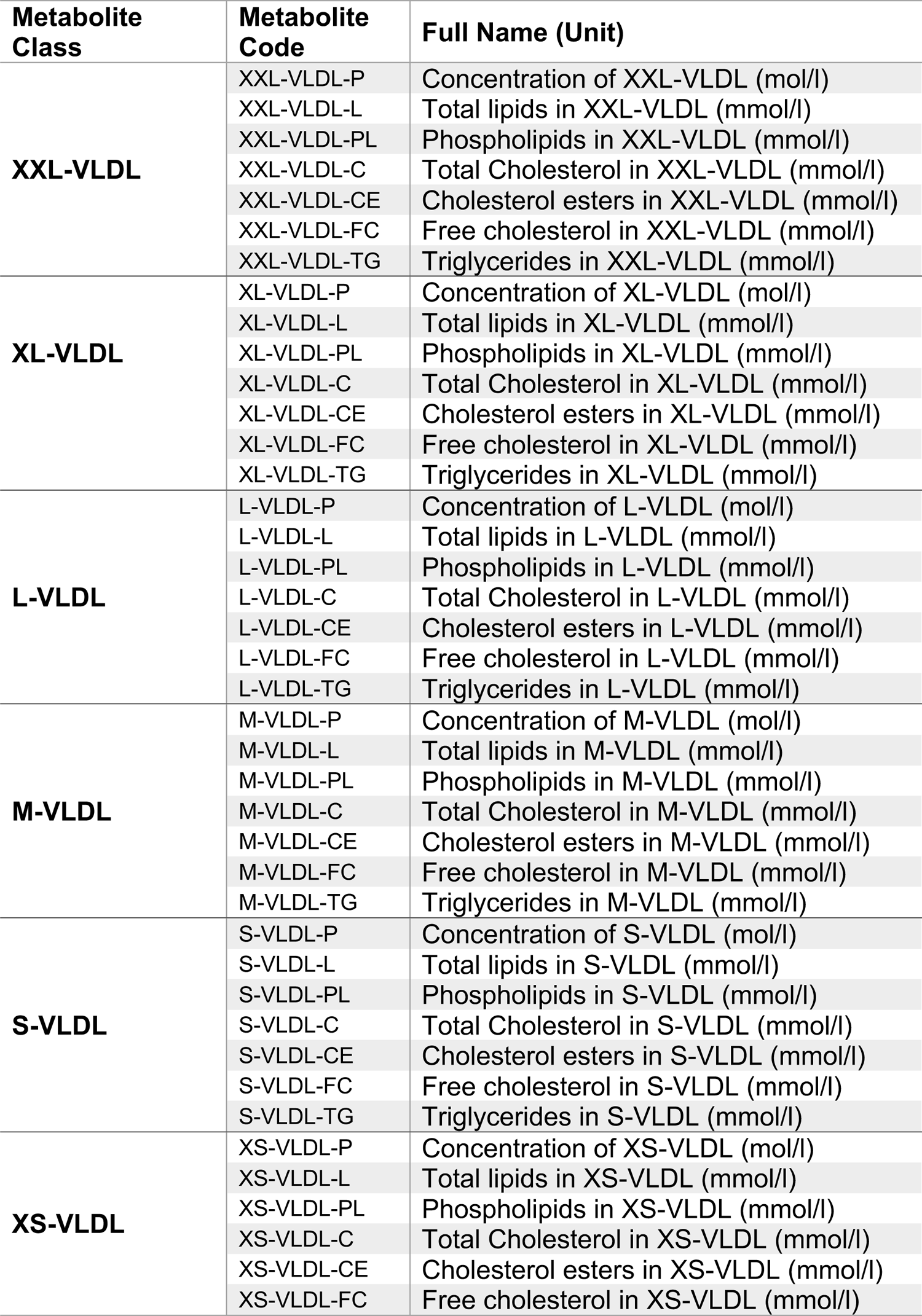
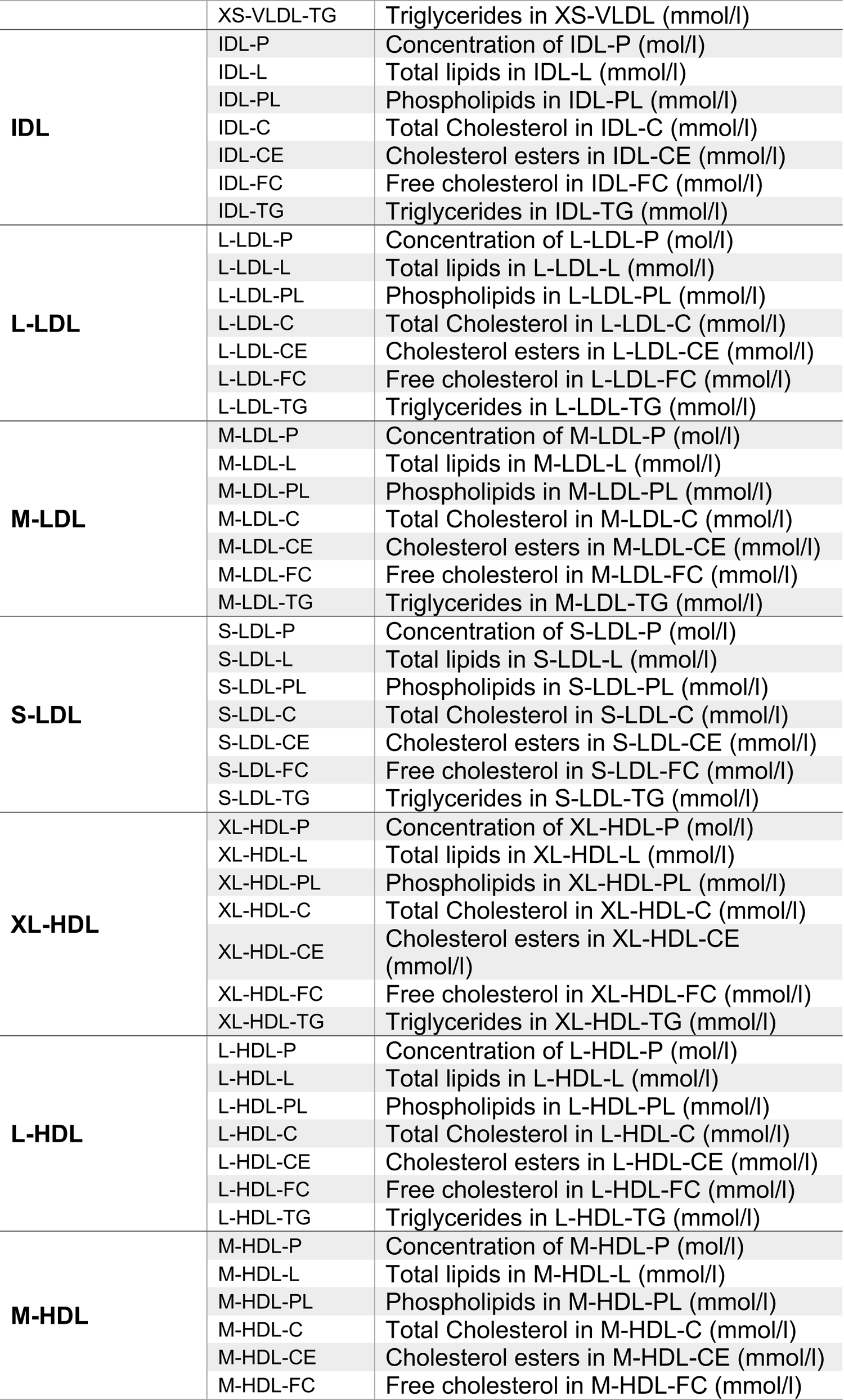
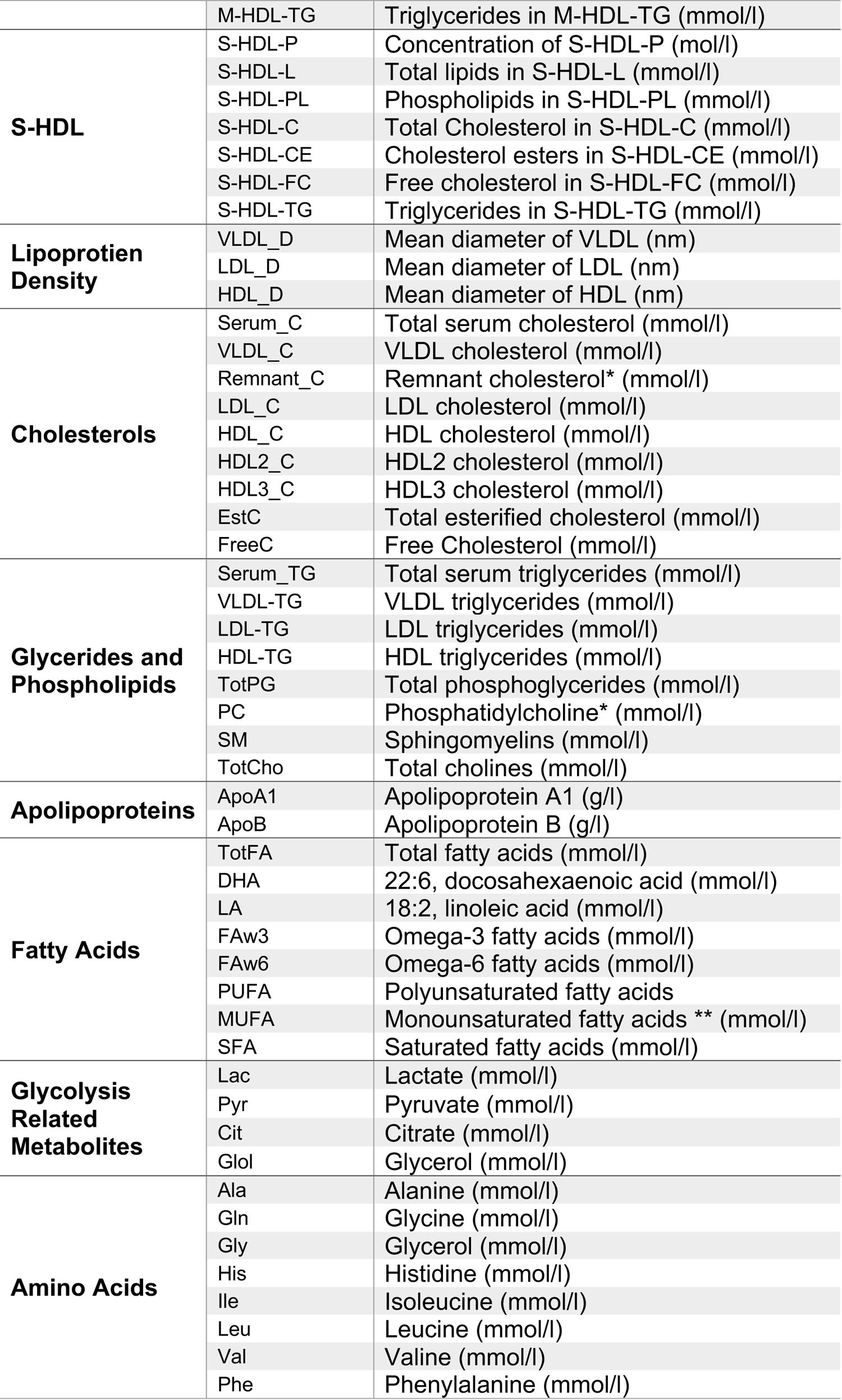
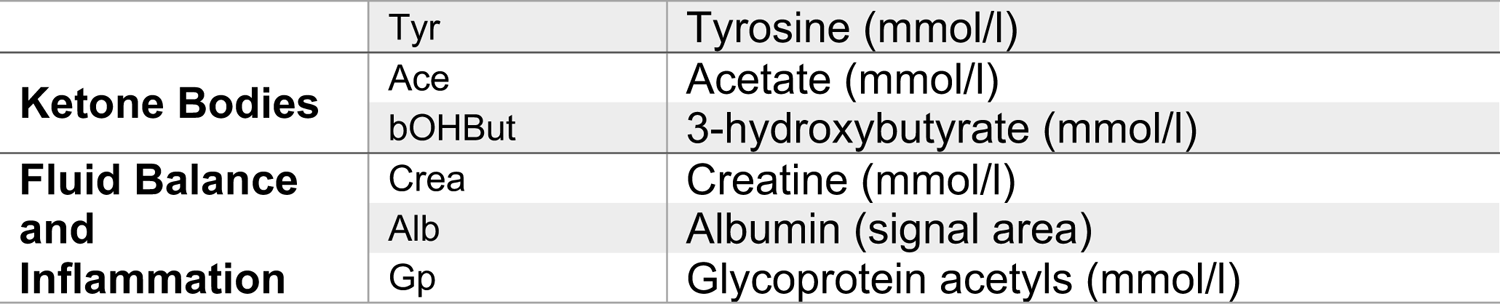
List of metabolite values included within this study.

| **Metabolite Class** | **Metabolite Code** | **Full Name (Unit)** |
| --- | --- | --- |
| **XXL-VLDL** | XXL-VLDL-P | Concentration of XXL-VLDL (mol/l) |
|  | XXL-VLDL-L | Total lipids in XXL-VLDL (mmol/l) |
|  | XXL-VLDL-PL | Phospholipids in XXL-VLDL (mmol/l) |
|  | XXL-VLDL-C | Total Cholesterol in XXL-VLDL (mmol/l) |
|  | XXL-VLDL-CE | Cholesterol esters in XXL-VLDL (mmol/l) |
|  | XXL-VLDL-FC | Free cholesterol in XXL-VLDL (mmol/l) |
|  | XXL-VLDL-TG | Triglycerides in XXL-VLDL (mmol/l) |
| **XL-VLDL** | XL-VLDL-P | Concentration of XL-VLDL (mol/l) |
|  | XL-VLDL-L | Total lipids in XL-VLDL (mmol/l) |
|  | XL-VLDL-PL | Phospholipids in XL-VLDL (mmol/l) |
|  | XL-VLDL-C | Total Cholesterol in XL-VLDL (mmol/l) |
|  | XL-VLDL-CE | Cholesterol esters in XL-VLDL (mmol/l) |
|  | XL-VLDL-FC | Free cholesterol in XL-VLDL (mmol/l) |
|  | XL-VLDL-TG | Triglycerides in XL-VLDL (mmol/l) |
| **L-VLDL** | L-VLDL-P | Concentration of L-VLDL (mol/l) |
|  | L-VLDL-L | Total lipids in L-VLDL (mmol/l) |
|  | L-VLDL-PL | Phospholipids in L-VLDL (mmol/l) |
|  | L-VLDL-C | Total Cholesterol in L-VLDL (mmol/l) |
|  | L-VLDL-CE | Cholesterol esters in L-VLDL (mmol/l) |
|  | L-VLDL-FC | Free cholesterol in L-VLDL (mmol/l) |
|  | L-VLDL-TG | Triglycerides in L-VLDL (mmol/l) |
| **M-VLDL** | M-VLDL-P | Concentration of M-VLDL (mol/l) |
|  | M-VLDL-L | Total lipids in M-VLDL (mmol/l) |
|  | M-VLDL-PL | Phospholipids in M-VLDL (mmol/l) |
|  | M-VLDL-C | Total Cholesterol in M-VLDL (mmol/l) |
|  | M-VLDL-CE | Cholesterol esters in M-VLDL (mmol/l) |
|  | M-VLDL-FC | Free cholesterol in M-VLDL (mmol/l) |
|  | M-VLDL-TG | Triglycerides in M-VLDL (mmol/l) |
| **S-VLDL** | S-VLDL-P | Concentration of S-VLDL (mol/l) |
|  | S-VLDL-L | Total lipids in S-VLDL (mmol/l) |
|  | S-VLDL-PL | Phospholipids in S-VLDL (mmol/l) |
|  | S-VLDL-C | Total Cholesterol in S-VLDL (mmol/l) |
|  | S-VLDL-CE | Cholesterol esters in S-VLDL (mmol/l) |
|  | S-VLDL-FC | Free cholesterol in S-VLDL (mmol/l) |
|  | S-VLDL-TG | Triglycerides in S-VLDL (mmol/l) |
| **XS-VLDL** | XS-VLDL-P | Concentration of XS-VLDL (mol/l) |
|  | XS-VLDL-L | Total lipids in XS-VLDL (mmol/l) |
|  | XS-VLDL-PL | Phospholipids in XS-VLDL (mmol/l) |
|  | XS-VLDL-C | Total Cholesterol in XS-VLDL (mmol/l) |
|  | XS-VLDL-CE | Cholesterol esters in XS-VLDL (mmol/l) |
|  | XS-VLDL-FC | Free cholesterol in XS-VLDL (mmol/l) |
|  | XS-VLDL-TG | Triglycerides in XS-VLDL (mmol/l) |
| **IDL** | IDL-P | Concentration of IDL-P (mol/l) |
|  | IDL-L | Total lipids in IDL-L (mmol/l) |
|  | IDL-PL | Phospholipids in IDL-PL (mmol/l) |
|  | IDL-C | Total Cholesterol in IDL-C (mmol/l) |
|  | IDL-CE | Cholesterol esters in IDL-CE (mmol/l) |
|  | IDL-FC | Free cholesterol in IDL-FC (mmol/l) |
|  | IDL-TG | Triglycerides in IDL-TG (mmol/l) |
| **L-LDL** | L-LDL-P | Concentration of L-LDL-P (mol/l) |
|  | L-LDL-L | Total lipids in L-LDL-L (mmol/l) |
|  | L-LDL-PL | Phospholipids in L-LDL-PL (mmol/l) |
|  | L-LDL-C | Total Cholesterol in L-LDL-C (mmol/l) |
|  | L-LDL-CE | Cholesterol esters in L-LDL-CE (mmol/l) |
|  | L-LDL-FC | Free cholesterol in L-LDL-FC (mmol/l) |
|  | L-LDL-TG | Triglycerides in L-LDL-TG (mmol/l) |
| **M-LDL** | M-LDL-P | Concentration of M-LDL-P (mol/l) |
|  | M-LDL-L | Total lipids in M-LDL-L (mmol/l) |
|  | M-LDL-PL | Phospholipids in M-LDL-PL (mmol/l) |
|  | M-LDL-C | Total Cholesterol in M-LDL-C (mmol/l) |
|  | M-LDL-CE | Cholesterol esters in M-LDL-CE (mmol/l) |
|  | M-LDL-FC | Free cholesterol in M-LDL-FC (mmol/l) |
|  | M-LDL-TG | Triglycerides in M-LDL-TG (mmol/l) |
| **S-LDL** | S-LDL-P | Concentration of S-LDL-P (mol/l) |
|  | S-LDL-L | Total lipids in S-LDL-L (mmol/l) |
|  | S-LDL-PL | Phospholipids in S-LDL-PL (mmol/l) |
|  | S-LDL-C | Total Cholesterol in S-LDL-C (mmol/l) |
|  | S-LDL-CE | Cholesterol esters in S-LDL-CE (mmol/l) |
|  | S-LDL-FC | Free cholesterol in S-LDL-FC (mmol/l) |
|  | S-LDL-TG | Triglycerides in S-LDL-TG (mmol/l) |
| **XL-HDL** | XL-HDL-P | Concentration of XL-HDL-P (mol/l) |
|  | XL-HDL-L | Total lipids in XL-HDL-L (mmol/l) |
|  | XL-HDL-PL | Phospholipids in XL-HDL-PL (mmol/l) |
|  | XL-HDL-C | Total Cholesterol in XL-HDL-C (mmol/l) |
|  | XL-HDL-CE | Cholesterol esters in XL-HDL-CE (mmol/l) |
|  | XL-HDL-FC | Free cholesterol in XL-HDL-FC (mmol/l) |
|  | XL-HDL-TG | Triglycerides in XL-HDL-TG (mmol/l) |
| **L-HDL** | L-HDL-P | Concentration of L-HDL-P (mol/l) |
|  | L-HDL-L | Total lipids in L-HDL-L (mmol/l) |
|  | L-HDL-PL | Phospholipids in L-HDL-PL (mmol/l) |
|  | L-HDL-C | Total Cholesterol in L-HDL-C (mmol/l) |
|  | L-HDL-CE | Cholesterol esters in L-HDL-CE (mmol/l) |
|  | L-HDL-FC | Free cholesterol in L-HDL-FC (mmol/l) |
|  | L-HDL-TG | Triglycerides in L-HDL-TG (mmol/l) |
| **M-HDL** | M-HDL-P | Concentration of M-HDL-P (mol/l) |
|  | M-HDL-L | Total lipids in M-HDL-L (mmol/l) |
|  | M-HDL-PL | Phospholipids in M-HDL-PL (mmol/l) |
|  | M-HDL-C | Total Cholesterol in M-HDL-C (mmol/l) |
|  | M-HDL-CE | Cholesterol esters in M-HDL-CE (mmol/l) |
|  | M-HDL-FC | Free cholesterol in M-HDL-FC (mmol/l) |
|  | M-HDL-TG | Triglycerides in M-HDL-TG (mmol/l) |
| **S-HDL** | S-HDL-P | Concentration of S-HDL-P (mol/l) |
|  | S-HDL-L | Total lipids in S-HDL-L (mmol/l) |
|  | S-HDL-PL | Phospholipids in S-HDL-PL (mmol/l) |
|  | S-HDL-C | Total Cholesterol in S-HDL-C (mmol/l) |
|  | S-HDL-CE | Cholesterol esters in S-HDL-CE (mmol/l) |
|  | S-HDL-FC | Free cholesterol in S-HDL-FC (mmol/l) |
|  | S-HDL-TG | Triglycerides in S-HDL-TG (mmol/l) |
| **Lipoprotein Density** | VLDL_D | Mean diameter of VLDL (nm) |
|  | LDL_D | Mean diameter of LDL (nm) |
|  | HDL_D | Mean diameter of HDL (nm) |
| **Cholesterols** | Serum_C | Total serum cholesterol (mmol/l) |
|  | VLDL_C | VLDL cholesterol (mmol/l) |
|  | Remnant_C | Remnant cholesterol* (mmol/l) |
|  | LDL_C | LDL cholesterol (mmol/l) |
|  | HDL_C | HDL cholesterol (mmol/l) |
|  | HDL2_C | HDL2 cholesterol (mmol/l) |
|  | HDL3_C | HDL3 cholesterol (mmol/l) |
|  | EstC | Total esterified cholesterol (mmol/l) |
|  | FreeC | Free Cholesterol (mmol/l) |
| **Glycerides and Phospholipids** | Serum_TG | Total serum triglycerides (mmol/l) |
|  | VLDL-TG | VLDL triglycerides (mmol/l) |
|  | LDL-TG | LDL triglycerides (mmol/l) |
|  | HDL-TG | HDL triglycerides (mmol/l) |
|  | TotPG | Total phosphoglycerides (mmol/l) |
|  | PC | Phosphatidylcholine* (mmol/l) |
|  | SM | Sphingomyelins (mmol/l) |
|  | TotCho | Total cholines (mmol/l) |
| **Apolipoproteins** | ApoA1 | Apolipoprotein A1 (g/l) |
|  | ApoB | Apolipoprotein B (g/l) |
| **Fatty Acids** | TotFA | Total fatty acids (mmol/l) |
|  | DHA | 22:6, docosahexaenoic acid (mmol/l) |
|  | LA | 18:2, linoleic acid (mmol/l) |
|  | FAw3 | Omega-3 fatty acids (mmol/l) |
|  | FAw6 | Omega-6 fatty acids (mmol/l) |
|  | PUFA | Polyunsaturated fatty acids |
|  | MUFA | Monounsaturated fatty acids ** (mmol/l) |
|  | SFA | Saturated fatty acids (mmol/l) |
| **Glycolysis Related Metabolites** | Lac | Lactate (mmol/l) |
|  | Pyr | Pyruvate (mmol/l) |
|  | Cit | Citrate (mmol/l) |
|  | GloI | Glycerol (mmol/l) |
| **Amino Acids** | Ala | Alanine (mmol/l) |
|  | Gln | Glycine (mmol/l) |
|  | Gly | Glycerol (mmol/l) |
|  | His | Histidine (mmol/l) |
|  | Ile | Isoleucine (mmol/l) |
|  | Leu | Leucine (mmol/l) |
|  | Val | Valine (mmol/l) |
|  | Phe | Phenylalanine (mmol/l) |
|  | Tyr | Tyrosine (mmol/l) |
| **Ketone Bodies** | Ace | Acetate (mmol/l) |
|  | bOHBut | 3-hydroxybutyrate (mmol/l) |
| **Fluid Balance**<br>**and**<br>**Inflammation** | Crea | Creatine (mmol/l) |
|  | Alb | Albumin (signal area) |
|  | Gp | Glycoprotein acetyls (mmol/l) |

**Supplementary Table 2:** Proportion of outliers identified in each ethnicity.

| Metabolite | South Asian |  | White European |  |
| --- | --- | --- | --- | --- |
|  | Final Sample Size | % Removed Outliers | Final Sample Size | % Removed Outliers |
| XXL-VLDL-P | 3586 | 2.90 | 3247 | 3.79 |
| XXL-VLDL-L | 3589 | 2.82 | 3250 | 3.70 |
| XXL-VLDL-PL | 3592 | 2.73 | 3253 | 3.61 |
| XXL-VLDL-C | 3605 | 2.38 | 3270 | 3.11 |
| XXL-VLDL-CE | 3615 | 2.11 | 3276 | 2.93 |
| XXL-VLDL-FC | 3591 | 2.76 | 3264 | 3.29 |
| XXL-VLDL-TG | 3584 | 2.95 | 3241 | 3.97 |
| XL-VLDL-P | 3585 | 2.92 | 3261 | 3.38 |
| XL-VLDL-L | 3587 | 2.87 | 3261 | 3.38 |
| XL-VLDL-PL | 3587 | 2.87 | 3255 | 3.56 |
| XL-VLDL-C | 3599 | 2.55 | 3259 | 3.44 |
| XL-VLDL-CE | 3600 | 2.52 | 3256 | 3.53 |
| XL-VLDL-FC | 3595 | 2.65 | 3258 | 3.47 |
| XL-VLDL-TG | 3587 | 2.87 | 3262 | 3.35 |
| L-VLDL-P | 3588 | 2.84 | 3264 | 3.29 |
| L-VLDL-L | 3592 | 2.73 | 3265 | 3.26 |
| L-VLDL-PL | 3596 | 2.63 | 3266 | 3.23 |
| L-VLDL-C | 3590 | 2.79 | 3257 | 3.50 |
| L-VLDL-CE | 3607 | 2.33 | 3256 | 3.53 |
| L-VLDL-FC | 3593 | 2.71 | 3265 | 3.26 |
| L-VLDL-TG | 3588 | 2.84 | 3262 | 3.35 |
| M-VLDL-P | 3601 | 2.49 | 3266 | 3.23 |
| M-VLDL-L | 3603 | 2.44 | 3261 | 3.38 |
| M-VLDL-PL | 3610 | 2.25 | 3268 | 3.17 |
| M-VLDL-C | 3614 | 2.14 | 3271 | 3.08 |
| M-VLDL-CE | 3616 | 2.09 | 3289 | 2.55 |
| M-VLDL-FC | 3608 | 2.30 | 3266 | 3.23 |
| M-VLDL-TG | 3589 | 2.82 | 3268 | 3.17 |
| S-VLDL-P | 3617 | 2.06 | 3295 | 2.37 |
| S-VLDL-L | 3615 | 2.11 | 3291 | 2.49 |
| S-VLDL-PL | 3627 | 1.79 | 3301 | 2.19 |
| S-VLDL-C | 3627 | 1.79 | 3303 | 2.13 |
| S-VLDL-CE | 3627 | 1.79 | 3309 | 1.96 |
| S-VLDL-FC | 3617 | 2.06 | 3300 | 2.22 |
| S-VLDL-TG | 3606 | 2.36 | 3271 | 3.08 |
| XS-VLDL-P | 3623 | 1.90 | 3316 | 1.75 |
| XS-VLDL-L | 3627 | 1.79 | 3315 | 1.78 |
| XS-VLDL-PL | 3627 | 1.79 | 3311 | 1.90 |
| XS-VLDL-C | 3632 | 1.65 | 3305 | 2.07 |
| XS-VLDL-CE | 3634 | 1.60 | 3308 | 1.99 |
| XS-VLDL-FC | 3626 | 1.81 | 3308 | 1.99 |
| XS-VLDL-TG | 3614 | 2.14 | 3295 | 2.37 |
| IDL-P | 3628 | 1.76 | 3312 | 1.87 |
| IDL-L | 3629 | 1.73 | 3310 | 1.93 |
| IDL-PL | 3628 | 1.76 | 3307 | 2.01 |
| IDL-C | 3628 | 1.76 | 3311 | 1.90 |
| IDL-CE | 3630 | 1.71 | 3310 | 1.93 |
| IDL-FC | 3624 | 1.87 | 3318 | 1.69 |
| IDL-TG | 3619 | 2.00 | 3303 | 2.13 |
| L-LDL-P | 3630 | 1.71 | 3307 | 2.01 |
| L-LDL-L | 3630 | 1.71 | 3308 | 1.99 |
| L-LDL-PL | 3632 | 1.65 | 3304 | 2.10 |
| L-LDL-C | 3627 | 1.79 | 3307 | 2.01 |
| L-LDL-CE | 3628 | 1.76 | 3306 | 2.04 |
| L-LDL-FC | 3622 | 1.92 | 3315 | 1.78 |
| L-LDL-TG | 3621 | 1.95 | 3310 | 1.93 |
| M-LDL-P | 3629 | 1.73 | 3307 | 2.01 |
| M-LDL-L | 3628 | 1.76 | 3310 | 1.93 |
| M-LDL-PL | 3634 | 1.60 | 3307 | 2.01 |
| M-LDL-C | 3628 | 1.76 | 3308 | 1.99 |
| M-LDL-CE | 3628 | 1.76 | 3304 | 2.10 |
| M-LDL-FC | 3634 | 1.60 | 3307 | 2.01 |
| M-LDL-TG | 3621 | 1.95 | 3310 | 1.93 |
| S-LDL-P | 3631 | 1.68 | 3313 | 1.84 |
| S-LDL-L | 3636 | 1.54 | 3306 | 2.04 |
| S-LDL-PL | 3635 | 1.57 | 3309 | 1.96 |
| S-LDL-C | 3631 | 1.68 | 3310 | 1.93 |
| S-LDL-CE | 3623 | 1.90 | 3308 | 1.99 |
| S-LDL-FC | 3630 | 1.71 | 3311 | 1.90 |
| S-LDL-TG | 3628 | 1.76 | 3303 | 2.13 |
| XL-HDL-P | 3646 | 1.27 | 3340 | 1.04 |
| XL-HDL-L | 3645 | 1.30 | 3343 | 0.95 |
| XL-HDL-PL | 3649 | 1.19 | 3345 | 0.89 |
| XL-HDL-C | 3640 | 1.44 | 3340 | 1.04 |
| XL-HDL-CE | 3646 | 1.27 | 3344 | 0.92 |
| XL-HDL-FC | 3633 | 1.62 | 3338 | 1.10 |
| XL-HDL-TG | 3642 | 1.38 | 3323 | 1.54 |
| L-HDL-P | 3646 | 1.27 | 3340 | 1.04 |
| L-HDL-L | 3645 | 1.30 | 3340 | 1.04 |
| L-HDL-PL | 3648 | 1.22 | 3342 | 0.98 |
| L-HDL-C | 3644 | 1.33 | 3342 | 0.98 |
| L-HDL-CE | 3647 | 1.25 | 3342 | 0.98 |
| L-HDL-FC | 3647 | 1.25 | 3339 | 1.07 |
| L-HDL-TG | 3638 | 1.49 | 3325 | 1.48 |
| M-HDL-P | 3645 | 1.30 | 3320 | 1.63 |
| M-HDL-L | 3642 | 1.38 | 3321 | 1.60 |
| M-HDL-PL | 3645 | 1.30 | 3312 | 1.87 |
| M-HDL-C | 3640 | 1.44 | 3332 | 1.27 |
| M-HDL-CE | 3638 | 1.49 | 3330 | 1.33 |
| M-HDL-FC | 3643 | 1.35 | 3329 | 1.36 |
| M-HDL-TG | 3634 | 1.60 | 3315 | 1.78 |
| S-HDL-P | 3635 | 1.57 | 3324 | 1.51 |
| S-HDL-L | 3633 | 1.62 | 3324 | 1.51 |
| S-HDL-PL | 3635 | 1.57 | 3307 | 2.01 |
| S-HDL-C | 3625 | 1.84 | 3307 | 2.01 |
| S-HDL-CE | 3618 | 2.03 | 3317 | 1.72 |
| S-HDL-FC | 3636 | 1.54 | 3310 | 1.93 |
| S-HDL-TG | 3618 | 2.03 | 3311 | 1.90 |
| VLDL_D | 3644 | 1.33 | 3326 | 1.45 |
| LDL_D | 3625 | 1.84 | 3234 | 4.18 |
| HDL_D | 3668 | 0.68 | 3354 | 0.62 |
| Serum_C | 3638 | 1.49 | 3309 | 1.96 |
| VLDL_C | 3626 | 1.81 | 3292 | 2.46 |
| Remnant_C | 3626 | 1.81 | 3314 | 1.81 |
| LDL_C | 3625 | 1.84 | 3304 | 2.10 |
| HDL_C | 3643 | 1.35 | 3340 | 1.04 |
| HDL2_C | 3650 | 1.16 | 3338 | 1.10 |
| HDL3_C | 3631 | 1.68 | 3323 | 1.54 |
| EstC | 3640 | 1.44 | 3311 | 1.90 |
| FreeC | 3640 | 1.44 | 3306 | 2.04 |
| Serum_TG | 3611 | 2.22 | 3278 | 2.87 |
| **VLDL-TG** | 3601 | 2.49 | 3266 | 3.23 |
| **LDL-TG** | 3621 | 1.95 | 3308 | 1.99 |
| **HDL-TG** | 3618 | 2.03 | 3319 | 1.66 |
| **TotPG** | 3642 | 1.38 | 3326 | 1.45 |
| **PC** | 3648 | 1.22 | 3311 | 1.90 |
| **SM** | 3641 | 1.41 | 3312 | 1.87 |
| **TotCho** | 3637 | 1.52 | 3320 | 1.63 |
| **ApoA1** | 3642 | 1.38 | 3328 | 1.39 |
| **ApoB** | 3626 | 1.81 | 3312 | 1.87 |
| **TotFA** | 3630 | 1.71 | 3311 | 1.90 |
| **DHA** | 3602 | 2.46 | 3303 | 2.13 |
| **LA** | 3638 | 1.49 | 3323 | 1.54 |
| **FAw3** | 3616 | 2.09 | 3305 | 2.07 |
| **FAw6** | 3639 | 1.46 | 3314 | 1.81 |
| **PUFA** | 3640 | 1.44 | 3308 | 1.99 |
| **MUFA** | 3611 | 2.22 | 3298 | 2.28 |
| **SFA** | 3622 | 1.92 | 3302 | 2.16 |
| **Lac** | 3516 | 4.79 | 3244 | 3.88 |
| **Pyr** | 3578 | 3.11 | 3288 | 2.58 |
| **Cit** | 3641 | 1.41 | 3302 | 2.16 |
| **Glol** | 3624 | 1.87 | 3293 | 2.43 |
| **Ala** | 3640 | 1.44 | 3338 | 1.10 |
| **Gln** | 3642 | 1.38 | 3320 | 1.63 |
| **Gly** | 3629 | 1.73 | 3328 | 1.39 |
| **His** | 3634 | 1.60 | 3324 | 1.51 |
| **Ile** | 3609 | 2.27 | 3284 | 2.70 |
| **Leu** | 3635 | 1.57 | 3309 | 1.96 |
| **Val** | 3621 | 1.95 | 3319 | 1.66 |
| **Phe** | 3640 | 1.44 | 3328 | 1.39 |
| **Tyr** | 3649 | 1.19 | 3320 | 1.63 |
| **Ace** | 3594 | 2.68 | 3280 | 2.81 |
| **bOHBut** | 3519 | 4.71 | 3184 | 5.66 |
| **Crea** | 3654 | 1.06 | 3329 | 1.36 |
| **Alb** | 3472 | 5.98 | 3158 | 6.43 |
| **Gp** | 3630 | 1.71 | 3296 | 2.34 |

**Supplementary Table 3:**
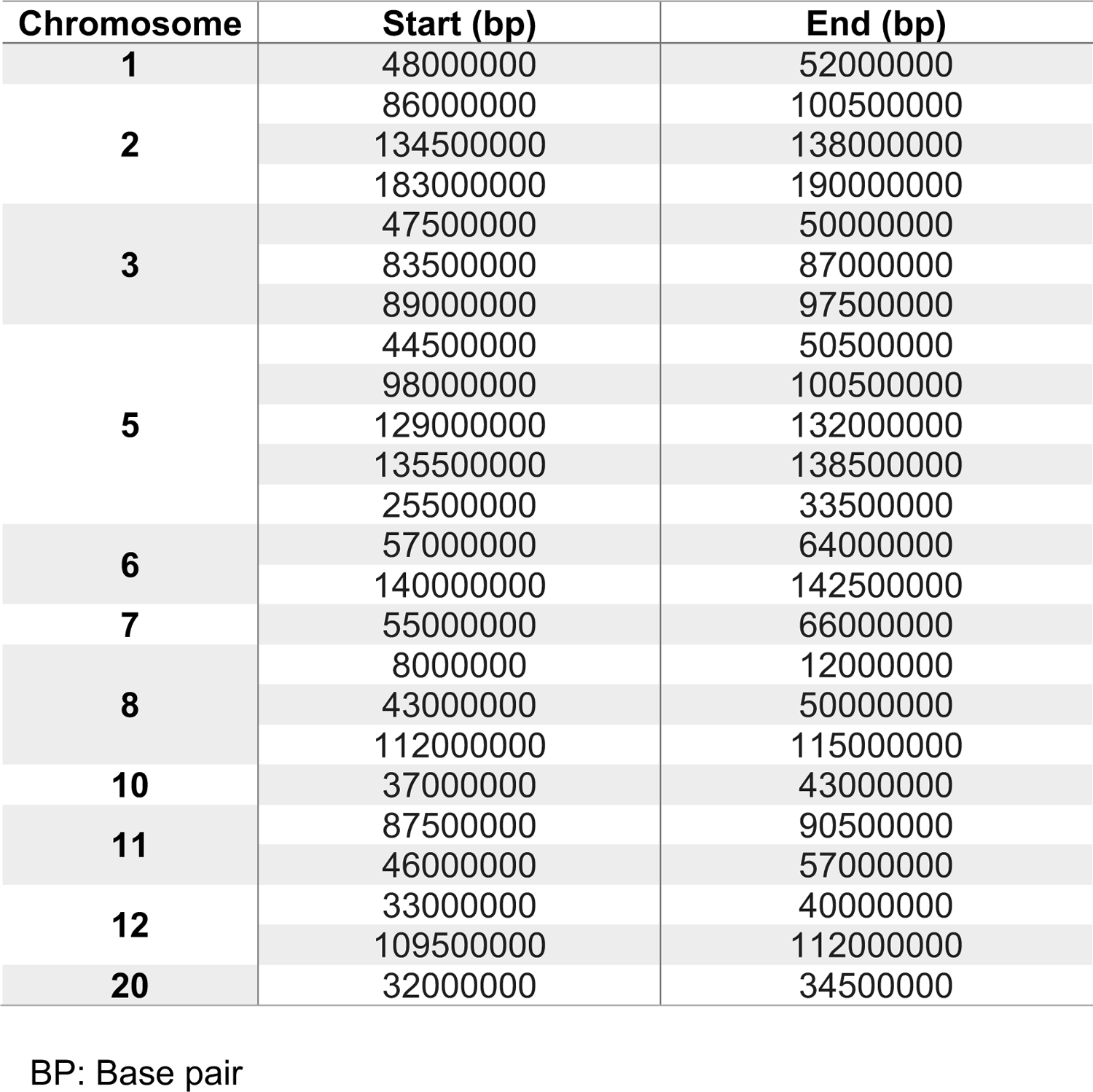
Regions of high linkage disequilibrium (LD) excluded from PCA of genetic data.

**Supplementary Table 4:** Absolute deviations in λ from 1 in each ethnicity. Average differences of λ values from 1 in each ethnicity averaged across 146 metabolite values. MAF: Minor Allele Frequency. A: Map of the Indian subcontinent with the location of each South Asian 1000G population illustrated. Brackets represent the country in which the sample was taken from. Colours of labels illustrate data points in PCA plots B: PC1 vs PC2. C: PC2 vs PC3. Base map for panel A was obtained from the *rworld* map package in R studio.

| **MAF cut off** | **White European** | **South Asian** |
| --- | --- | --- |
| **MAF <0.001** | 0.184 | 0.047 |
| **0.001 ≤ MAF <0.005** | 0.071 | 0.028 |
| **0.005 ≤ MAF <0.01** | 0.070 | 0.030 |
| **0.01 ≤ MAF <0.05** | 0.029 | 0.017 |
| **0.05 ≤ MAF <0.10** | 0.020 | 0.026 |
| **0.10 ≤ MAF <0.15** | 0.023 | 0.018 |
| **0.15 ≤ MAF <0.20** | 0.011 | 0.010 |
| **0.20 ≤ MAF <0.25** | 0.011 | 0.010 |
| **0.25 ≤ MAF <0.30** | 0.015 | 0.012 |
| **0.30 ≤ MAF <0.35** | 0.016 | 0.014 |
| **0.35 ≤ MAF <0.40** | 0.022 | 0.014 |
| **0.40 ≤ MAF <0.45** | 0.026 | 0.020 |
| **0.45 ≤ MAF <0.50** | 0.022 | 0.023 |

Average differences of $\lambda$ values from 1 in each ethnicity averaged across 146 metabolite values. MAF: Minor Allele Frequency.

**Supplementary Table 5:**
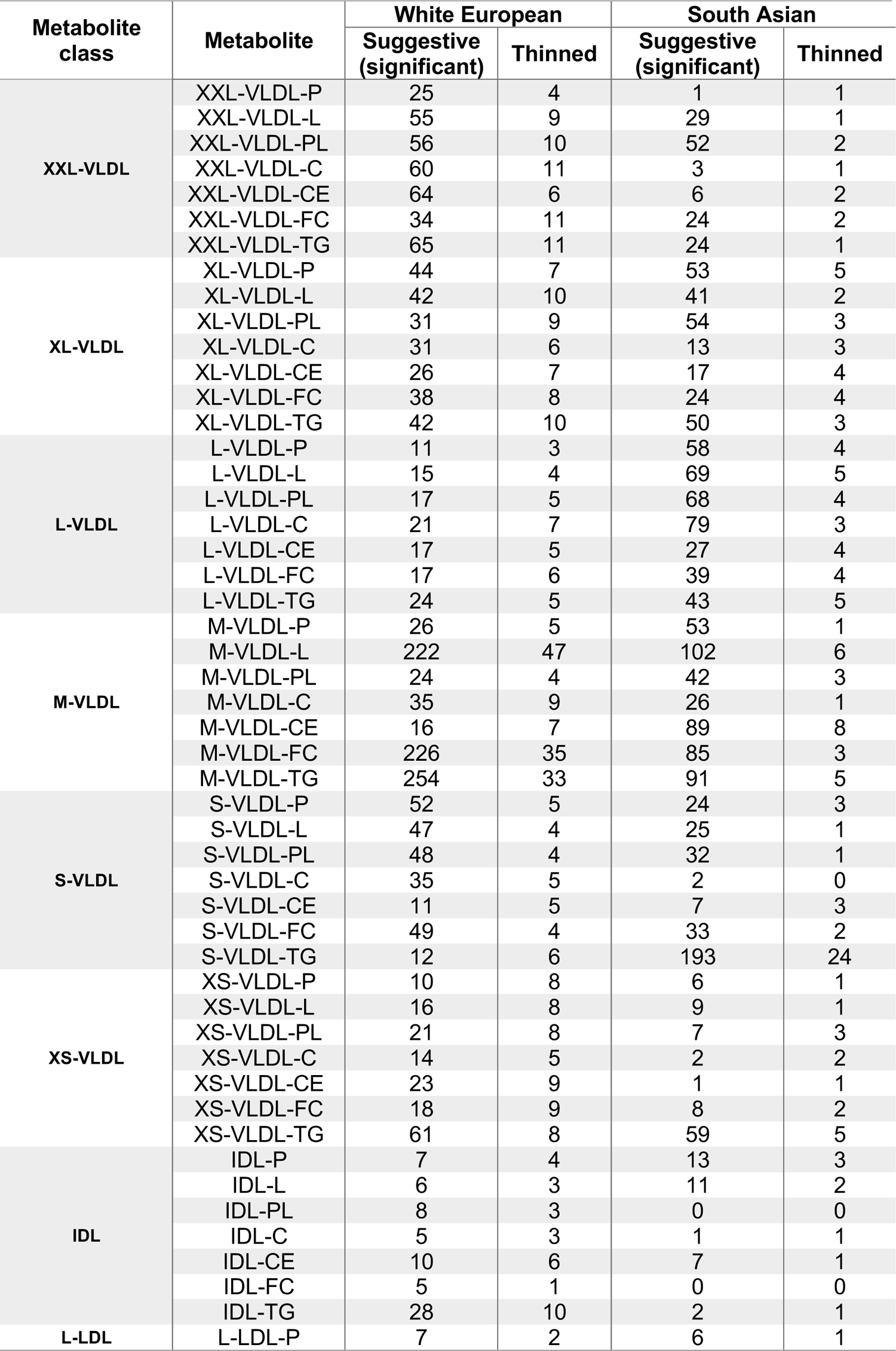
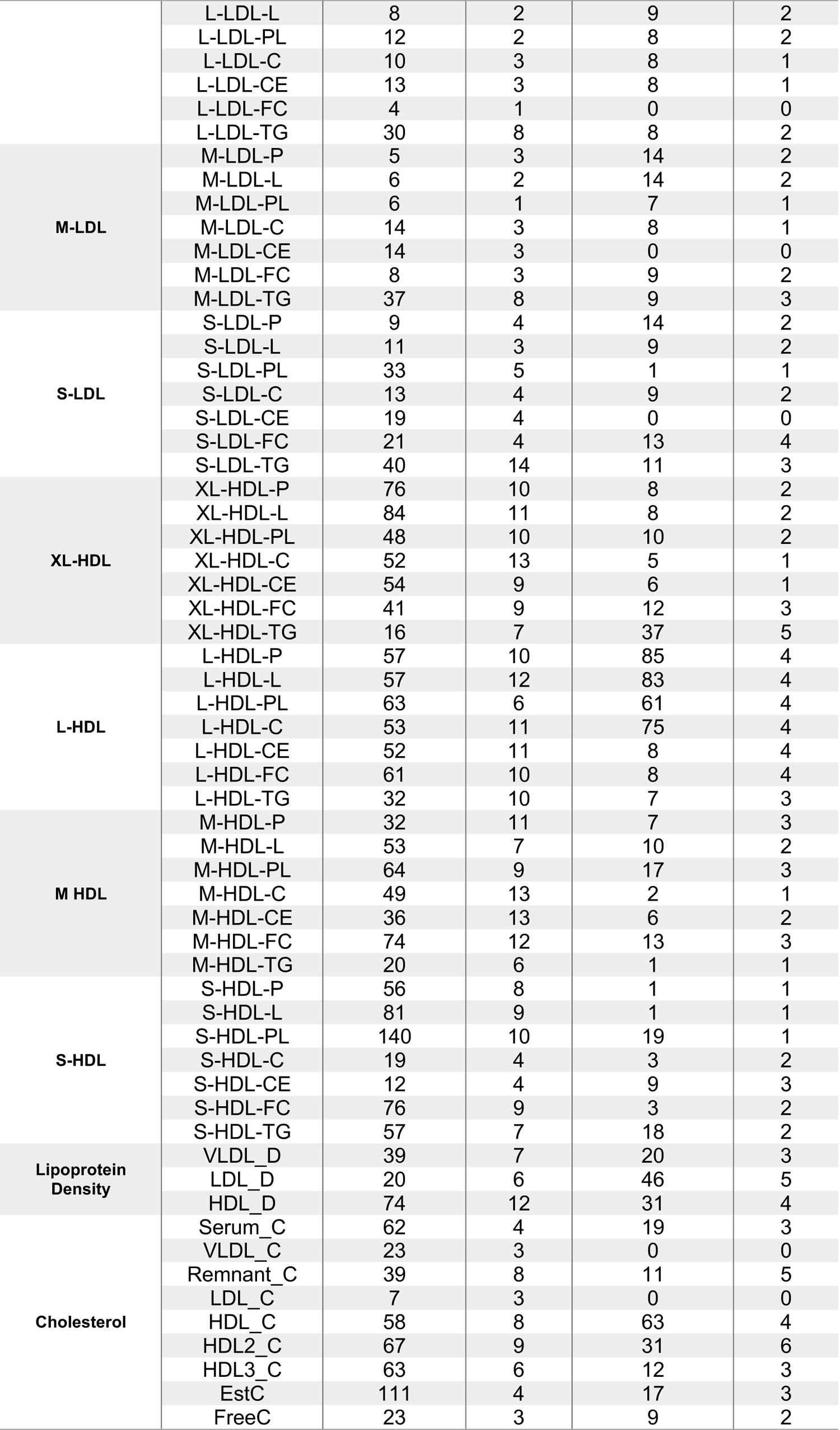
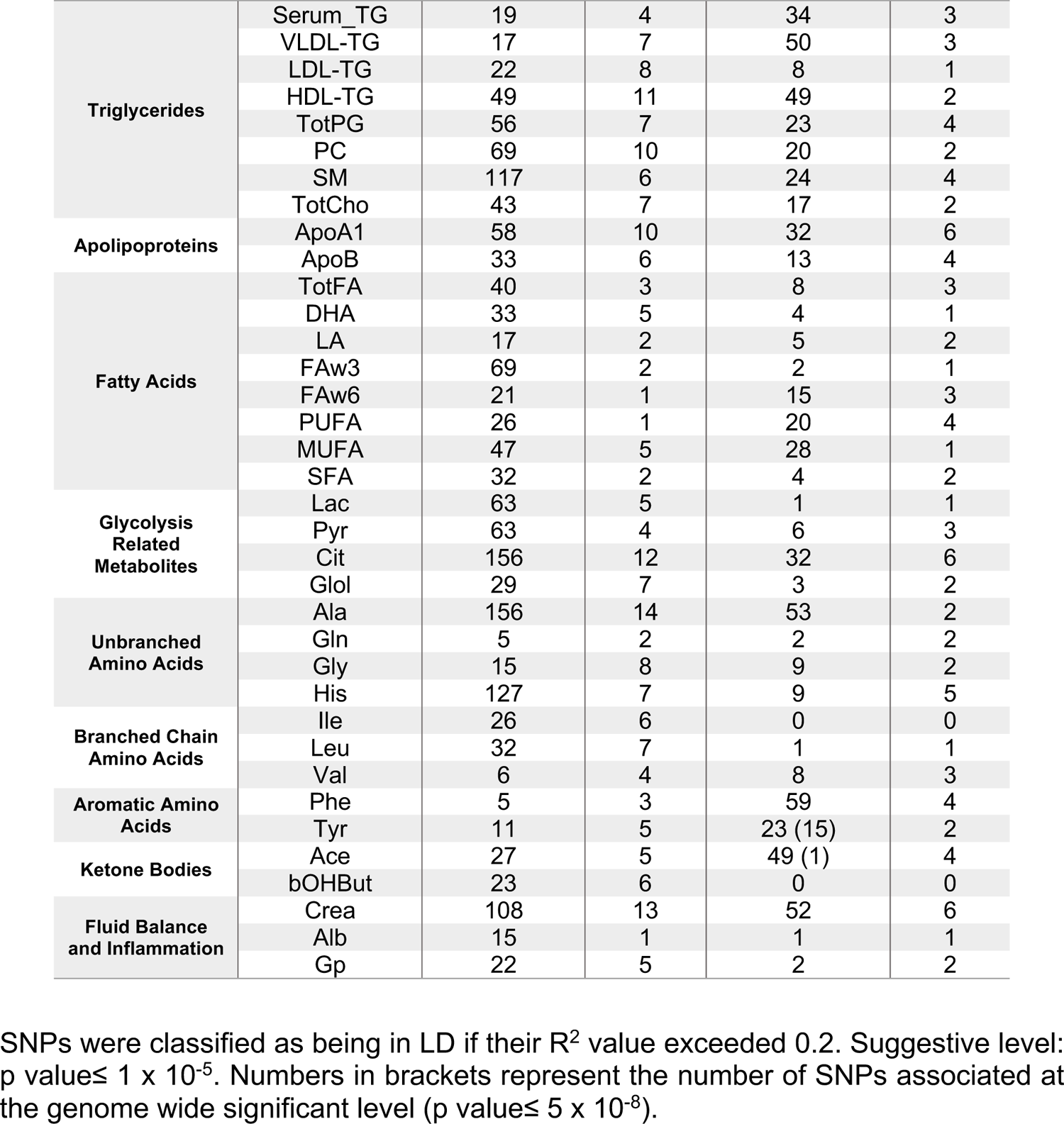
Number of SNPs identified in each ethnicity.

**Supplementary Table 6:**
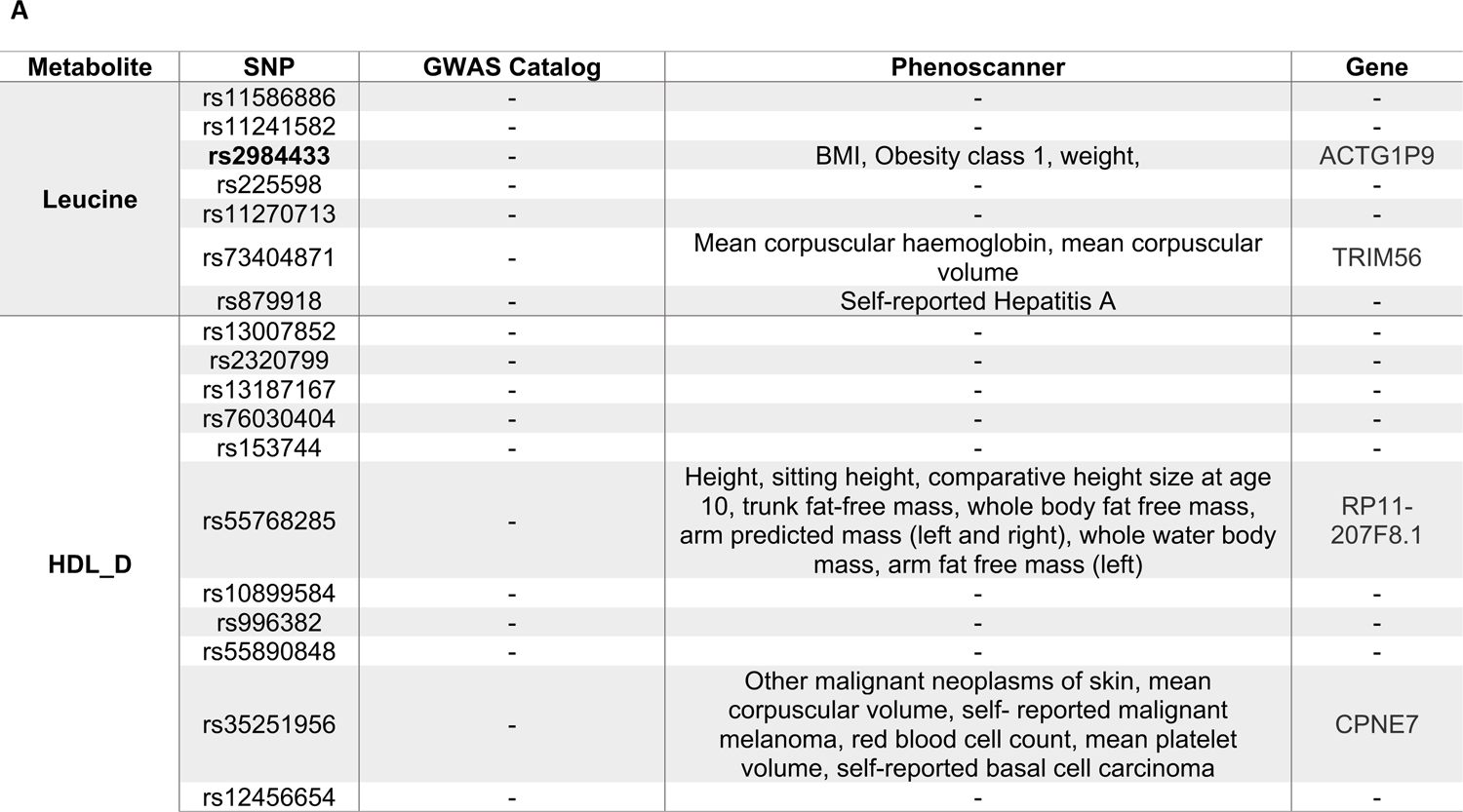
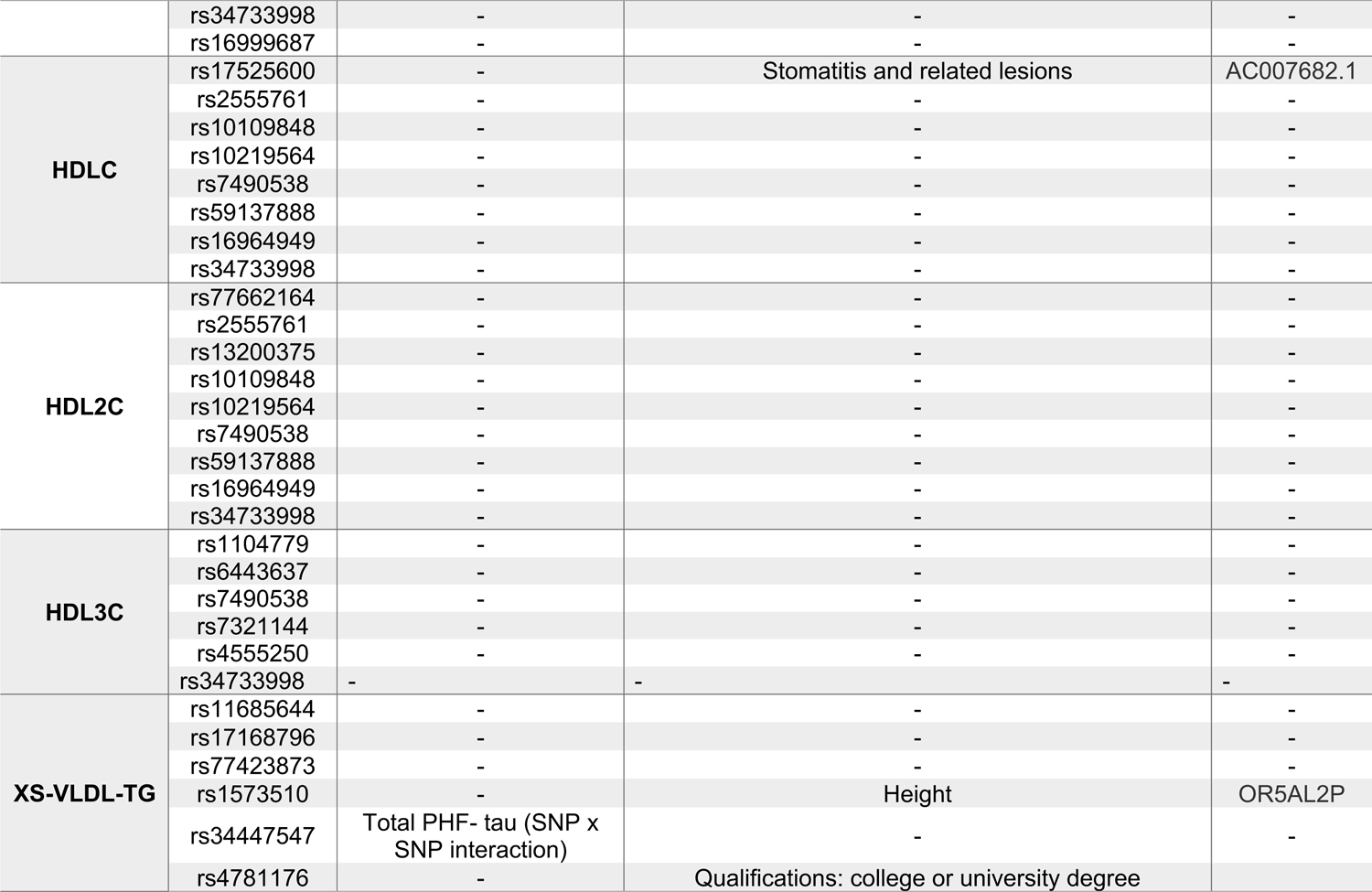
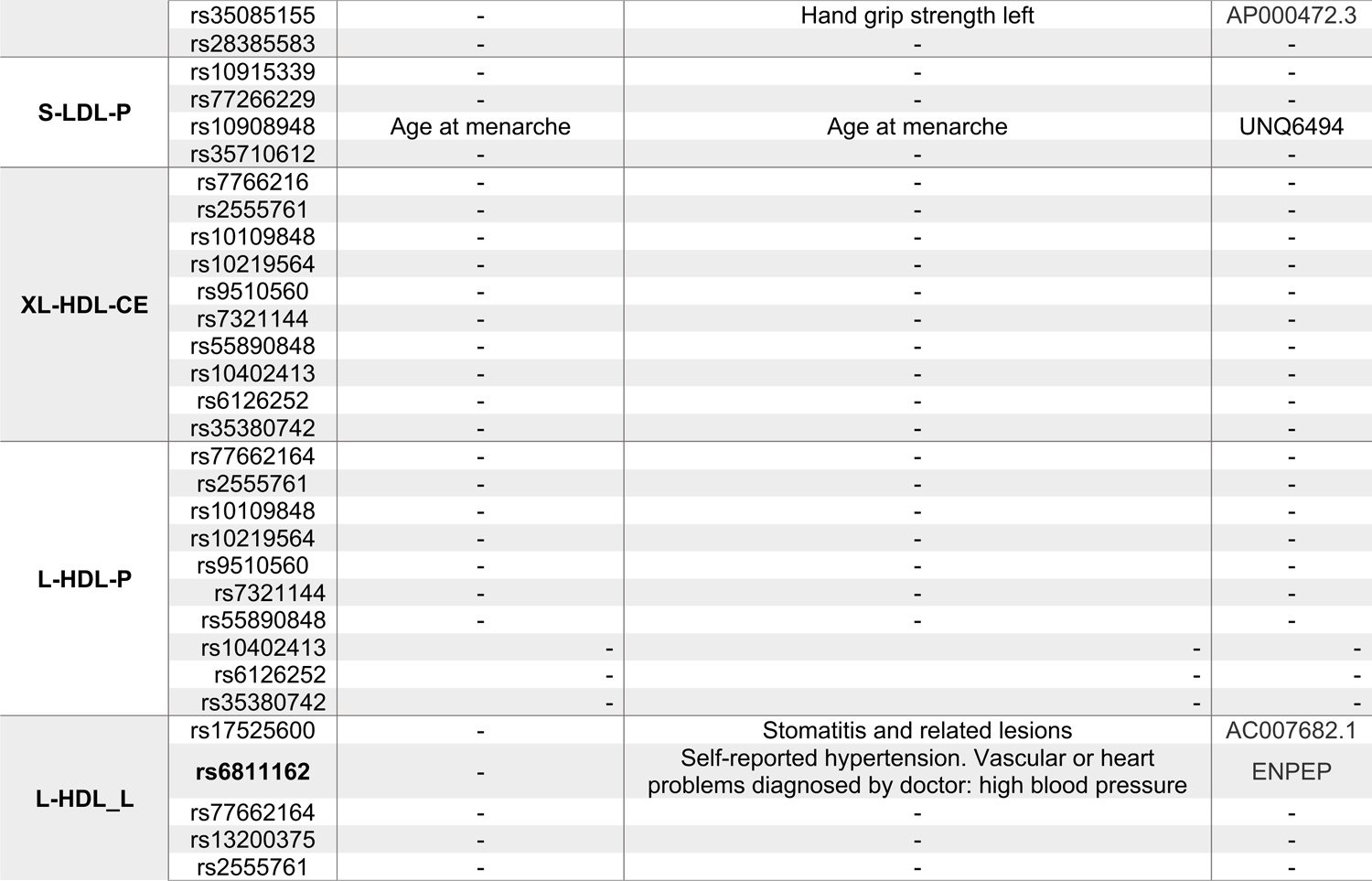
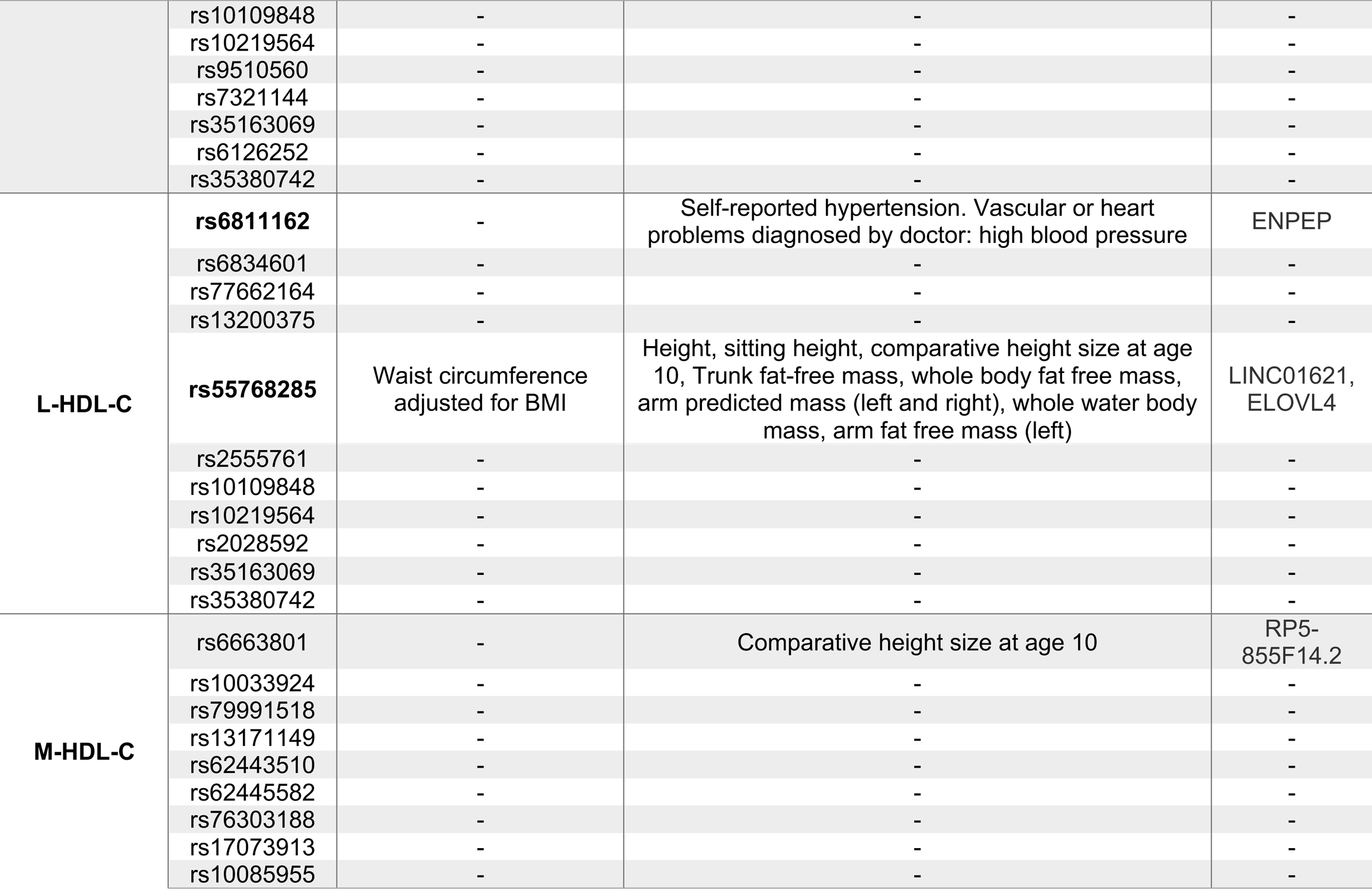
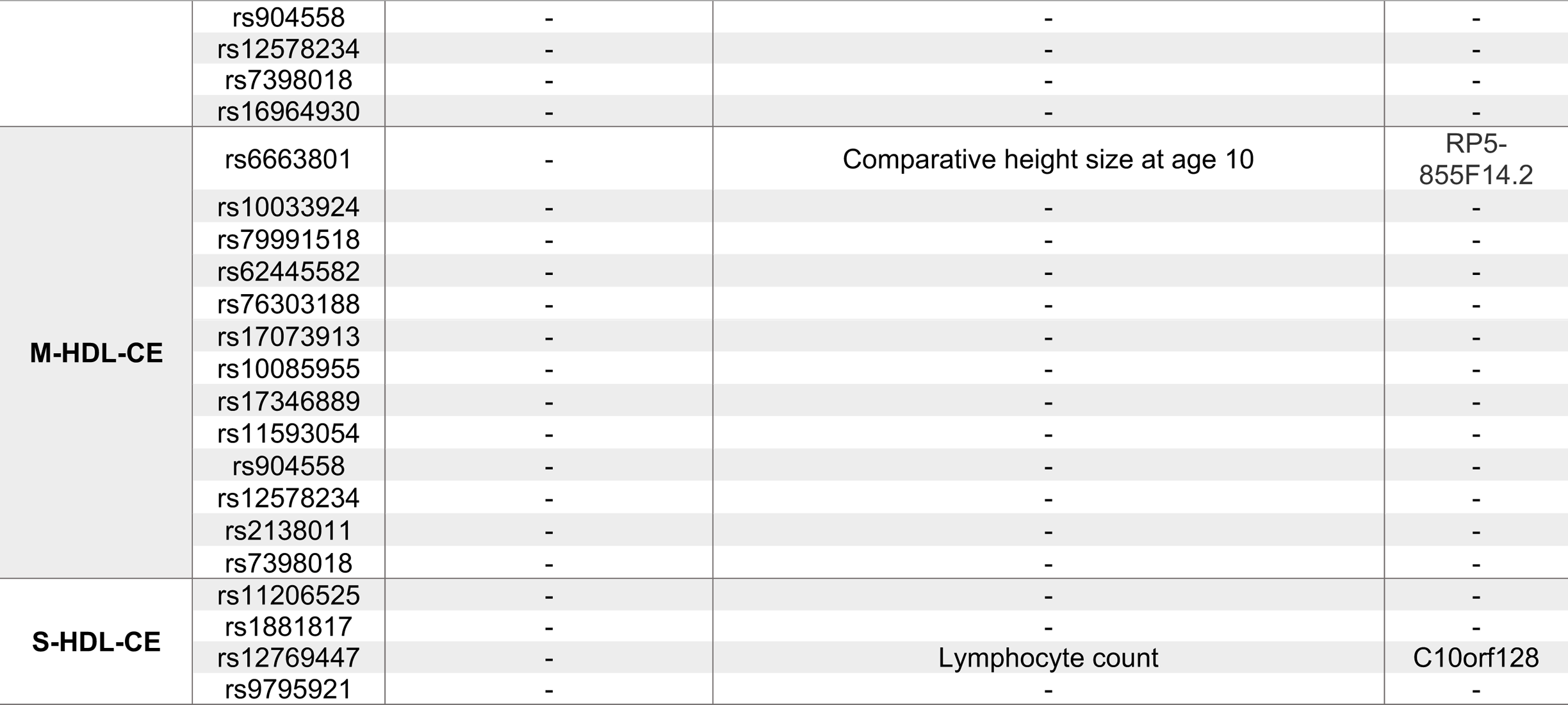
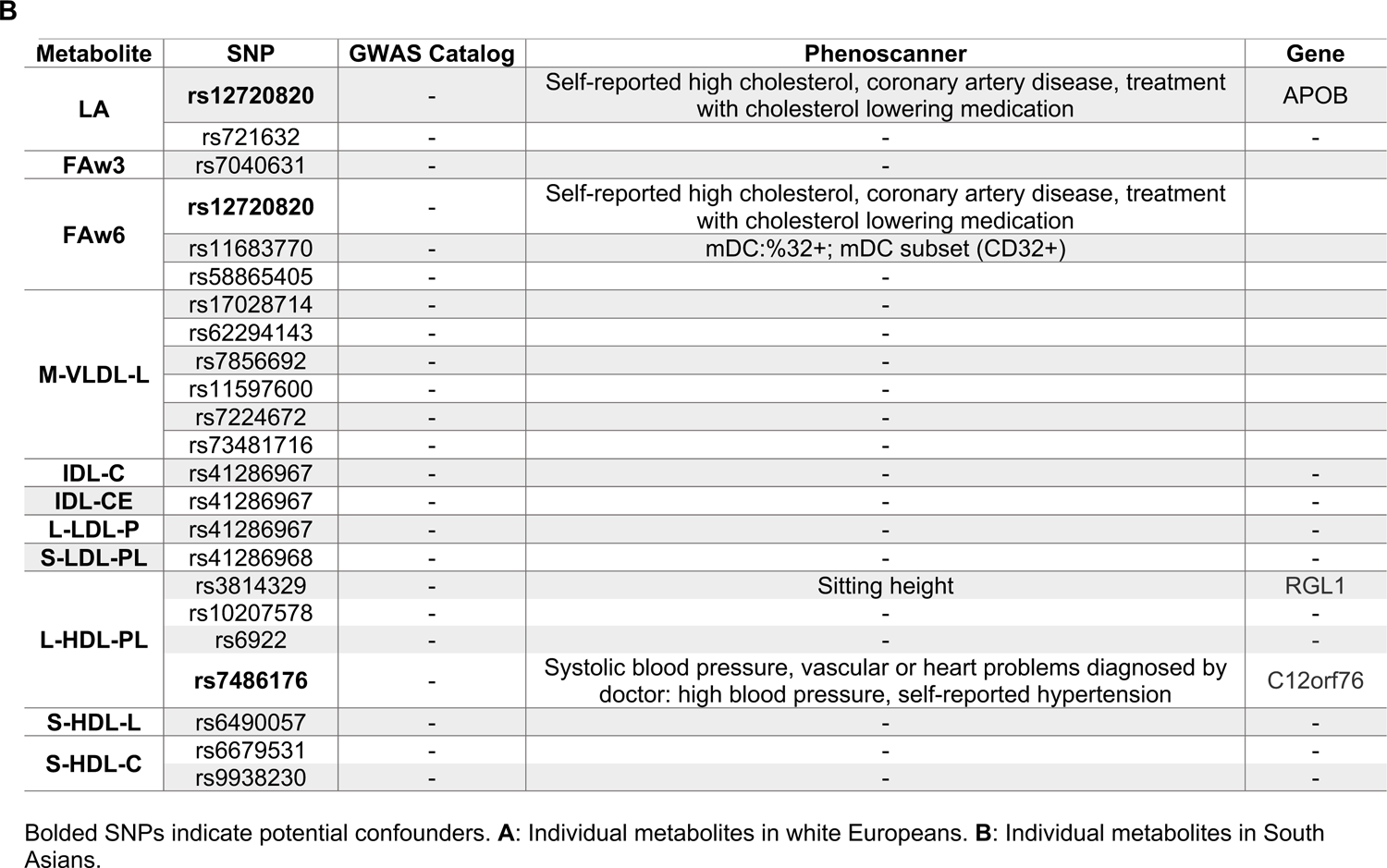
Investigation of pleiotropy in the GWAS Catalog and Phenoscanner databases.

**Supplementary Table 7:** Percentage of variation explained by PC1 in each metabolite class.

| **Metabolite class** | **White Europeans** | **South Asians** |
| --- | --- | --- |
| **XS-VLDL** | 88 | 91.5 |
| **S-VLDL** | 96.8 | 91.8 |
| **M-VLDL** | 98.9 | 95.6 |
| **L-VLDL** | 98.5 | 99.0 |
| **XL-VLDL** | 99.1 | 96.7 |
| **XXL-VLDL** | 96.7 | 95.1 |
| **All VLDL classes** | 91.9 | 86 |
| **IDL** | 85.8 | 92.1 |
| **S-LDL** | 98.0 | 97.5 |
| **M-LDL** | 95.5 | 95.3 |
| **L-LDL** | 92.5 | 91.7 |
| **All LDL classes** | 95.0 | 94.5 |
| **S-HDL** | 57.3 | 56 |
| **M-HDL** | 92.1 | 97.3 |
| **L-HDL** | 97.0 | 91.9 |
| **XL-HDL** | 95.2 | 97.8 |
| **All HDL classes** | 49.2 | 97.8 |
| **Glycolysis Related Metabolites** | 63.9 | 51.7 |
| **Fatty Acids** | 82.7 | 87.4 |
| **Lipoprotein Density** | 80.9 | 72.2 |
| **Cholesterol** | 69.7 | 59.7 |
| **Glycerides and Phospholipids** | 79.1 | 81.5 |
| **Branched Amino Acids** | 78.8 | 79.3 |
| **Unbranched Amino Acids** | 48.3 | 84.8 |
| **All Amino Acids** | 45.8 | 66 |

Percentage of variation explained by PC1 in the PCA analysis of each metabolite class following the removal of extreme outliers (3 x IQR). GWAS were performed on all classes with $\geq 2$ metabolites where PC1 explained $\geq 70\%$ of the variation.

**Supplementary Table 8:**
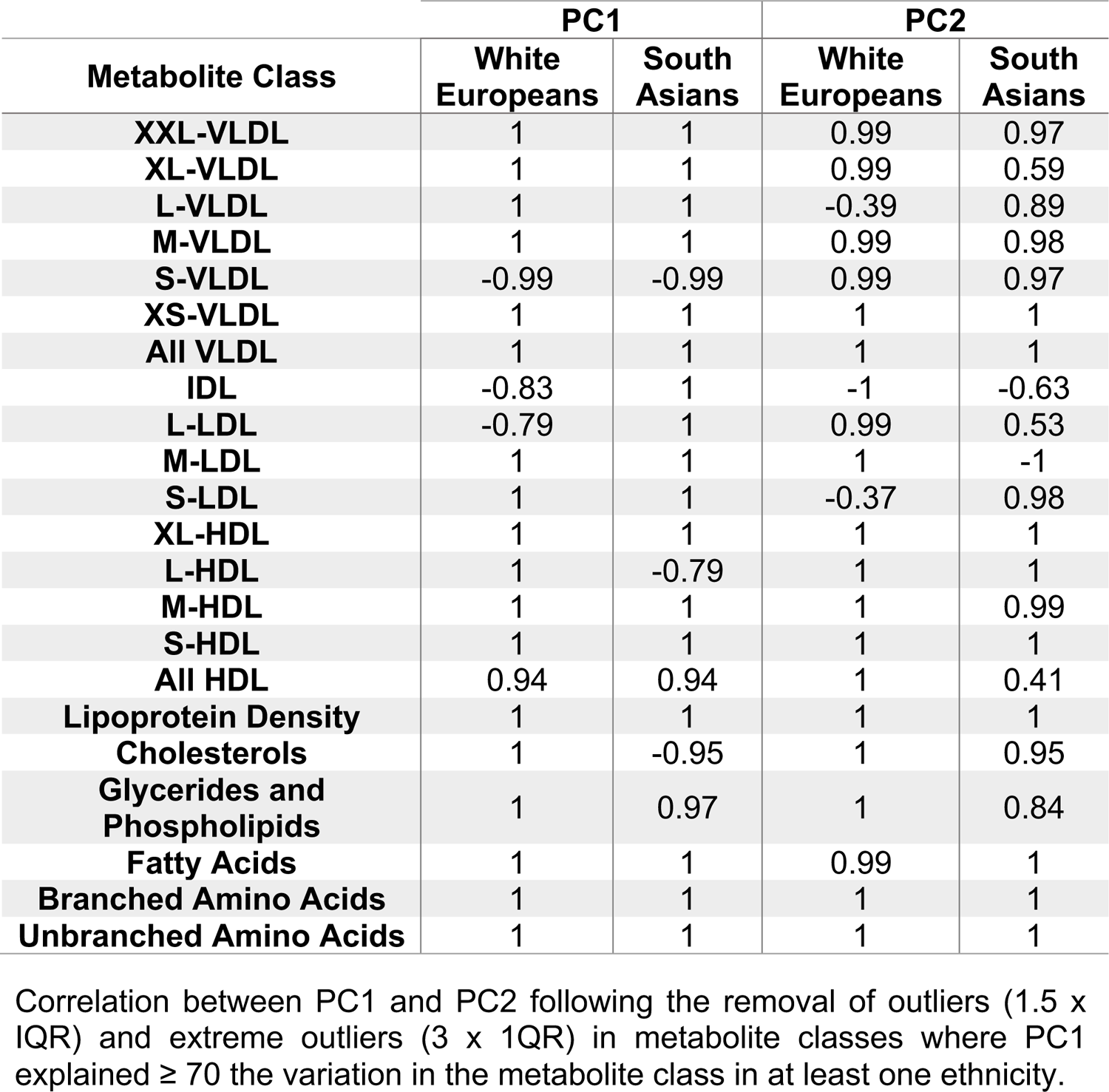
Correlation between PC1 and PC2 following outlier removal.

| Metabolite Class | PC1 |  | PC2 |  |
| --- | --- | --- | --- | --- |
|  | White Europeans | South Asians | White Europeans | South Asians |
| **XXL-VLDL** | 1 | 1 | 0.99 | 0.97 |
| **XL-VLDL** | 1 | 1 | 0.99 | 0.59 |
| **L-VLDL** | 1 | 1 | -0.39 | 0.89 |
| **M-VLDL** | 1 | 1 | 0.99 | 0.98 |
| **S-VLDL** | -0.99 | -0.99 | 0.99 | 0.97 |
| **XS-VLDL** | 1 | 1 | 1 | 1 |
| **All VLDL** | 1 | 1 | 1 | 1 |
| **IDL** | -0.83 | 1 | -1 | -0.63 |
| **L-LDL** | -0.79 | 1 | 0.99 | 0.53 |
| **M-LDL** | 1 | 1 | 1 | -1 |
| **S-LDL** | 1 | 1 | -0.37 | 0.98 |
| **XL-HDL** | 1 | 1 | 1 | 1 |
| **L-HDL** | 1 | -0.79 | 1 | 1 |
| **M-HDL** | 1 | 1 | 1 | 0.99 |
| **S-HDL** | 1 | 1 | 1 | 1 |
| **All HDL** | 0.94 | 0.94 | 1 | 0.41 |
| **Lipoprotein Density** | 1 | 1 | 1 | 1 |
| **Cholesterols** | 1 | -0.95 | 1 | 0.95 |
| **Glycerides and Phospholipids** | 1 | 0.97 | 1 | 0.84 |
| **Fatty Acids** | 1 | 1 | 0.99 | 1 |
| **Branched Amino Acids** | 1 | 1 | 1 | 1 |
| **Unbranched Amino Acids** | 1 | 1 | 1 | 1 |

Correlation between PC1 and PC2 following the removal of outliers (1.5 x IQR) and extreme outliers (3 x 1QR) in metabolite classes where PC1 explained $\geq 70$ the variation in the metabolite class in at least one ethnicity.

**Supplementary Table 9:**
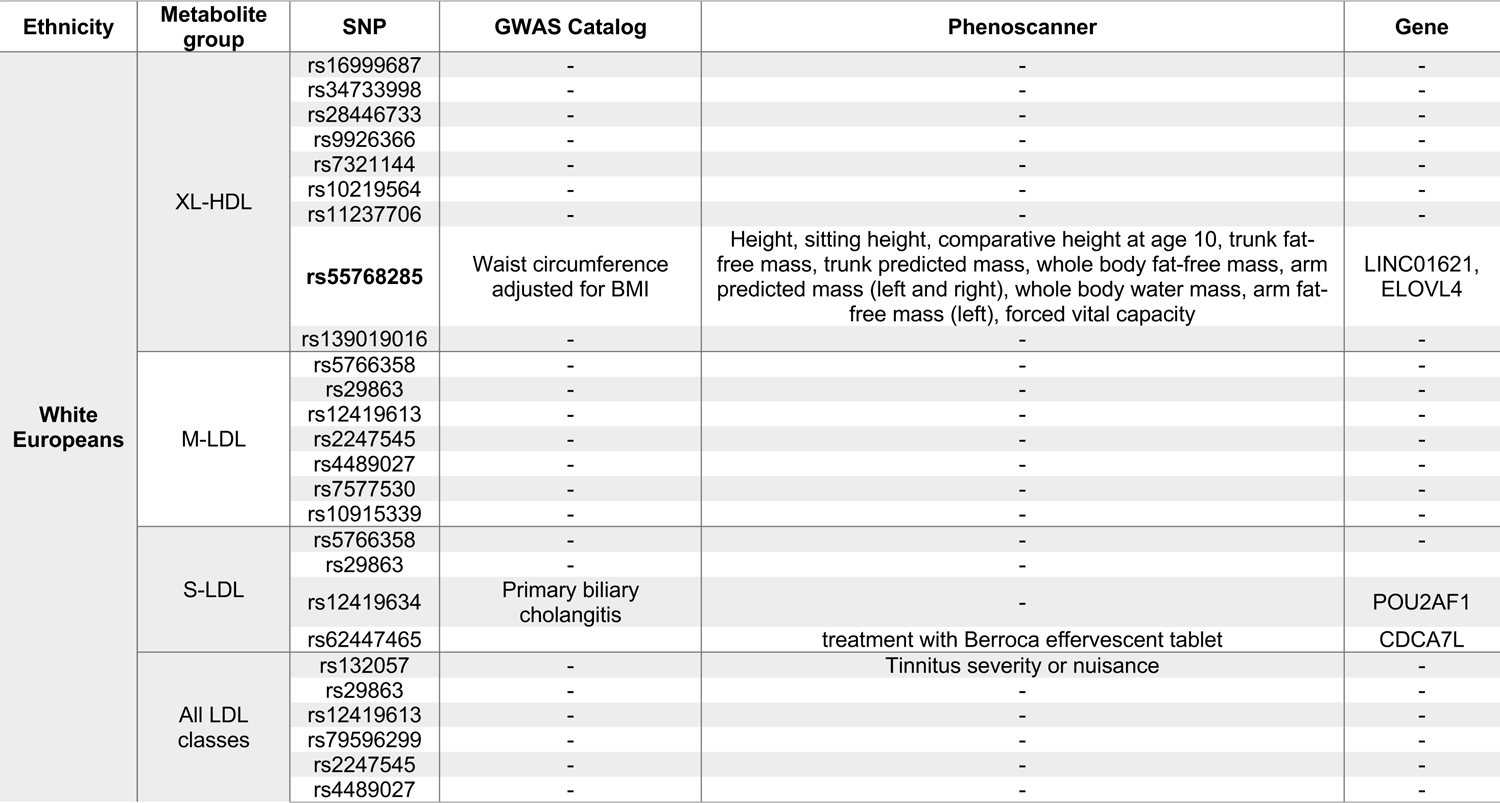
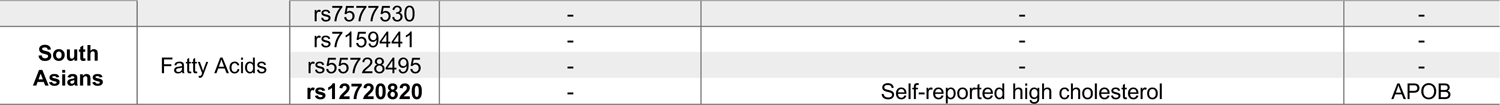
Investigation of pleiotropy in the GWAS Catalog and Phenoscanner databases for identified metabolite classes.

**Supplementary Table 10:**
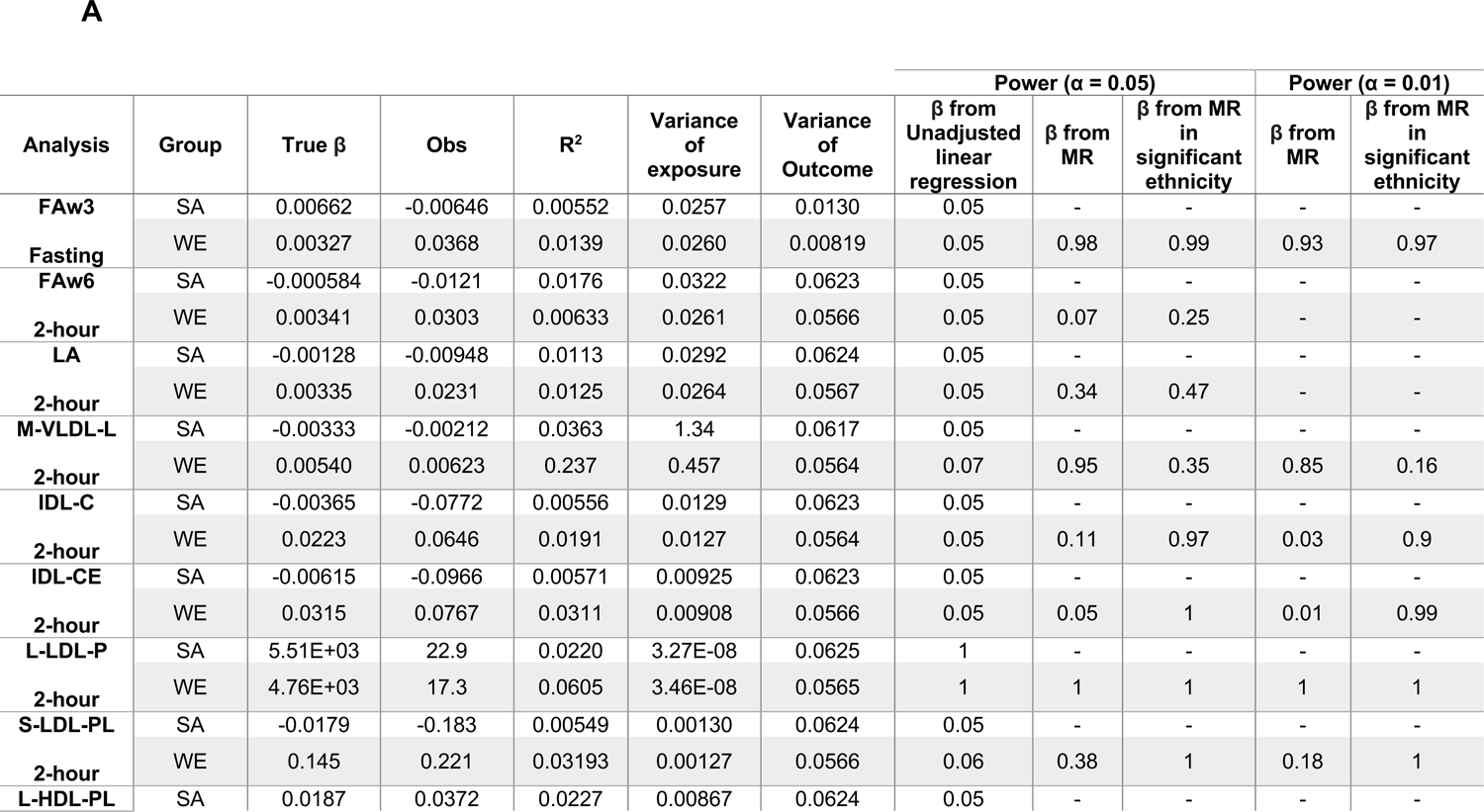
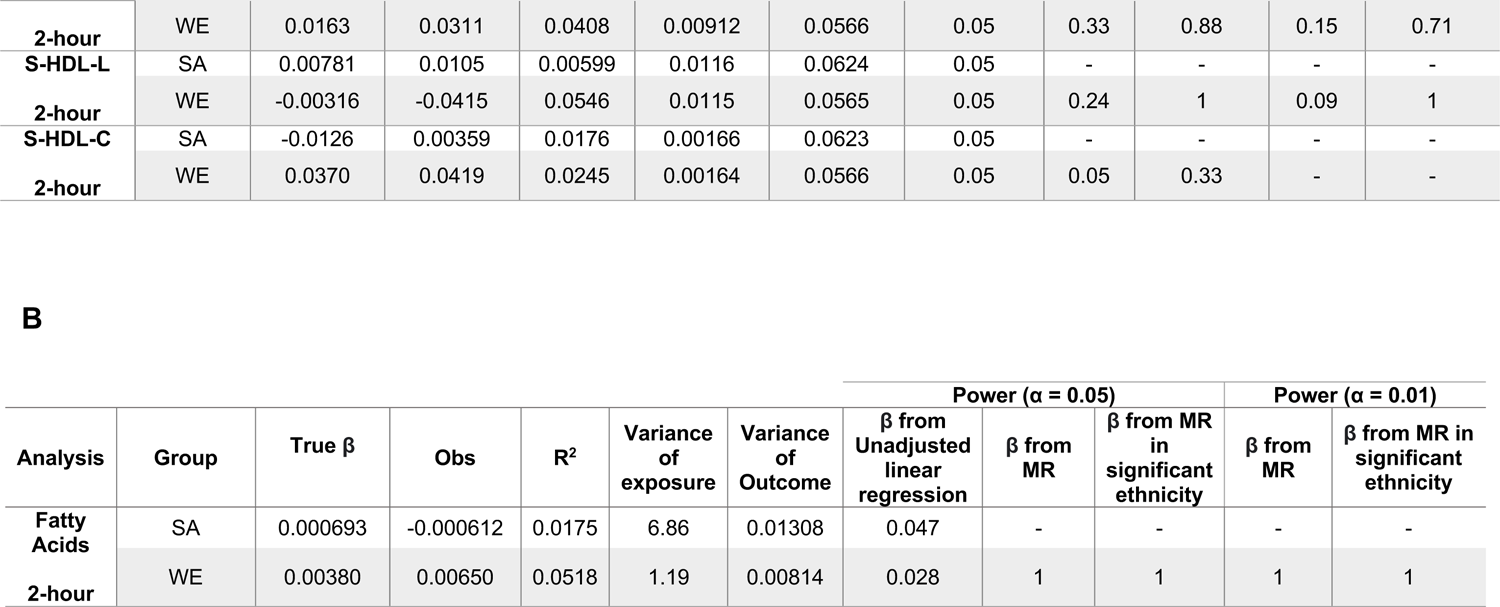
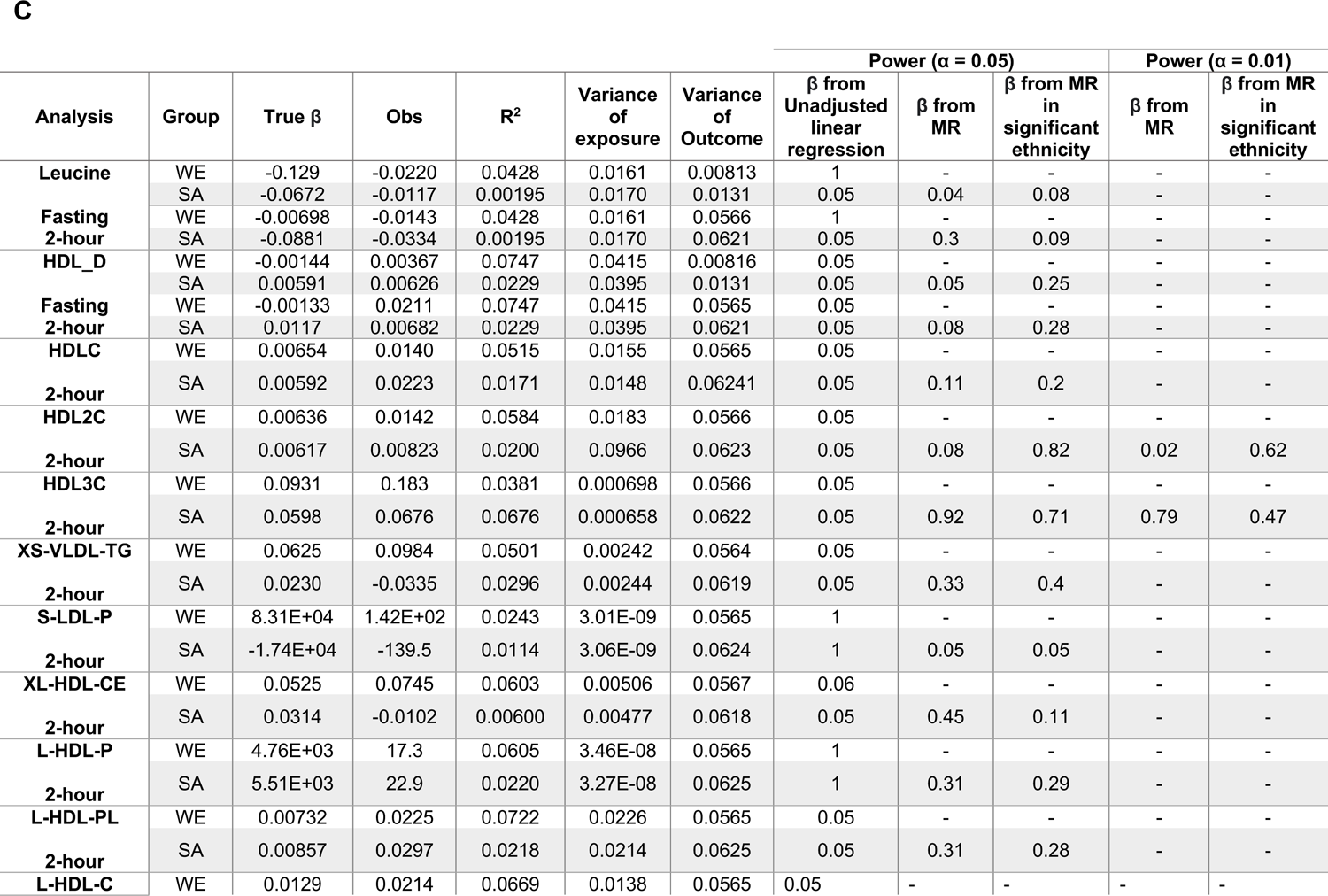
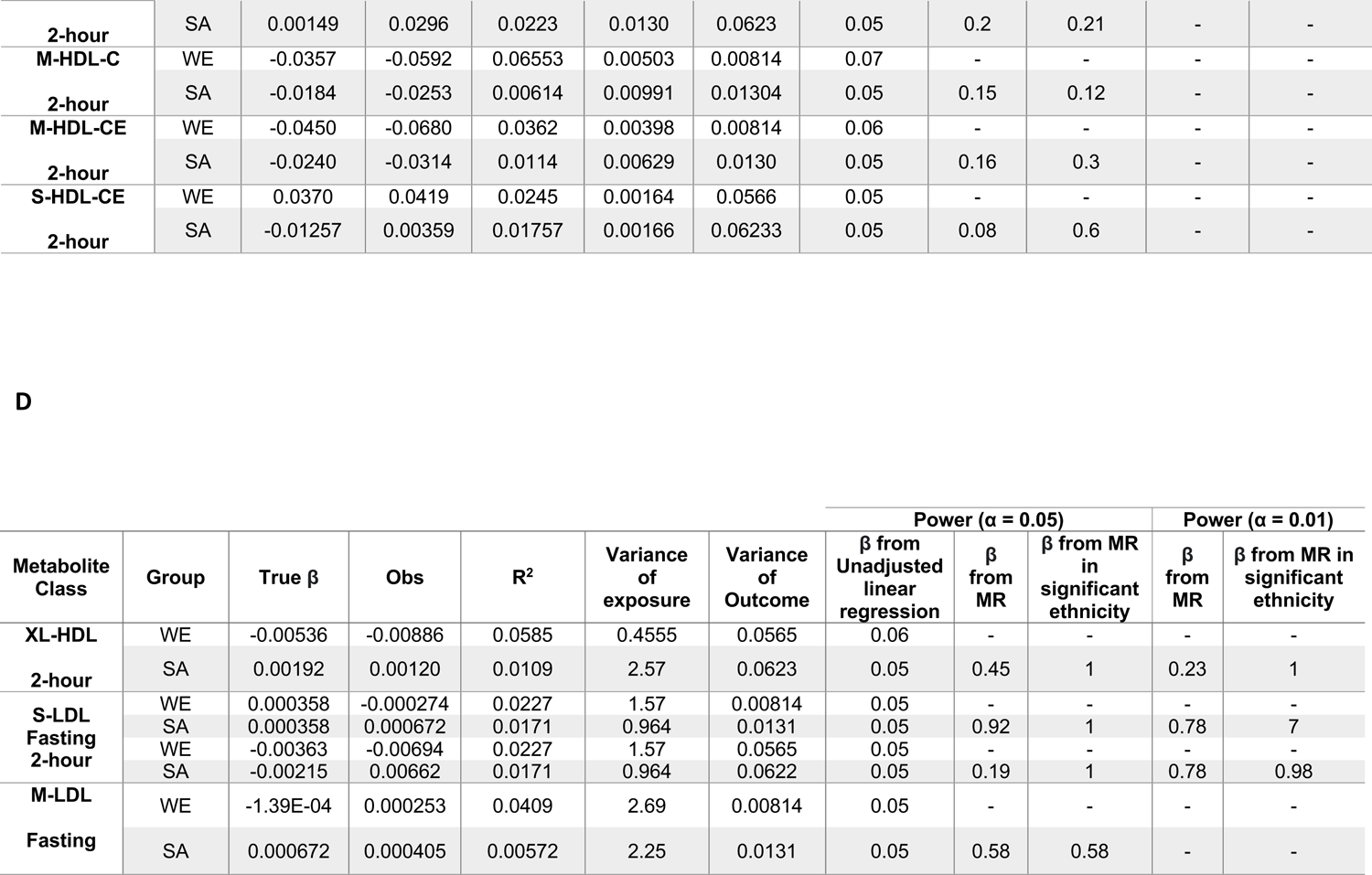
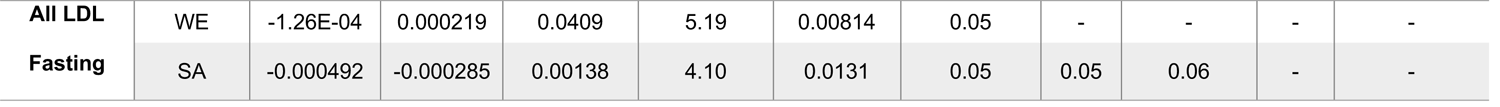
Post-hoc power analysis. Tables showing *post-hoc* MR calculation from mRND (https://shiny.cnsgenomics.com/mRnd/). Observational associations were obtained from linear regression models adjusted for maternal age (years), BMI (continuous), smoking status, multiple pregnancy, parity, and gestational age. Initial true β estimates were obtained from unadjusted linear regression models. Additional power analyses were performed in the non-significant ethnicity to determine the power to predict the β estimate obtained from the MR analyses and the power to detect the β estimate from the significant model in the alternative ethnicity. If the power from either analysis exceeded 80% then power was also calculated for α = 0.01. Obs: β from adjusted observational studies **A:** Individual metabolite analysis in South Asians. **B:** Analysis of metabolite classes in South Asians. **C:** Analysis of Individual metabolites in White Europeans. **D:** Analysis of metabolite classes in White Europeans.

